# The Human Hearing Atlas: a canonical map of human hearing

**DOI:** 10.64898/2026.09.14.26363004

**Authors:** Zakarya Hadj-Allal, Antti Aarnisalo, Antti A. Mäkitie

## Abstract

For 140 years, hearing science and otology have rested on a premise treated as a principle of human physiology: that the sense of hearing, in health and disease, comes in distinct kinds. Its standard measure, the audiogram, was collapsed into a severity average and a subjective shape label. The reduction carried the premise from otology’s founding fathers into the digital and artificial intelligence era. We tested this premise and falsified it. Human hearing does not resolve into a reproducible catalogue of types; it occupies a continuous, five-dimensional coordinate system. We call it the Human Hearing Atlas. Established across more than 4.29 million audiograms spanning 33 datasets, eight countries, 62 years and two calibration standards, the Atlas reconstructs unseen audiograms to near-decibel accuracy and preserves its clinical details. It maps within-ear hearing change over hours or decades, half of which is invisible to the conventional severity average. We demonstrate that the varying numbers of hearing types reported since 1932 are artifacts of 5 dB quantisation and cohort pooling. Finally, we establish the Law of Human Hearing States, characterized by four empirical invariants. The Atlas replaces arbitrary audiogram categories with positions and trajectories universal across individuals, populations and eras, opening a new paradigm for precision otology, genetics, epidemiology, regenerative medicine and clinical trials.

## Introduction

Every fundamental property of human physiology eventually receives its law. The nerve impulse received its equations, the heartbeat its axis, colour vision its three dimensions. The cochlea received a map of frequency. Hearing never received one.

For 140 years, human hearing has rested on a premise so old it was treated as a principle of human physiology: that human hearing, in health and disease, comes in kinds. In 1885, Arthur Hartmann introduced a graphical representation of hearing across frequency^1,2^; in 1886, he described types of hearing loss^3^. In 1922, Fowler and Wegel established the modern audiogram^4^. By 1932, Guild was sorting audiogram curves into types he named A-F through a punch-card filing system^5^; Carhart refined these types in 1945, naming them F, G, M, R and T^6^. Schuknecht linked their shapes to histopathology in 1964^7,8^. In 1980, the World Health Organization (WHO) introduced a classification grading system for hearing impairment^9^. In 1986, it codified this into international severity grades based on the pure-tone average (PTA)^10,11^.

When the era of digital medicine and artificial intelligence arrived, it inherited that premise intact. In 2007, Margolis and Saly introduced AMCLASS, an automated audiogram classification system built from 161 handcrafted conditional rules: 23 for configuration, 45 for severity, 56 for site of lesion and 37 for symmetry^12^. Other approaches replaced explicit rules with data-driven clustering, searching increasingly large audiometric datasets for recurring groups that might correspond to distinct underlying causes of hearing loss^13,14^. Deep networks then carried the same logic into machine learning, training on human-assigned audiogram labels to produce black-box classifications^15^, even though expert raters agree on those labels in fewer than half of cases^12^.

By the mid-2020s, the field had arrived not at consensus but at a crisis of its foundations^16–18^. Reported subtype counts ranged from three to twenty-nine and replicated nowhere^14,16,17,19,20^; nearly half of real audiograms fit no canonical pattern^14^; only one audiogram in five fitted the animal-model-derived phenotype classes clearly^21^; a ten-cluster taxonomy derived from 132,504 US audiograms failed to replicate intact in 109,854 independent UK audiograms^17^. Early work described two dimensions and dismissed a third as noise^22,23^; presbycusis was reported as continuous^24^, yet four decades later the proposed remedy for categorical labelling was still a single four-frequency average^25^.

By 2026, six frameworks (the baseline audiometric profile, Bisgaard profiles, WHO hearing-impairment grades, WARHICS levels, Dubno audiometric phenotypes and Parthasarathy general phenotypes) classified the audiogram using different definitions and boundaries^18^. The disagreement persisted even within the same dataset: the resulting classification depended on the analytical procedure and the number of groups sought^26^. In a 2019 analysis, the Bulletin of the WHO concluded that “there is no scientific or rational basis for the uneven steps between the various grades of severity”^11^. That standard has been revised repeatedly since its first classification in 1986, most recently in 2021, when the upper limit of normal hearing was lowered from 25 to 20 dB HL and the threshold for disabling hearing loss from 40 to 35 dB HL^10,11^. That single 5 dB step nearly doubled the number of children identified in a school screening programme^25,27^.

The field treated this fragmentation and instability as a problem of method, of finding the right algorithm, cohort or number of groups^16,17,26^. The more fundamental possibility, that the categorical premise itself was wrong, went untested for 140 years^1–8,12–14,16,17,19,21–24,26,28–34^.

We tested the premise and falsified it. Human audiograms do not resolve into a reproducible catalogue of types. They occupy a universal, continuous space of four to five dimensions. We call it the Human Hearing Atlas (HHA; Fig. 1).

**Figure 1.**
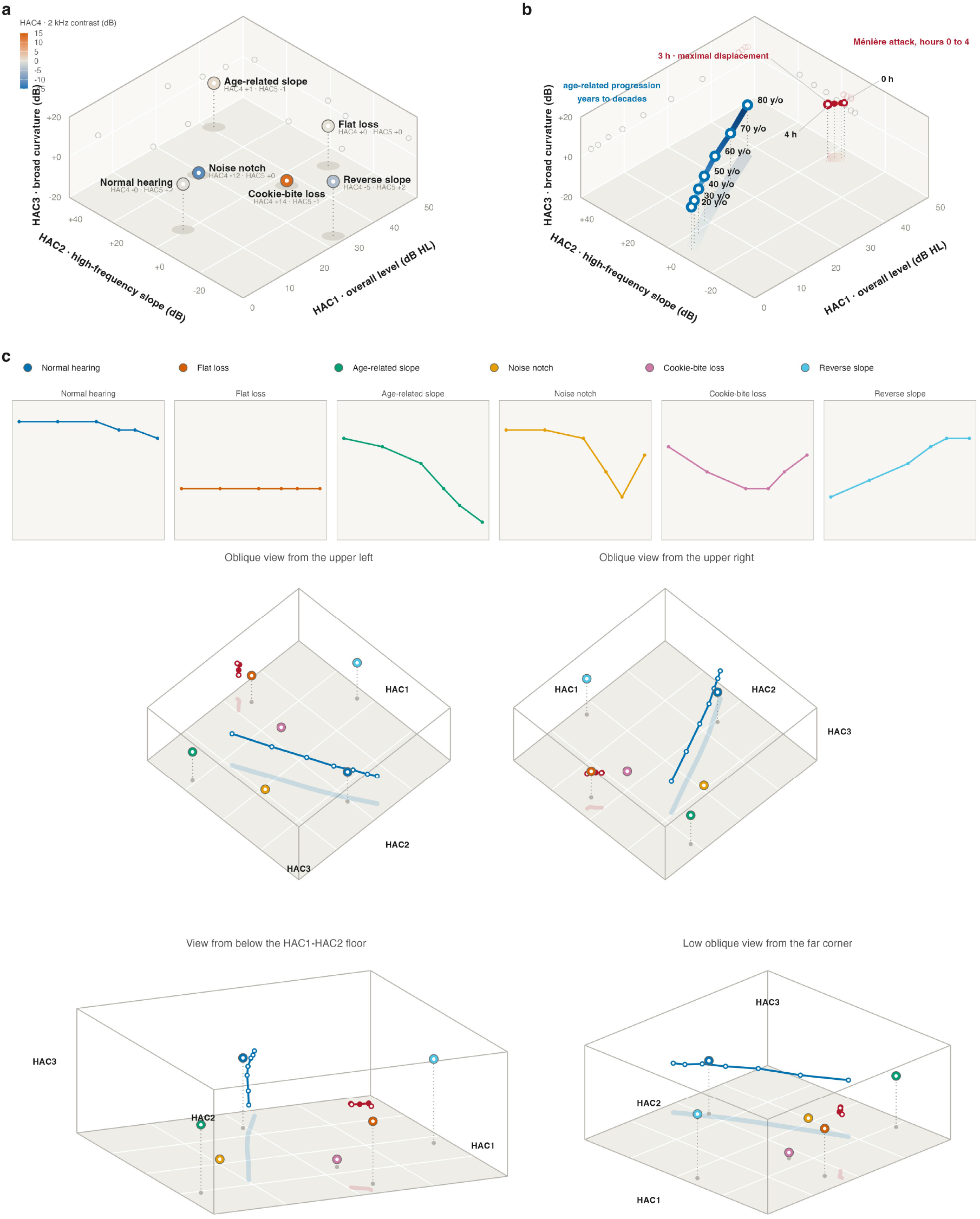
| The Atlas coordinate space of human hearing.(a) Six illustrative audiometric configurations projected with the frozen basis into the first three Atlas coordinates: position encodes HAC1 (overall level), HAC2 (high-frequency slope) and HAC3 (broad curvature); marker fill encodes HAC4 (2 kHz contrast); the printed pair beneath each label gives HAC4 and HAC5 (3 kHz contrast) exactly. Grey ellipses mark floor positions and open circles the wall projections.(b) Hearing change as movement through the same room. Chronic age-related progression over decades (blue, schematic) and the measured acute Ménière trajectory over hours 0 to 4 (crimson, group means over 356 ear-series at hours 0 to 3 and 354 at hour 4). Colour runs with elapsed time along each band; each trajectory casts a floor shadow in its own hue; dotted stems drop each waypoint to the HAC1-HAC2 floor.(c) Top: the six configurations, colour-coded (Wong palette; Reverse slope nudged toward cyan for discriminability), with each configuration’s audiogram (0.5 to 6 kHz, 0 to 70 dB HL). Below: the same room from four orthographic viewpoints (upper-left and upper-right obliques, from below the HAC1-HAC2 floor, and a low oblique from the far corner). Both trajectories and their floor shadows appear in every view; dotted stems anchor each configuration to the floor. Coordinates computed from the frozen basis; HAC4 and HAC5 are not represented in the four views.

We asked whether, in people no model had seen, hearing sorts into kinds or flows along a continuum, and required of any category that it recur when the sample, the frequencies and the era change^35–37^.

The result was the same in every large adult population, nation and era: the continuous model provided superior fit. Discrete groupings surfaced within individual cohorts and age strata, yet their number and occurrence were cohort-dependent, and when those populations were pooled the structure dissolved. The subtype counts reported since 1932 emerge from two artifacts of measurement and sampling identified here: 5 dB quantisation, in which the audiometer’s steps snap different ears onto identical tracings; and cohort pooling, in which combining generations blurs each cohort’s local structure.

We assembled a cross-population and cross-era corpus of more than 4.29 million audiograms from 33 datasets spanning eight countries, 62 years and two calibration standards. The Atlas was derived, then frozen and evaluated without refitting in independent datasets from six countries and across multiple frequency grids. To test whether the derived Atlas was merely a cohort-specific fit, we tested independent recovery and transportability: unrelated datasets from five countries independently recovered Atlas axes without access to the canonical coordinates, rank or labels, with every shape axis recurring across multiple cohort families; independently derived frozen coordinate systems also transported across populations without refitting.

Across the populations, generations, instruments, standards and measurement routes examined, four to five independent dimensions recur. We fix this structure in five Hearing Atlas Coordinates (HAC): HAC1, overall hearing level; HAC2, high-frequency slope; HAC3, broad curvature; HAC4, local 2-kHz contrast; and HAC5, local 3-kHz contrast (Figs. 1a and 2). Five coordinates locate any audiogram on a supported grid and, in people who did not contribute to the derivation, reconstruct it to 1.35 dB. The audiogram itself does not need replacing: it contains the information and always has.

**Figure 2.**
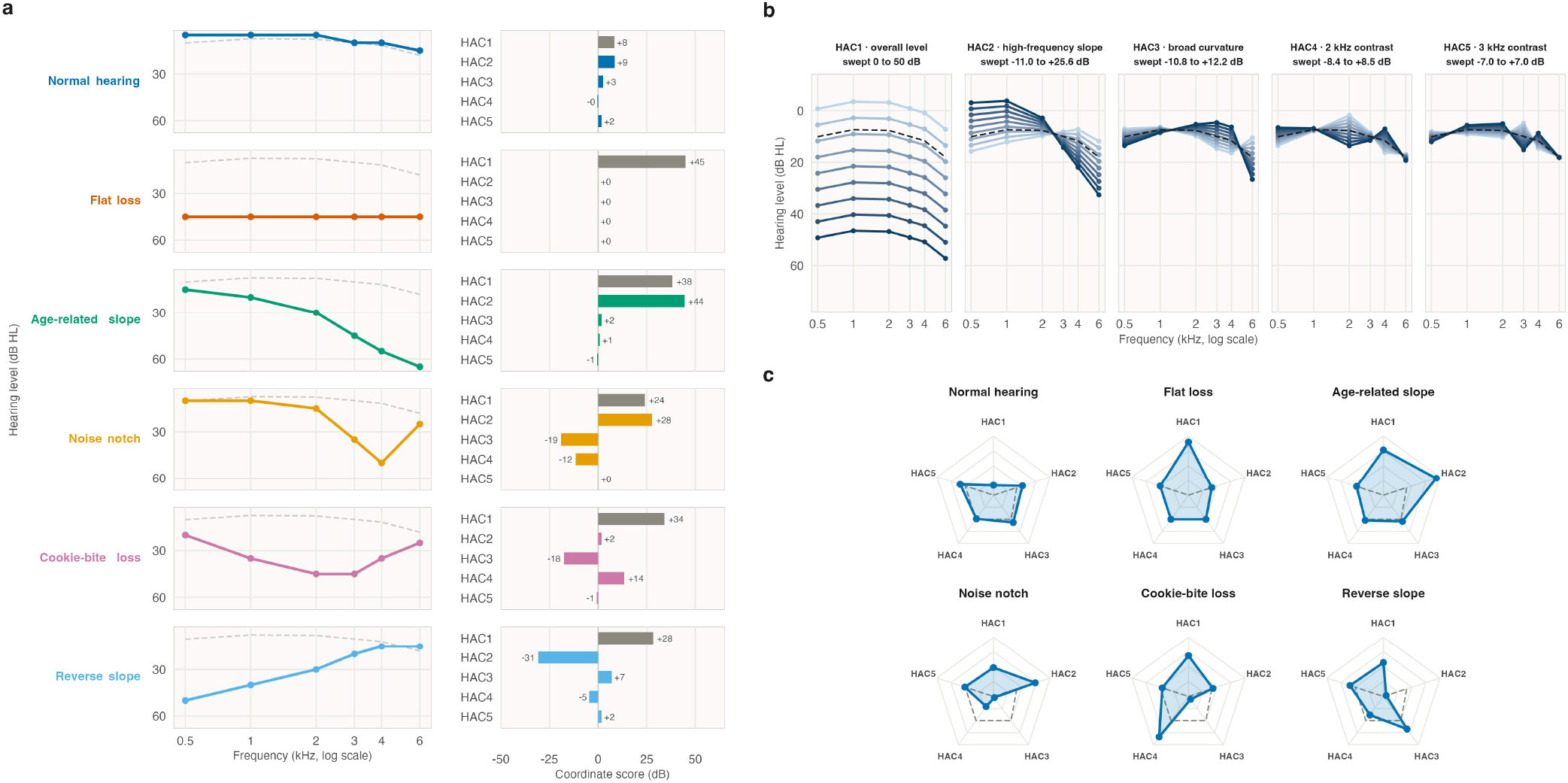
| Classic audiometric configurations and their Atlas coordinates.(a) Six illustrative audiometric configurations as six-frequency audiograms (left, dashed grey is the population median) and as their five frozen Human Hearing Atlas coordinates (right). HAC1 is the overall-level coordinate and HAC2-HAC5 are signed shape coordinates.(b) Each coordinate swept independently about the population median audiogram (dashed), with all other coordinates held at their median values. HAC1 shifts overall level without changing shape, swept 0 to 50 dB, an illustrative range; HAC2 to HAC5 are the orthonormal shape coordinates, each swept across the population 10th to 90th percentile range. Curve shading encodes the swept score, light at the lower bound and dark at the upper bound.(c) The same six configurations as pentagons over the five coordinates. Spokes are scaled to display ranges (HAC1 0 to 50 dB, HAC2 -35 to +50, HAC3 and HAC4 -20 to +20, HAC5 -10 to +10); the dashed pentagon marks zero on every shape coordinate, so lobes outside it are positive scores and dents inside it negative. Flat loss is the pure zero-shape pentagon at high HAC1; the noise notch throws opposite lobes on HAC2 and HAC3.The named configurations are familiar clinical landmarks within a continuous coordinate system rather than mutually exclusive biological patient types.

This breakthrough discovery has six foundational transformations for hearing science and otology, with major implications for medicine and science more broadly. First, it settles the 140-year-old belief that human hearing is organised into discrete categories: human hearing is a continuum^3,5,6^. Second, it resolves the field’s foundational crisis of fragmentation and instability by showing that methodological artefacts and a categorical premise caused classifications to proliferate and fail to replicate. Third, it demonstrates and quantifies how much information is lost by the PTA, and how conventional configuration labels divide a continuous hearing space into arbitrary regions^29,32,34,38^ (Fig. 2). Fourth, it unifies the field through the HHA, a universal coordinate system reproducible through independent recovery and generalizable across populations, generations, calibration standards and measurement routes through zero-shot transfer (Fig. 3d). Fifth, it formulates and empirically demonstrates a new physiological law, the Law of Human Hearing States (LHHS), characterized by four empirical invariants: continuity, low dimensionality, universality, and state-motion correspondence (Fig. 3 and Fig. 4). Sixth, it constitutes a paradigm shift in how science and medicine describe and investigate the human sense of hearing, opening new directions for precision otology, audiology, hearing rehabilitation, cochlear implantation, genetics, epidemiology, regenerative medicine and clinical trials.

**Figure 3.**
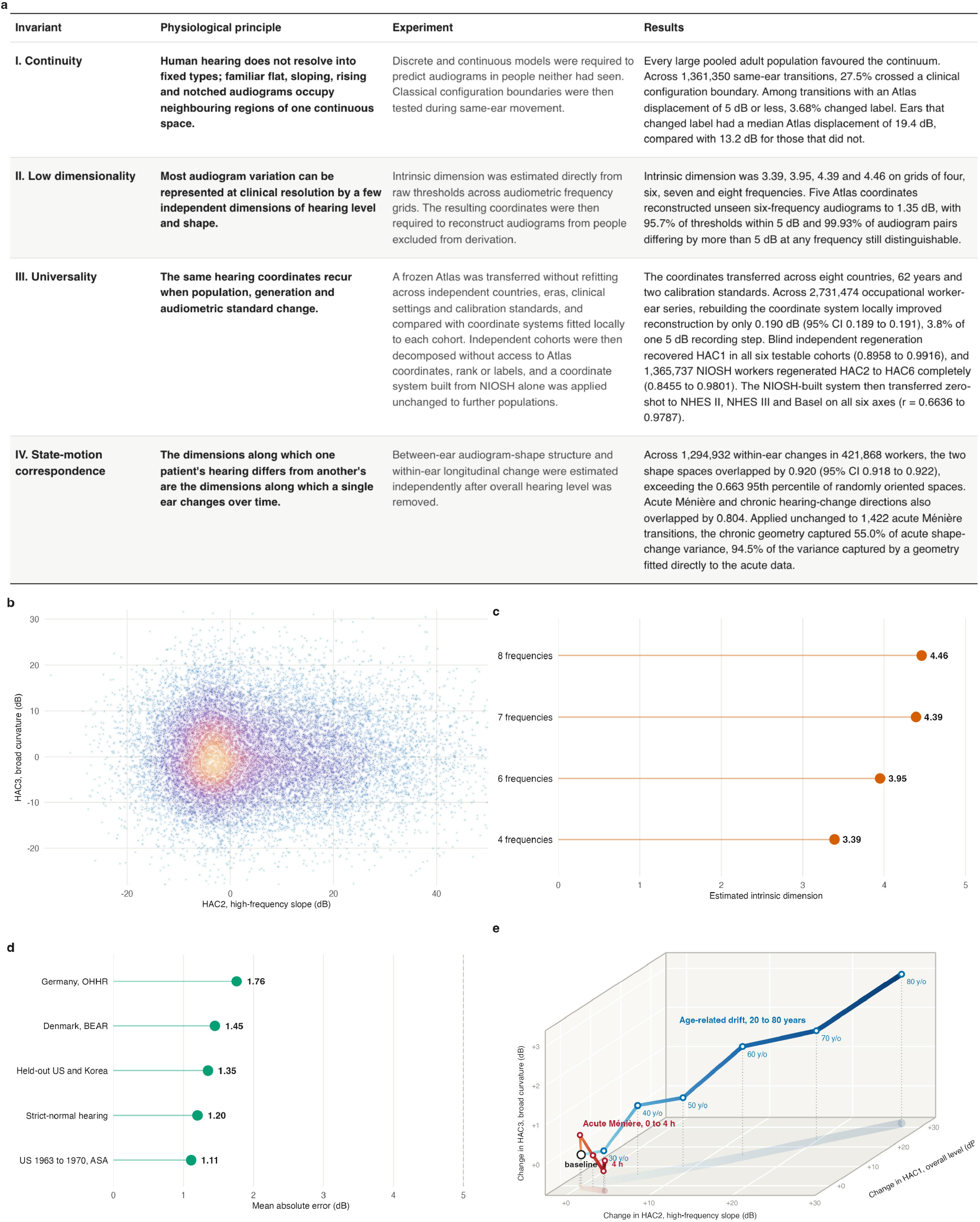
| The Law of Human Hearing States.(a) For each of the four invariants: the physiological principle, the experiment that tested it, and the result.(b) Conventional configurations occupy one continuous space. A schematic population of 16,000 ears in HAC2 (high-frequency slope) and HAC3 (broad curvature), coloured by local kernel density; white rings are isodensity contours. Across 1,361,350 same-ear transitions, 27.5% crossed a conventional label boundary; only 3.68% of transitions within 5 dB changed label.(c) Dimensionality converges at four to five. Intrinsic dimension rose from 3.39 on four-frequency grids to 4.46 on eight-frequency grids; five coordinates reconstructed held-out thresholds to 1.35 dB.(d) The frozen frame transfers across independent datasets. Out-of-sample error stayed below 2 dB in every cohort; local refitting across 2,731,474 occupational ear-examinations improved error by only 0.190 dB (95% CI 0.189 to 0.191). Recurrent independent recovery of the coordinate axes and zero-shot transport are detailed in Supplementary Figure S3.(e) Within-ear and between-ear variation share the same shape dimensions. Chronic age-related drift (blue, schematic, 20 to 80 years) and the measured acute Ménière trajectory (crimson, group means over 356 ear-series at hours 0 to 3 and 354 at hour 4), each from its own baseline. Across 1,294,932 within-ear changes in 421,868 workers, between-ear and within-ear shape spaces overlapped by 0.920 (95% CI 0.918 to 0.922) against a random-orientation null whose 95th percentile was 0.663 (Monte Carlo P = 0.0005); acute and chronic directions overlapped by 0.804 (null median 0.409, 95th percentile 0.671; Monte Carlo P = 0.0105). Applied without refitting to 1,422 acute Ménière successive transitions, the frozen chronic NIOSH rank-2 frame captured 55.02% of acute shape-change variance against a 58.20% locally fitted acute ceiling, a transfer efficiency of 94.53%, and reconstructed shape-change at 1.793 dB mean absolute error against 1.790 dB for the locally fitted frame (random rank-2 null P = 0.0015 for variance captured and P = 0.0010 for error).

**Figure 4.**
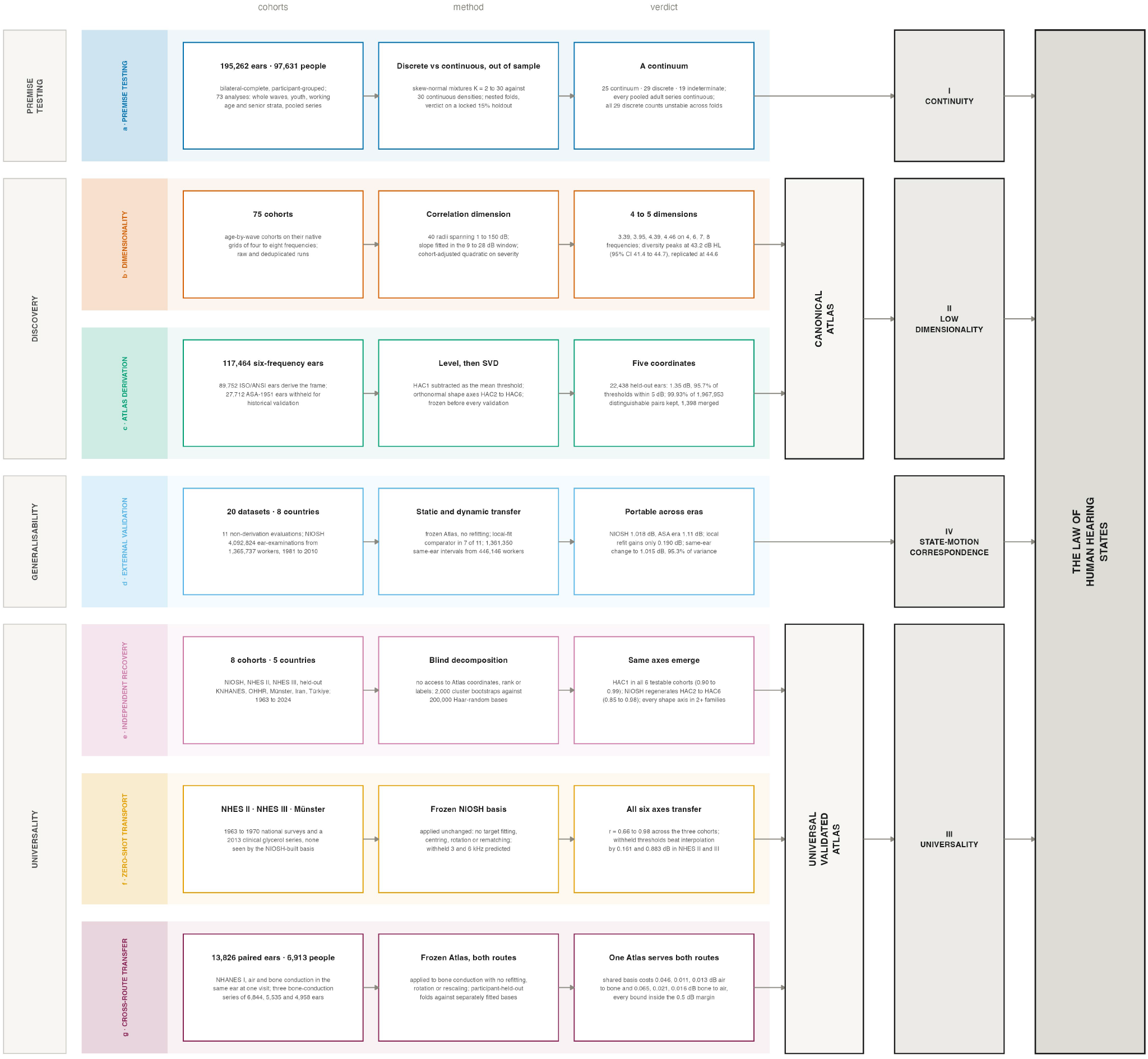
| Study design and analytical validation of the Human Hearing Atlas.The study comprised seven analytical pipelines spanning premise testing, dimensionality estimation, Atlas derivation, external validation, independent coordinate recovery, zero-shot transport and cross-route transfer. Together, these analyses tested four empirical properties of human audiometric variation: continuity, low dimensionality, universality and correspondence between cross-sectional and longitudinal structure. Rows are the seven pipelines, grouped by phase (left); columns give the cohorts analysed, the method applied and the result.(a) Premise testing. Across 195,262 ears in 73 participant-grouped analyses, discrete models exhibited fold instability, whereas every large pooled adult series favoured a continuum.(b) Dimensionality. Intrinsic dimension reaches 4.46 on eight-frequency grids and peaks in moderate hearing loss, at 43.2 dB HL.(c) Atlas derivation. The coordinate frame was derived from 89,752 ears by singular-value decomposition after extraction of overall hearing level. Five coordinates reconstruct held-out thresholds to 1.35 dB, preserving 99.93% of clinically distinguishable pairs.(d) External validation. The frozen Atlas was evaluated without refitting on independent datasets. Static error across 4,092,824 occupational ear-examinations was 1.018 dB; dynamic error across 1,361,350 longitudinal same-ear changes was 1.015 dB.(e) Independent recovery. Independent decomposition of eight external cohorts recovered the canonical axes without access to Atlas coordinates, rank or labels.(f) Zero-shot transport. The independently recovered frame transports across datasets without target fitting, predicting jointly withheld 3 and 6 kHz thresholds better than interpolation.(g) Cross-route transfer. Universality was tested across measurement route as well as across population and era. In 13,826 paired ear-records from 6,913 participants of the first US National Health and Nutrition Examination Survey, air and bone conduction were measured in the same ears at one visit, yielding three bone-conduction series of 6,844, 5,535 and 4,958 ears. The canonical Atlas was applied to bone conduction without refitting, rotation or rescaling. Against models permitted to fit separate air- and bone-conduction representations, the shared Atlas cost 0.046, 0.011 and 0.013 dB reconstructing bone conduction from air conduction, and 0.065, 0.021 and 0.016 dB in the reverse direction, every upper confidence bound falling inside the prespecified 0.5 dB noninferiority margin.

## Results

### Audiograms do not form universal types

Every large pooled adult series favoured the continuous model: in the Korea National Health and Nutrition Examination Survey (KNHANES) 2019-2023, KNHANES 2009-2012, the US National Health and Nutrition Examination Survey (NHANES) 1999-2020 and US NHANES 1971-1974 the continuous model predicted unseen audiograms better by wide margins, as did the pooled four-country analysis (Supplementary Figure S1a and Supplementary Table S1) of 170,028 ISO/ANSI-referenced ears.

### The 5 dB measurement grid creates artificial audiogram clusters

Clinical 5 dB recording concentrated identical audiograms: 41.0% of Korean, 51.7% of US post-1999 and 69.5% of US pre-1999 ears shared their complete six-frequency pattern with another ear, rising to 96.2% across 4,092,824 occupational ear-examinations, which collapsed onto 353,408 distinct patterns, the largest holding 25,083 ears (Supplementary Figure S1b). We term this the 5 dB quantisation artifact.

Treating thresholds as 5 dB intervals shifted eighteen cohort analyses from continuous to discrete and none the other way; out of sample, 20 discrete classifications disappeared and none became discrete (Supplementary Table S1g).

In all 29 discrete-verdict analyses the selected component count varied across outer folds, with 1,569 of 2,884 evaluable fold fits (54.4%) selecting a count different from the final reported count (Supplementary Figure S1d) (Supplementary Table S1e and Supplementary Methods S2).

### The current conventional audiogram labels occupy overlapping regions of the Atlas

Conventional configurations (including normal, flat, age-related sloping, noise-notched, cookie-bite and reverse-sloping audiograms) could each be expressed as a position in the same Atlas coordinate system (Fig. 2). Across the full corpus, these configurations did not partition the Atlas into separate types (Fig. 5). They occupied heavily overlapping, contiguous regions of the same coordinate space.

**Figure 5.**
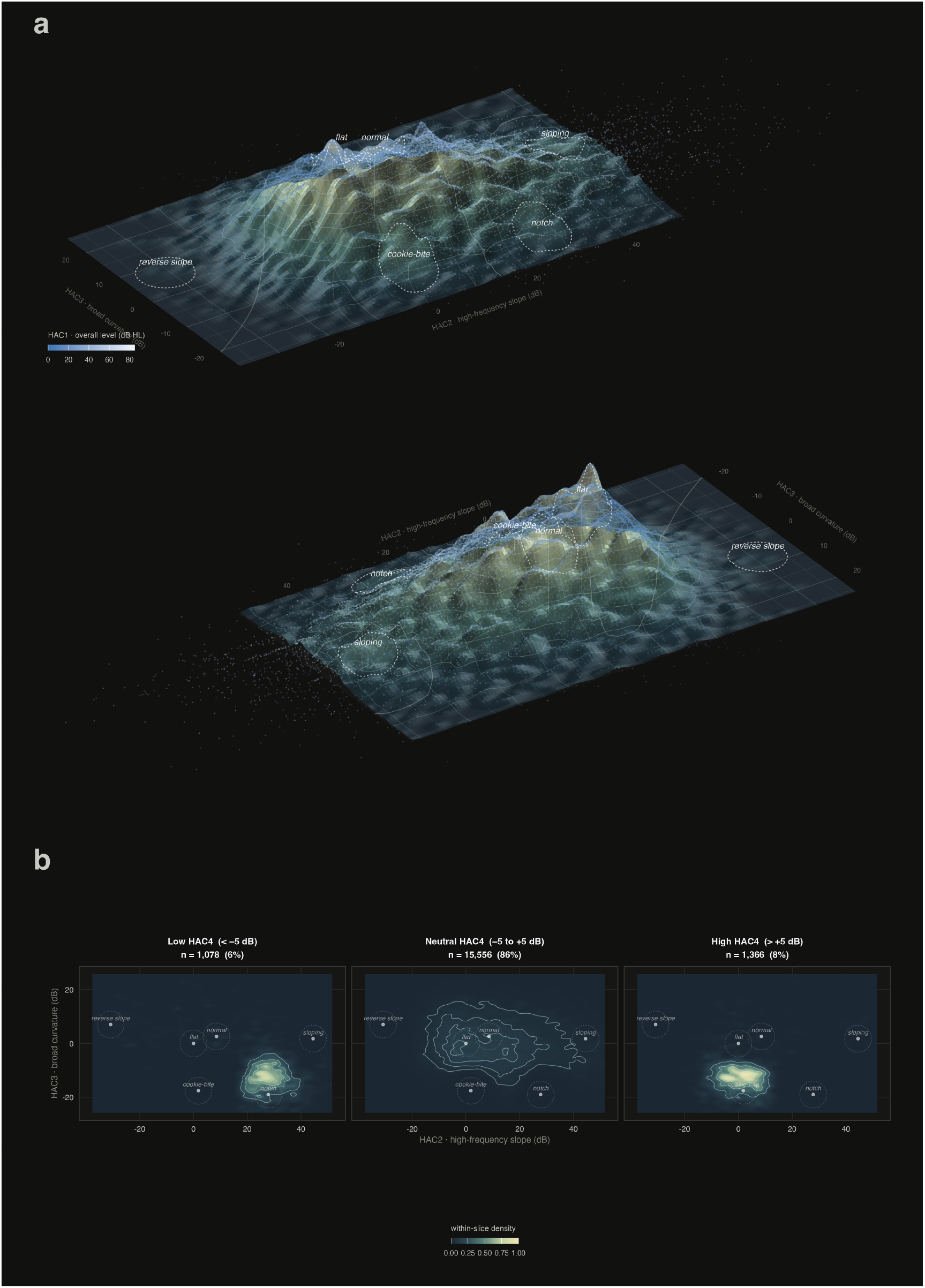
| Continuous topography of the Human Hearing Atlas and the location of classical audiometric configurations.(a) A schematic population of 18,000 ears in the two dominant shape coordinates, lifted by the density of the displayed cloud. HAC2 is high-frequency slope; HAC3 is broad curvature; vertical relief is the kernel-density estimate of the displayed cloud, not an Atlas coordinate. Two orthographic views, 180 degrees apart, of the same landscape. Brightness rises with overall level (HAC1). Conventional audiogram configurations are 5-unit Atlas neighbourhoods, draped and shaded through the same three-dimensional geometry as the coordinate fabric. Faint ivory isolines are local 2 kHz contrast (HAC4), interpolated from the textbook configurations and draped on the landscape (solid positive, dashed negative; HAC4 = 0 is the emphasized contour).(b) The same schematic population and HAC2-HAC3 window, split by HAC4: low (below -5 dB), neutral (-5 to +5 dB) and high (above +5 dB). Colour is within-slice density on a shared scale, so the broader neutral panel is less intense than the concentrated tails. Textbook centres are fixed. HAC4 redistributes occupancy inside one continuous geography; it does not invent separate hearing types. Slice sizes are printed on each panel.

Across 1,361,350 successive same-ear comparisons, 27.5% crossed at least one conventional label boundary, labels being joint severity-by-configuration classes, and 59.9% of label changes were shape-label changes at unchanged severity grade. Among the 37,715 comparisons with an isometric Atlas displacement of 5 dB or less, 3.68% changed label (Fig. 3b).

### Human hearing varies along few continuous dimensions

Across 75 cohorts, dimensionality estimated directly in native threshold space increased as additional frequencies exposed previously unobserved audiogram structure: 3.39 on four-frequency audiograms, 3.95 on six, 4.39 on seven and 4.46 on eight (Fig. 3c).

### Five coordinates preserve audiograms

Five coordinates separate overall hearing level from four dimensions of audiogram shape: HAC1, overall hearing level; HAC2, high-frequency slope; HAC3, broad curvature; HAC4, local 2 kHz contrast; and HAC5, local 3 kHz contrast (Figs. 1a and 2a, b). HAC6, a local 1 kHz contrast, completes the six-frequency representation (Supplementary Table S2).

In 22,438 ears from 11,219 participants who contributed nothing to the derivation, five coordinates reconstructed thresholds to 1.35 dB, placing 95.7% of thresholds and every measured frequency of 81.4% of ears within 5 dB (Fig. 6b, c). Of 1,967,953 audiogram pairs differing by more than 5 dB at any frequency, 99.93% remained distinguishable and 1,398 merged (Supplementary Table S2).

**Figure 6.**
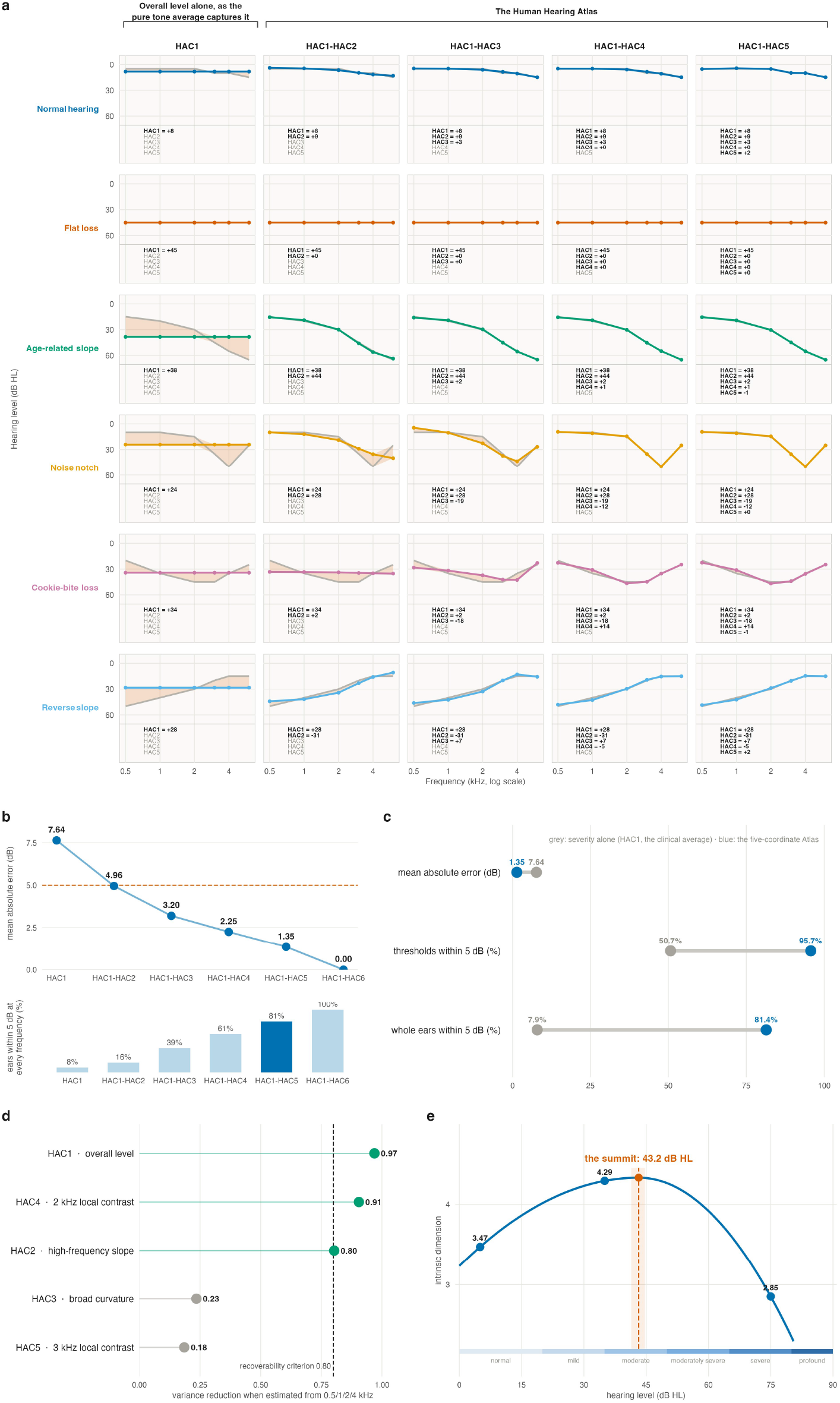
| Six classic audiometric configurations, rebuilt one coordinate at a time.(a) Rows are the six illustrative configurations of Figure 2, measured at 0.5, 1, 2, 3, 4 and 6 kHz. Columns add coordinates in order, HAC1 (overall level) through HAC5 (3 kHz local contrast). Grey, the measured audiogram; colour, the reconstruction at that depth; the vermillion wash is the residual. Included coordinates are printed in black with their value, the rest in grey. (b) Held-out reconstruction error by number of coordinates, in 22,438 unseen ears from 11,219 held-out participants: 7.64, 4.96, 3.20, 2.25 and 1.35 dB at depths one to five.(c) What the four shape coordinates buy, on the same held-out ears. Overall level alone (grey, HAC1) against the full five-coordinate frame (blue): mean absolute error falls from 7.64 to 1.35 dB, thresholds within 5 dB rise from 50.7% to 95.7%, and whole ears within a clinical step at every frequency rise from 7.9% to 81.4%.(d) What a four-frequency screen still carries. Proportional reduction in the variance of each coordinate when estimated from 0.5, 1, 2 and 4 kHz alone, United States cohort. Level (0.97), 2 kHz local contrast (0.90) and high-frequency slope (0.80) meet the prespecified 0.80 criterion (green); broad curvature (0.24) and 3 kHz local contrast (0.19) do not (grey). (e) Audiometric diversity peaks in moderate loss. Median intrinsic dimension peaked at a fitted summit of 43.2 dB HL (95% CI 41.4 to 44.7). Bands are the WHO grades.

### Moderate hearing loss contains the greatest diversity

Across severity bands, median dimensionality rose from 3.47 at 0-10 dB to 4.29 at 30-40 dB, then declined to 2.85 at 70-80 dB. A cohort-adjusted quadratic model placed the maximum at 43.2 dB (95% CI 41.4-44.7), and an independent pooled external analysis of 24 cohorts and 104 cohort-band cells placed it at 44.6 dB (95% CI 43.1-46.1). Ears within a 2 dB window at 43.2 dB differed by up to 124.5 dB summed across six frequencies rather than at any single frequency (Fig. 6e).

### Normal and impaired hearing are not independent categories, they lie on a continuous spectrum

In ears at 20 dB HL or better at all six frequencies, the unchanged Atlas reconstructed thresholds to 1.20 dB (95% CI 1.18-1.22), with 97.1% of thresholds and all six frequencies of 85.9% of ears within 5 dB. Representation did not change abruptly at the clinical boundary: estimated discontinuities at 15, 20 and 25 dB HL were all below the prespecified 0.5 dB limit. Normal hearing nevertheless retained substantial multidimensional shape variation, although HAC2-HAC5 captured it less completely than in impaired hearing; the prespecified equivalence criterion, and consequently the full-spectrum composite criterion, were therefore not met (Supplementary Methods S10).

### The Atlas transfers across populations

Coordinates derived independently from the ISO/ANSI US waves and the Korean surveys were closely congruent, with axis agreement of 0.954 to 0.998 (Supplementary Methods S8). After conversion to the ANSI-1969 reference, the Atlas reconstructed 27,712 ASA-1951 ears to 1.1088 dB, with 97.5-97.7% of thresholds within 5 dB (Supplementary Table S3).

Across the 20 external-validation datasets from all eight countries (Fig. 7), the canonical Atlas was applied without refitting in all 11 non-derivation evaluations (Supplementary Table S3 and Fig. 3d and Supplementary Figure S2). Seven supported a locally fitted comparator; in four of seven the universal coordinates reconstructed better than the local fit.

**Figure 7.**
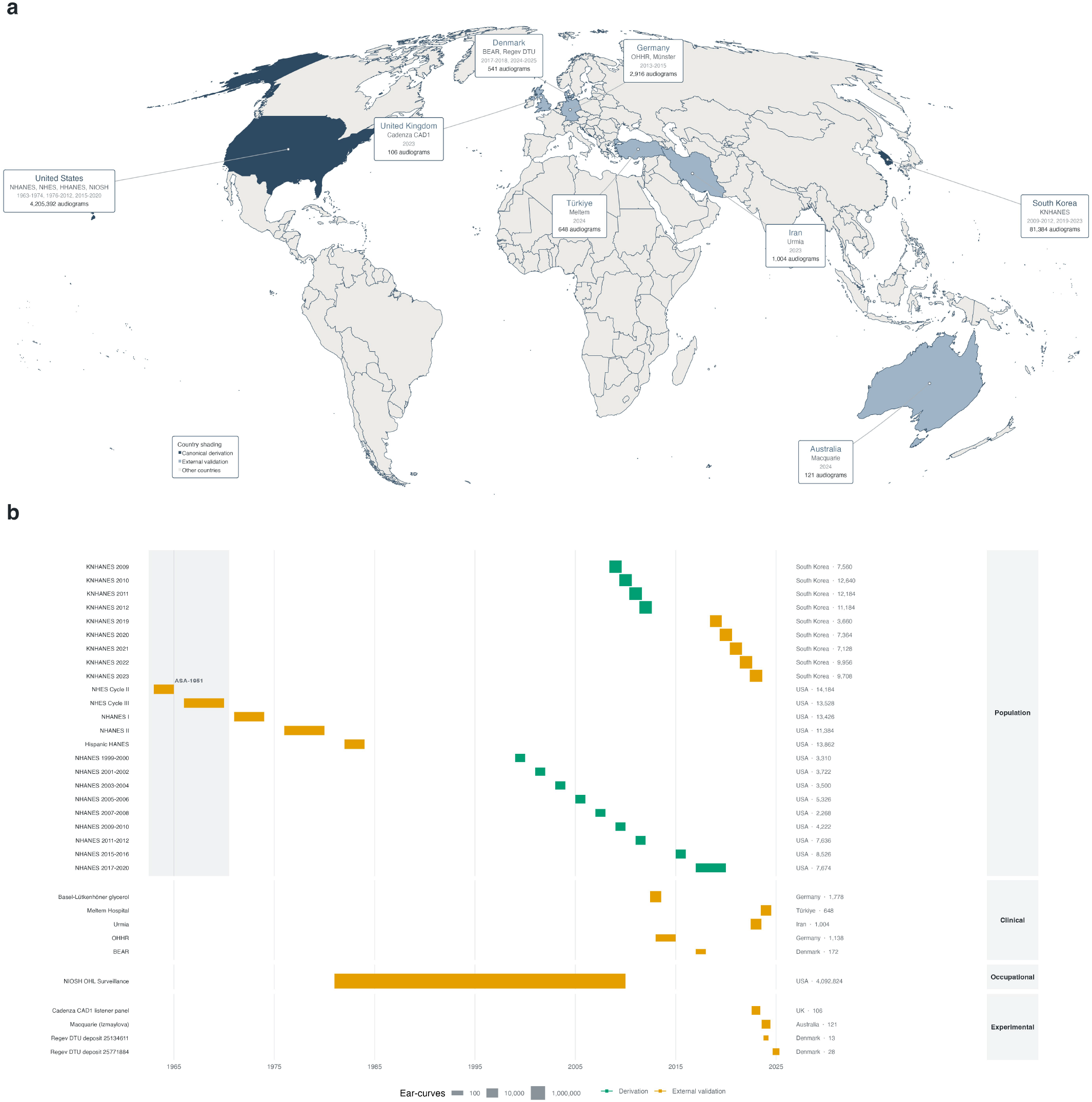
| Geographical distribution and scale of the derivation and external-validation cohorts. (a) Countries contributing audiograms. Dark shading marks the canonical derivation countries and light shading the external-validation countries; countries without study data are unshaded. Each card gives the country, its datasets, the calendar spans they cover and the aggregate record count.(b) Every dataset in the study on a calendar timeline from 1963 to 2025, grouped by cohort type. Bars span multi-year collections and squares mark single-year datasets; colour gives the role of the dataset in the Atlas, derivation or external validation. Bar thickness and marker size encode post-QC record count on a log10 scale, which is required because the datasets span five orders of magnitude, from occupational surveillance to small experimental deposits. Right-hand values give the country and that count in the unit each dataset reports: ears for most, curated ear-examination observations for the occupational archive, ear-series for the glycerol series and participants for the two smallest deposits. Counts in different units are not summed. The shaded band marks the ASA-1951 era, before thresholds were converted frequency-by-frequency to ANSI-1969 at decoding.

Applied without refitting to 4,092,824 ear-examinations from 1,365,737 US workers recorded 1981 to 2010, five coordinates reconstructed thresholds to 1.018 dB, 98.19% within 5 dB. Deriving a new coordinate system from this dataset improved reconstruction by 0.1900 dB, 3.8% of one 5 dB step (Supplementary Table S3).

### Air and bone conduction can use the same Atlas

We used 13,826 paired ear-records from 6,913 participants of the first US National Health and Nutrition Examination Survey (NHANES), in whom air conduction and bone conduction were measured in the same ears at one visit. The canonical Atlas was applied to bone conduction without refitting, rotation or rescaling. Air and bone conduction preserved the same patient-specific hearing pattern, supporting their representation as two measurement routes to the same underlying hearing state. This relationship was not driven by hearing-loss severity: the same-ear pattern remained recognizable after overall hearing level was removed and was also present among ears whose measured air-conduction thresholds were all normal. The relationship persisted across every stratum of side, age and hearing level. After accounting for ties caused by 5-dB quantisation, the correct cross-route partner fell at mean percentiles of 0.251, 0.287 and 0.308, compared with approximately 0.50 expected by chance (Supplementary Methods S25). Bone conduction therefore did not require a separate hearing representation: the same Human Hearing Atlas could represent both routes.

### Air and bone conduction reconstruct each other in both directions

The relationship between air and bone conduction was bidirectional: air conduction reconstructed bone conduction, and bone conduction reconstructed air conduction. One Atlas was sufficient in both directions. Compared with models allowed to fit separate air- and bone-conduction representations, the shared Atlas remained well within the prespecified 0.5 dB noninferiority margin in every series. For four-frequency reconstruction including overall hearing level, the shared Atlas differed from route-specific representations by 0.046, 0.011 and 0.013 dB for air-to-bone reconstruction and by 0.065, 0.021 and 0.016 dB for bone-to-air reconstruction. The frozen Atlas was also no worse than a representation fitted within the cohort itself, and performed better in one of the three series (Supplementary Methods S25).

### Independent populations reconstruct and transport the Atlas coordinates without refitting

To test whether the Atlas axes were reproducible rather than a particular fit to the 89,752-ear US and South Korean derivation frame, we independently decomposed the National Institute for Occupational Safety and Health (NIOSH) archive, National Health Examination Survey (NHES) Cycles II and III, held-out KNHANES waves, the Oldenburg Hearing Health Record (OHHR), Münster, Iran and Türkiye without access to Atlas coordinates, rank or labels. HAC1 emerged in all six testable cohorts (0.8958 to 0.9916), and 1,365,737 NIOSH workers regenerated HAC2 to HAC6 completely (0.8455 to 0.9801), with every shape axis replicated across at least two cohort families (Supplementary Figure S3a). We then froze this NIOSH-built coordinate system and applied it unchanged to NHES II, NHES III and Münster. All six axes transferred unchanged without refitting (r = 0.6636 to 0.9787; Supplementary Figure S3b), and predicted jointly withheld 3 and 6 kHz thresholds better than interpolation in both NHES waves (mean absolute error reductions 0.161 and 0.883 dB; Supplementary Figure S3c, Supplementary Table S4 and Supplementary Methods S24).

The prespecified five-part conjunctive recovery criterion was not met (Supplementary Methods S24); axis-level recovery and zero-shot transport were therefore evaluated separately.

### Previous described dimensions were independently replicated

On a 0.25, 0.5, 1, 2, 4 and 8 kHz grid, Bamford et al. (201 ears) and Bench (391 ears) identified overall hearing level and slope, accounting for 85.72% and 87% of variation, and each dismissed a third dimension of 8.21% and 6% as noise^22,23^.

To reproduce their limitation experimentally, we restricted 11,084 held-out ears from the US NHES II and III surveys (1963-1970) to the Bamford-Bench frequency grid while keeping the canonical Atlas frozen. Variance reduction was 0.933 for HAC1, 0.666 for HAC2, 0.296 for HAC3, 0.894 for HAC4 and 0.282 for HAC5; HAC3 and HAC5 therefore did not meet the prespecified 0.80 recoverability criterion. The loss replicated in 1,638,338 held-out NIOSH ear-examinations after joint removal of 3 and 6 kHz, with variance reduction of 0.445 for HAC3 and 0.090 for HAC5 (Supplementary Table S3d-e).

### Missing frequencies remove information

Masking the six-frequency audiogram down to 0.5, 1, 2 and 4 kHz showed which coordinates a four-frequency audiogram still carries. Overall level, high-frequency slope and the local 2 kHz contrast were retained; broad curvature and the local 3 kHz contrast were not (Supplementary Table S3b and Fig. 6d), and reconstructed thresholds at the two hidden frequencies carried mean absolute errors of 5.09 dB at 3 kHz and 8.26 dB at 6 kHz.

### The Atlas partially predicts missing frequencies

Compared with log-frequency interpolation between neighbouring measured thresholds, in participants held out from fitting, the Atlas predicted 8 kHz better in both large cohorts, improving explained variance by 1.98 percentage points across 2,046,160 occupational examinations and by 1.53 points in US NHANES 2011-2012. At the interior frequency 1.5 kHz the two were indistinguishable; below 0.5 kHz the Atlas was worse than simple extrapolation, losing 3.0 and 2.4 points at 125 and 250 Hz.

### Five coordinates reconstruct longitudinal change to 1.0 dB

Across 1,361,350 successive same-ear changes from 446,146 workers, five HAC reconstructed the observed change in the six measured thresholds with a mean absolute error of 1.015 dB and accounted for 95.32% of the observed variation in threshold change, against 4.28 dB and 39.6% of variance from overall hearing level alone, a fourfold reduction (Supplementary Figure S4a and Supplementary Table S5).

### The Audiogram’s shape dominates longitudinal change

Overall hearing level accounted for 42.84% of total movement through the Atlas (95% CI 42.6-43.0%), whereas changes in audiogram shape accounted for 57.16% of movement (Supplementary Figure S4b and Supplementary Table S5b). The greatest diversity of change occurred in ears beginning between 40 and 49 dB, with a median baseline hearing level of 43.3 dB, closely coinciding with the intermediate-severity region in which cross-sectional audiogram diversity was greatest (Supplementary Figure S4d).

### Human hearing change has no persistent direction

The starting audiogram accounted for 4.9% of subsequent hearing change, exceeding chance expectation (permutation P = 0.001; Supplementary Methods S16). Across repeated examinations, however, changes in audiogram shape did not continue consistently in the same direction. Instead, a change observed during one interval tended to be partly reversed during the next, even after changes in overall hearing level were removed (mean cosine -0.30; Supplementary Methods S20).

### The Human hearing atlas dimensions track change within the same person, and between persons

Across 1,294,932 longitudinal changes from 421,868 workers, the patterns of slope, curvature and local frequency differences that distinguished different ears showed 0.920 overlap with the patterns along which the same ear changed (95% CI 0.918-0.922), against a random-orientation null with median 0.403 and 95th percentile 0.663 (P = 0.0005; Fig. 3e and Supplementary Figure S4e, and Supplementary Table S5b).

Scrambling the order of examinations within each worker preserved the relationship.

### Acute Ménière hourly change traces a trajectory in the same hearing space as multi-decade hearing loss change

In 356 unilateral-ear Ménière series followed across five successive timepoints, each acute episode was represented as movement through the same Atlas (Fig. 1b). The group trajectory involved simultaneous changes in overall hearing level and high-frequency slope, rather than a uniform shift of the whole audiogram.

After overall hearing level was removed, the principal patterns of audiogram-shape change during the Ménière series showed 0.804 overlap with those observed during chronic same-ear change, substantially more than expected between randomly oriented hearing patterns (null median 0.409, 95th percentile 0.671; P = 0.0105; Fig. 3e and Supplementary Figure S4e).

Series-level resampling gave a 95% CI of 0.736 to 0.859. Applied without refitting to 1,422 hourly transitions, the frozen chronic geometry reached 94.53% of the locally achievable variance capture (Supplementary Methods S18).

## Discussion

For 140 years, human hearing has been interpreted through discrete types and severity grades^3,5–8^. Here, every large pooled adult population favoured a continuous model. Human audiograms therefore do not form a reproducible catalogue of kinds; they occupy a continuous, low-dimensional space. The HHA provides coordinates for that space (Fig. 1), separating overall hearing level from slope, curvature and local frequency contrasts, and reconstructing unseen six-frequency audiograms to 1.35 dB while preserving 99.93% of clinically distinguishable pairs (Fig. 6b, c). Earlier studies identified low-dimensional audiometric structure or a continuum in presbycusis^22–24^, but neither established continuity across populations, eras and clinical states. Our results do so while preserving an important distinction: cohort-specific density clusters can produce recurrent local groupings, but they do not constitute universal human hearing types.

This distinction explains why audiogram taxonomies proliferated yet failed to replicate in the digital and AI era. Clinical audiometers record thresholds in 5 dB steps^39–41^, concentrating different underlying ears onto identical recorded vectors, so that clustering such lattice points converts instrumental resolution into apparent phenotypes^16,24,35,43^. This is the grouped-data problem described mathematically by Sheppard in 1898, 34 years before Guild’s systematic classification of audiogram curves in 1932^44^. We observed this directly: in all 29 analyses favouring discreteness the selected count varied across folds. This empirical instability accords with theoretical results showing that, under model misspecification, finite-mixture component counts need not converge to a stable underlying class count^35^. A second artifact arises from cohort composition: age, exposure and recruitment generate local density concentrations, whereas pooling populations and generations dissolves groupings that do not recur (Supplementary Figure S1c, d)^17,18^. A third source is the widespread practice of treating highly correlated left and right ears as independent statistical observations, which causes algorithms to mistake paired within-patient measurements for local density concentrations^14,16,17^. The three-to-29 groups reported by successive classification systems^14,16–18^ are therefore not competing approximations to a hidden universal catalogue but sample- and measurement-dependent partitions of a continuous space: refitting the winning discrete model across resampled folds changed which ears were grouped together even when predictive scores changed little.

Conventional configurations likewise occupied overlapping, contiguous regions rather than separate states (Figs. 2 and 5)^29,32,34,38^. Carhart explicitly acknowledged this arbitrariness, instructing that audiograms fitting two categories equally be assigned to the category closest to Flat^6^. Schuknecht and Gacek later reported mixed pathology and no consistent pathological correlate in about 25% of presbycusis cases^8^. Guild, Carhart and Schuknecht were not wrong in what they observed. They were wrong in what they inferred from those observations: a catalogue of kinds rather than a continuous space.

The Atlas also defines what the PTA retains and discards. The Global Burden of Disease framework estimates hearing loss from the better-ear average of thresholds at 0.5, 1, 2 and 4 kHz and assigns that scalar to severity bands^45^; by this measure, 1.57 billion people had hearing loss in 2019, of whom 1.17 billion (74.3%) occupied the single 20-34-dB mild band^45^. Yet audiometric structure was richest at intermediate severity, dimensionality peaking at 43.2 dB within the moderate band (Fig. 6e), and reducing the audiogram to four frequencies preserved overall level and some slope, but missed curvature and the 3-kHz feature (Fig. 6d). Independently, overall level accounted for only 42.84% of longitudinal movement through hearing space, against 57.16% for changes in audiogram shape. PTA and WHO grades remain practical summaries of burden^10,11^, but discard the dimensions precisely where most hearing diversity and change occurs.

Over 45 years ago, Bamford et al.^22^ described overall hearing level and audiogram slope as two dimensions of audiometric variation in a cohort of 201 ears, and Bench reproduced the same dimensions in an independent cohort of 391 ears^23^. Both studies used a six-frequency grid that omitted 3 and 6 kHz. Both also recovered a smaller third dimension: 8.21% in Bamford and 6% in Bench, which they dismissed as noise. Our independent population-scale study replicated the same three dimensions. We also explain Bamford-Bench’s finding of three dimensions and our finding of five. In 11,084 held-out ears from the US NHES II and III surveys (1963-1970), measured at 0.25, 0.5, 1, 2, 3, 4, 6 and 8 kHz, we removed 3 and 6 kHz to reproduce exactly the six-frequency grid used in their studies (0.25, 0.5, 1, 2, 4 and 8 kHz), while keeping the Atlas frozen. HAC3 and HAC5 then became unrecoverable. We independently repeated the same experiment in 1,638,338 held-out NIOSH ear-examinations, again removing 3 and 6 kHz, and HAC3 and HAC5 again became unrecoverable. The dimensions missed by their studies were therefore the dimensions that could not be recovered when 3 and 6 kHz were not measured.

Independent populations recovered the same hearing dimensions from their own data, and independently derived HACs transferred across historical surveys, contemporary populations and clinical data (Supplementary Figure S3, Supplementary Figure S3). Their recurrence across populations, eras, measurement standards and the physical route of measurement establishes HAC1 to HAC6 as reproducible dimensions of human hearing. The same coordinates located the same ear when measurement moved from air conduction to bone conduction, and one Atlas served both routes, so the structure is not a property of the air-conduction apparatus. The same dimensions that distinguish one person’s hearing from another’s also describe how an individual’s hearing changes over time. This correspondence was present in both chronic hearing change over years and acute Ménière fluctuations over hours^8,33^. This state-motion correspondence extends across timescales from hours to decades, is evidence that the Atlas captures biological organisation.

Taken together, these findings reveal a single organising principle: human hearing is not a collection of discrete states, but a continuous system whose structure is preserved across people, populations and time. We term this the Law of Human Hearing States (LHHS): human hearing has states, not types. These states occupy a continuous, low-dimensional space rather than forming discrete categories. The law is characterized by four empirically demonstrated invariants of those states (Fig. 3): 1) Continuity: hearing states occupy a continuous space rather than discrete types. 2) Low dimensionality: variation among hearing states is organised along a small number of independent dimensions, which form its coordinates. 3) Universality: the same state coordinates recur across populations, generations, clinical settings, audiometric standards and measurement routes, including air and bone conduction. 4) State-motion correspondence: the dimensions that distinguish hearing states between individuals are also the dimensions along which the same ear changes over time. The HHA is the coordinate map of this space; the LHHS describes the organisation and dynamics of the states it represents. Normal hearing therefore has no sharp natural boundary, and substantial audiogram-shape variation persists within it.

The Human Hearing Atlas (HHA) coordinate structure persists across air and bone conduction, supporting interpretation of the HHA as a representation of human hearing state rather than of a single measurement route. It does not by itself identify the anatomical or molecular cause of a given hearing state, leaving the relationship between cochlear states and hearing states open to investigation. Within these boundaries, the HHA provides hearing science and otology with a universal, interoperable quantitative reference frame for the study of hearing in health and disease. Across 33 datasets, eight countries, 62 years, two calibration standards and both routes of measurement, the same low-dimensional structure persisted. This stability changes the unit of description from a label to a location and the unit of longitudinal change from an arbitrary category to a trajectory. Cochlear regenerative and gene therapy is now a reality, and the need for a more faithful representation of hearing is becoming acute, where emerging interventions produce frequency-specific longitudinal treatment responses that conventional PTA cannot fully describe^46–48^. The REGAIN regenerative-drug trial explicitly identified the inability of current auditory tests to provide deep phenotyping as a limitation^47^. Its PTA across 2, 4 and 8 kHz did not change significantly, although substantial individual-frequency changes occurred in subsets of participants; similarly, FX-322 produced speech-intelligibility improvements without corresponding group-level PTA improvement^48^. Human OTOF gene therapy now produces gradual multiyear trajectories of behavioural and electrophysiological hearing recovery^46^, while preclinical CLIC5 gene replacement produces frequency specific changes that the PTA misses^49^. The HHA addresses exactly this limitation through a quantitative framework for deep audiometric phenotyping, allowing patient selection, efficacy assessment and monitoring therapeutic response on short or long-term as multidimensional movement through hearing state rather than as a change in a single severity average and arbitrary categories. Genetic and epidemiological studies can replace heterogeneous case labels with continuous traits; registries can compare hearing across sites and eras; and hearing rehabilitation can be monitored against multidimensional change rather than PTA alone^50^. What the Atlas changes is more fundamental: after 140 years of asking what type of hearing an ear has, the audiogram can instead be read as a position in a universal human hearing space. Human hearing has states, not types; hearing change is movement through those states. The audiogram was not a catalogue of kinds. It was a map.

## Materials and Methods

### Study design and reporting

This observational study combines cross-sectional population audiometry with longitudinal analysis of repeated occupational audiograms and serial audiograms collected during acute Ménière episodes. The unit of analysis is the ear. No new ethical approval was required for this secondary analysis of de-identified data. Ethics approval, consent and governance for the original source cohorts are detailed in Supplementary Table S6c.

### Data

We assembled the largest audiogram corpus in a single analysis reported to date in the peer-reviewed literature, comprising 33 datasets of pure-tone air-conduction audiometry spanning six decades (1963-2025) and eight countries: the United States, South Korea (KNHANES), Germany (Oldenburg Hearing Health Record, OHHR, and the Münster glycerol series of Lütkenhöner and Basel), Denmark (BEAR and two Danish deposits), Türkiye, the United Kingdom, Australia and Iran. Twenty-three of these are population survey waves; the remainder are clinical, occupational and experimental cohorts reserved for external validation. Together they comprise 4,291,863 ear-curves from 1,464,402 people; the full dataset inventory is given in Supplementary Table S6. The United States contributed six successive federal survey programmes - NHES Cycles II and III, NHANES I and II, the Hispanic HANES and continuous NHANES - whose 1960s to 1980s files required reconstruction from their original documentation. The source datasets contained 210,319 records, which resolved to 208,041 audiograms across the 25 waves. After quality control, 201,387 ears from 100,904 people remained. The subtype analysis included 195,262 ears from 97,631 people with complete bilateral audiograms, retaining both ears only when both were complete across the frequencies tested. In a sensitivity analysis, participants contributing only one eligible ear were restored and the discrete-versus-continuous comparison repeated under otherwise identical settings. The six-frequency corpus comprised 117,464 ears from the 15 waves measuring 0.5, 1, 2, 3, 4 and 6 kHz. The final Atlas was derived from the 89,752 ears recorded under International Organization for Standardization and American National Standards Institute (ISO/ANSI) calibration standards. The remaining 27,712 ears came from the two ASA-1951 waves and were withheld from the ISO/ANSI construction for external historical validation (Supplementary Methods S3).

### Testing for distinct subtypes

The discrete model assigned audiograms to mutually exclusive groups, whereas the continuous model allowed variation without forced assignment. Both were fitted on development data and compared by prediction in held-out participants. A cohort was considered discrete only when the subtype model consistently outperformed the continuous model out of sample. All models used the complete set of thresholds for each ear; the PTA was used only to group ears by severity and as a comparator, never as a model input.

### Minimum group size

A recovered subtype was required to contain at least the square root of the development-sample size; rarer audiogram configurations were not tested as separate subtypes. The rule was implemented as a mixture-weight floor: within each inner training split, any solution containing a component of weight below 1/√N was disqualified, N being the number of ears in that split.

### Age and sex groups

The test was applied to age groups within a wave (youth, working age and senior, aged 60 years or older), as well as to whole waves and to combined series. The source datasets recorded sex as a binary variable. Analyses were by country of survey; ethnicity was not compared because coding differed between datasets, and one US wave (the Hispanic HANES, 1982-1984) is itself an ethnicity-defined national sample.

### Reproducibility of the subtype count

Subtype-count stability was assessed within the same 100 participant-grouped outer validation folds used for discrete-versus-continuous model comparison. Each fold omitted approximately 1.0% of development participants, so any two fold-specific training samples differed in approximately 2.0% of participant membership. For each fold, component count was selected independently under the interval-aware criterion while population, frequency grid and model family remained fixed. For each of the 29 discrete-verdict analyses, every fold-selected model was refitted and, together with a taxonomy fitted on the full development set, applied to the same locked-holdout ears. Partition agreement was quantified by the adjusted Rand index; ear-level membership by a label-invariant co-assignment statistic; and predictive difference by interval likelihood on the identical holdout under one shared quadrature draw. Full definitions are provided in Supplementary Methods S2.

### Deriving the canonical coordinate map

The canonical Atlas was derived from the 89,752 ISO/ANSI ears alone, so that the reference frame rests on the modern international audiometric standard rather than on a mixture of modern and historically converted measurements. After frequency-specific conversion, the ASA-era ears were included only in a separately reported sensitivity analysis assessing stability of the recovered coordinate structure. HAC1 was defined as overall hearing level. Shape coordinates were derived from the remaining audiogram variation after separating overall level. Each coordinate was named according to the change in audiogram shape associated with increasing values of that coordinate. Full detail is given in Supplementary Methods S3.

### Coordinate stability

We tested whether the same shape patterns were recovered when each cohort was analysed separately and when the analysis was repeated in different samples of participants. Agreement between coordinate loadings was measured by Tucker’s congruence coefficient after matching coordinates in order and resolving the arbitrary sign of each singular vector. Axis agreement is reported as an observed value for every coordinate; no prespecified acceptance threshold was applied. Reconstruction accuracy was recomputed across five independent development-validation splits, and the range across those splits is reported alongside the point estimate.

### Independent recovery from unlabelled cohorts

Native-grid audiograms were analysed in raw dB with equal ear weights and participant or worker clusters; NIOSH retained each worker’s earliest complete bilateral examination. Each cohort was decomposed on its own grid without access to the Atlas coordinates, rank or labels, separating overall hearing level from the remaining shape variation. Solutions were fixed before any comparison. Axes were matched objectively and one to one, and agreement was summarised by congruence; uncertainty used 2,000 cluster bootstraps, and recovery required the 2.5th percentile to exceed the 99.9th percentile of a random-orientation null of 200,000 draws evaluated identically. For zero-shot transport, the NIOSH coordinate system was frozen and applied unchanged to NHES II, NHES III and Münster, with no fitting or adaptation to the receiving dataset. Functional transfer withheld 3 and 6 kHz from NHES, predicted them from the frozen NIOSH representation, and compared joint MAE with log-frequency interpolation using 2,000 paired cluster bootstraps.

### Transfer between populations

Portability was quantified as the reconstruction-error difference between a frozen donor coordinate system applied unchanged to an independent population and a coordinate system fitted within it. This transfer penalty is reported in decibels with an interval; penalties spanning zero are reported as not detectable. Transfer ran bidirectionally between the ISO/ANSI US waves and Korean surveys: the US-ISO coordinates, derived from 46,184 ears, were applied to 10,892 Korean ears from 5,446 held-out participants; the Korean coordinates, derived from the Korean development sample, were applied to 11,546 US-ISO ears from 5,773 held-out participants. Intervals are 95% percentile intervals from 2,000 participant-grouped bootstrap resamples of the paired per-ear difference, resampling participants with replacement and carrying both ears together.

### External validation on other frequency grids

Twenty datasets were reserved for external validation, contributing neither to Atlas derivation nor tuning, spanning population, clinical, occupational and experimental cohorts. The frozen coordinates were applied according to each dataset’s measured frequency grid, with coordinates requiring unmeasured frequencies reported as unavailable, and reconstruction accuracy compared where possible with a coordinate system fitted locally within that cohort. The German cohort contributed 1,138 ears from 569 participants measured at 0.125, 0.25, 0.5, 0.75, 1, 1.5, 2 and 4 kHz, and the Danish cohort 172 ears from 86 participants measured at 0.25, 0.5, 1, 2, 4 and 8 kHz; neither measures 3 or 6 kHz. Both were analysed on the 0.5, 1, 2 and 4 kHz grid shared with the canonical set, and the universal coordinates applied to each were derived from all eligible ears outside that country.

### Historical validation across calibration eras

The 27,712 ASA-1951 ears, comprising children and adolescents aged 6 to 18, provide an external test of whether the coordinates survive a change of calibration standard, being withheld from the ISO/ANSI construction for that purpose. American Standards Association (ASA) 1951 thresholds were converted during decoding using frequency-specific corrections taken from the calibration standard rather than estimated from the data: +14.0 dB at 0.5 kHz, +10.0 dB at 1 kHz, +8.5 dB at 2 and 3 kHz, +6.0 dB at 4 kHz and +9.5 dB at 6 kHz. The canonical ISO/ANSI coordinates were applied to them frozen and reconstruction accuracy recorded. We separately refitted the coordinate system with the ASA-era waves included and compared the two coordinate systems by Tucker’s congruence, expressing the difference as a reconstruction difference in decibels rather than as a correlation (Supplementary Methods S8).

### Measurement-route invariance

Paired air- and bone-conduction thresholds at 0.5, 1, 2 and 4 kHz came from the first US NHANES, 13,826 ear-records from 6,913 participants measured in the same ears at one visit. The source carries parallel bone-conduction series of undocumented relationship; these were never averaged or pooled, every analysis ran separately in each series, and a result was required to hold in all of them. The canonical Atlas was applied unchanged to bone conduction, without refitting, rotation, rescaling or realignment. Same-ear correspondence across routes was tested against candidate ears matched for side, age and hearing level, and repeated twice with severity controlled: once with the level term structurally deleted, and once restricted to ears whose measured air-conduction thresholds were all normal. A model fitting separate route-specific representations together with the mapping between them was compared against the frozen Atlas plus a route operator on identical folds, identical test ears and identical regularisation, with the two-representation model never receiving fewer fitted parameters; noninferiority was assessed against a 0.5 dB margin fixed before results, and the comparison was repeated across subsample sizes to establish saturation. Reconstruction was evaluated in held-out participants in both directions, from air conduction to bone conduction and from bone conduction to air conduction. The conventional scalar air-bone gap entered only as the reduced comparator against the full cross-route displacement. All splitting, bootstrap resampling and permutation carried both ears of a participant together, and intervals are participant-clustered bootstrap percentile intervals (Supplementary Methods S25).

### Which coordinates a frequency grid can observe

To determine which coordinates a reduced grid can recover, we masked complete six-frequency audiograms to 0.5, 1, 2 and 4 kHz, estimated the canonical coordinates from those four thresholds, and compared them with coordinates from the complete audiogram. Linear estimators were fitted only in source-country development participants and evaluated in held-out participants (Supplementary Methods S9). The recovery statistic is the proportional reduction in the variance of a coordinate achieved by estimating it from the masked grid, relative to its variance in the source population, reported alongside the correlation with the complete-audiogram coordinate. A coordinate was declared recoverable when that variance reduction reached the prespecified 0.80; the recovery value for every coordinate is reported. This masking analysis estimates recovery of the canonical HAC and is distinct from fitting a new reduced-grid coordinate system.

A sixth coordinate, representing the remaining local 1 kHz difference, is required only to reproduce all six thresholds exactly. For reduced-grid transfer analyses, four-frequency audiograms were represented by overall hearing level and the shape coordinates derived on that reduced grid.

### Frequency-grid replication of the Bamford-Bench studies

To test the effect of the frequency grid used by Bamford et al. and Bench, we restricted held-out NHES II and III audiograms to their historical frequencies of 0.25, 0.5, 1, 2, 4 and 8 kHz, thereby withholding 3 and 6 kHz relative to the canonical Atlas grid. The canonical Atlas remained frozen and was never refitted. Recovery of HAC1 to HAC5 from the restricted audiograms was quantified by the same variance-reduction statistic used for frequency-grid observability, with 0.80 prespecified as the recoverability threshold, and the withheld 3 and 6 kHz thresholds were reconstructed from the restricted grid and scored as mean absolute error. The analysis used 11,084 held-out NHES II/III ears. As an independent replication, 3 and 6 kHz were jointly withheld from 1,638,338 held-out NIOSH ear-examinations and coordinate recovery was evaluated under the same frozen-frame criterion.

### Reconstruction accuracy and preserved distinctions

Reconstruction was measured in people not used to derive the map, as mean absolute error in decibels and as the proportion of thresholds and of whole ears reconstructed within 5 dB. Two audiograms were considered different when they differed by more than 5 dB at any measured frequency, and we measured how often reconstruction made such pairs indistinguishable. Preserved distinctions were evaluated from 2,000,000 audiogram pairs sampled with replacement within the held-out target set, with self-pairs discarded. Full detail is given in Supplementary Methods S5.

### Full-spectrum validation across normal and impaired hearing

The primary composite conclusion required three prespecified criteria to be met using the unchanged Atlas representation. First, overall hearing level was removed, and the retained variation in HAC2-HAC5 was compared between normal and impaired ears. Equivalence required the retained proportion in normal hearing to be no more than 5 percentage points lower than in impaired hearing. Second, abrupt changes in reconstruction error were tested at 20 dB HL, with 15 and 25 dB HL as prespecified additional cutoffs; the acceptable reconstruction-error change was 0.5 dB. Third, normal-hearing transfer was tested in both national and historical directions with a 0.5 dB acceptability limit.

### How many independent dimensions an audiogram contains

We estimated the number of independently varying audiogram dimensions using a correlation-dimension method; the construction is given in Supplementary Methods S4. Correlation dimension was estimated from 40 logarithmically spaced radii spanning 1-150 dB, with the slope fitted over the prespecified 9-28 dB window. Threshold vectors were not standardised, Euclidean distances were calculated in decibels, and at least five valid radius points within the fitting window were required. This was done in 75 age- and wave-defined cohorts. Because the estimate cannot exceed the number of frequencies tested, comparisons were made only between cohorts tested on the same frequencies.

### Identical audiograms

Thresholds are recorded in 5 dB steps,^39–41^ so at low severity many ears share exactly the same set of thresholds. We counted the proportion of exact duplicates in every cohort and repeated the analysis with repeated threshold patterns counted only once. Repeated threshold patterns were counted once only when estimating the number of independent dimensions, where identical vectors carry no information about separation; all ears were retained when deriving the Atlas coordinates.

### Longitudinal hearing change

We applied the frozen Atlas to repeated audiograms from the NIOSH occupational hearing-conservation archive. Examinations from the same worker and ear were ordered in time and successive audiograms were represented using the same coordinates as the cross-sectional analyses. We measured how accurately changes in the first five coordinates reconstructed the observed change at each frequency and separated total longitudinal movement into change in overall hearing level and change in audiogram shape. Workers, rather than individual examinations, were used as the resampling unit. Full computational definitions and sensitivity analyses are provided in Supplementary Methods.

### Comparing differences between patients with changes within the same ear

To determine whether the same audiogram dimensions describe population differences and longitudinal change, we independently identified the major patterns of audiogram shape across different ears and across repeated measurements of the same ears. Overall hearing level was removed before this comparison so that the analysis tested changes in slope, curvature and local frequency differences rather than severity alone. We quantified how strongly the dominant patterns from the two analyses overlapped and compared the observed correspondence with randomly oriented hearing patterns. Examination order was permuted within workers to determine whether the correspondence depended on chronological direction. Full mathematical definitions and null constructions are provided in Supplementary Methods.

### State dependence of longitudinal hearing change

To test whether an ear’s current hearing state influenced how it subsequently changed, hearing space was divided into prespecified regions and the subsequent changes of ears beginning within each sufficiently populated region were compared. Regions with insufficient longitudinal observations were left unevaluated rather than smoothed or interpolated. We quantified how much variation in subsequent change was associated with starting location and compared the observed value with a worker-level permutation analysis.

### Acute Ménière trajectories

The acute analysis comprised 356 unilateral-ear series with five successive audiograms. Each audiogram was converted to the same frozen Atlas coordinates, allowing the episode to be represented as movement through hearing space over time. We then compared the major audiogram-shape patterns observed during these acute series with those observed during chronic longitudinal change, after removing overall hearing level from both analyses. We additionally tested whether the chronic eigenspace, frozen and unadapted, could reconstruct acute successive-transition shape change, quantified as the ratio of variance captured by the frozen frame to that captured by a rank-2 frame fitted directly to the acute transitions, benchmarked against 2,000 random rank-2 orientations for both variance captured and reconstruction error. Mathematical details are provided in Supplementary Methods S18.

No statistical method was used to predetermine sample size; all eligible records passing the prespecified quality-control rules were used. The study is observational; randomisation and blinding were not applicable. Sex was analysed as recorded by the source datasets, and no sex-specific biological inference was made. The pooled four-country analysis comprised the prespecified census of 170,028 ISO/ANSI referenced ears.

### Statistical analysis

The discrete model was a finite mixture fitted across two to thirty components using a restricted multivariate skew-normal family; the continuous comparator was selected from thirty single-population densities without discrete component structure (Supplementary Methods S1). Threshold vectors were standardised within each split. Because clinical thresholds are recorded on a quantised 5 dB grid, each recorded threshold was represented by its corresponding observation interval rather than as an infinitely precise value^40,42^. The same interval-based observation model was applied to the discrete and continuous families, with ceiling and floor codes treated as one-sided tails.

Model selection and comparison were performed out of sample using participant-grouped nested validation, with both ears of a participant always assigned to the same split. Component count and model specification were selected within the training data, refitted on the outer-training sample and evaluated in untouched outer-test participants. Final inference used a locked holdout that took no part in fitting or selection. The reported margin was the difference in held-out predictive log-likelihood between the discrete and continuous models; its uncertainty was estimated from 10,000 participant-level bootstrap resamples. A cohort was classified as discrete when the margin and its 95% interval lay entirely above zero, continuous when both lay below zero, and inconclusive when the interval contained zero (Supplementary Methods S1).

To estimate how dimensionality varied with hearing level, we fitted a quadratic fixed-effects regression of correlation dimension on severity-band midpoint after within-cohort demeaning, using deduplicated estimates with fit R^2^ ≥ 0.99 from 65 cohorts and 284 cohort-band observations. Uncertainty in the location of the maximum was estimated using 5,000 cohort-cluster bootstrap replicates. Data decoding, harmonisation, computations and analyses were implemented in Rust, Julia and C++ and executed on Roihu, Finland’s national supercomputer operated by CSC, IT Center for Science, using 20-60 compute nodes corresponding to 7,680-23,040 CPU cores and 15.7-47.2 TB of memory.

## Data Availability Statement

Source datasets analysed in this study remain available from their originating agencies or repositories under their respective access conditions, licences and data-use requirements. The dataset-specific provenance and access routes are described in the Methods and the full dataset list in Supplementary Table S6. No source record-level data are redistributed by this study. Aggregate numerical data supporting the reported analyses and display items are provided where permitted by the governing source terms.

## Code availability

The code necessary to reproduce the findings reported in this study may be made available for academic verification upon reasonable request, subject to applicable data-use and institutional requirements.

## Acknowledgments

We thank Dr Davide Venturelli, Fellow at the Universities Space Research Association Research Institute for Advanced Computer Science in Mountain View, California, USA, for subjecting the study’s methodology, validation strategy and statistical analysis to rigorous, independent critical scrutiny. We are deeply grateful to CSC, IT Center for Science, Finland, for providing access to Roihu and its national high-performance computing infrastructure, which made the scale and breadth of the analyses reported in this study possible. This work was supported by The Finnish ORL-HNS Foundation, Finland. The funder had no role in study design, data collection, data analysis, interpretation, or the decision to submit for publication.

## Author Contributions

ZHA conceived and designed the study; developed the methods for subtype testing, atlas construction and dimensionality analysis; retrieved, decoded and harmonised all datasets; built and implemented the analysis pipeline; performed all computational analyses; interpreted the scientific and clinical findings; identified, defined and described the HHA and the LHHS, and created all figures. AA and AM supervised the work and revised the manuscript. ZHA, AA and AM directly accessed and verified the underlying data reported in this manuscript. All authors had full access to all the data in the study, approved the final version, accept responsibility to submit for publication, and are accountable for the accuracy and integrity of the work.

## EXTENDED DATA

**Supplementary Figure S1.**
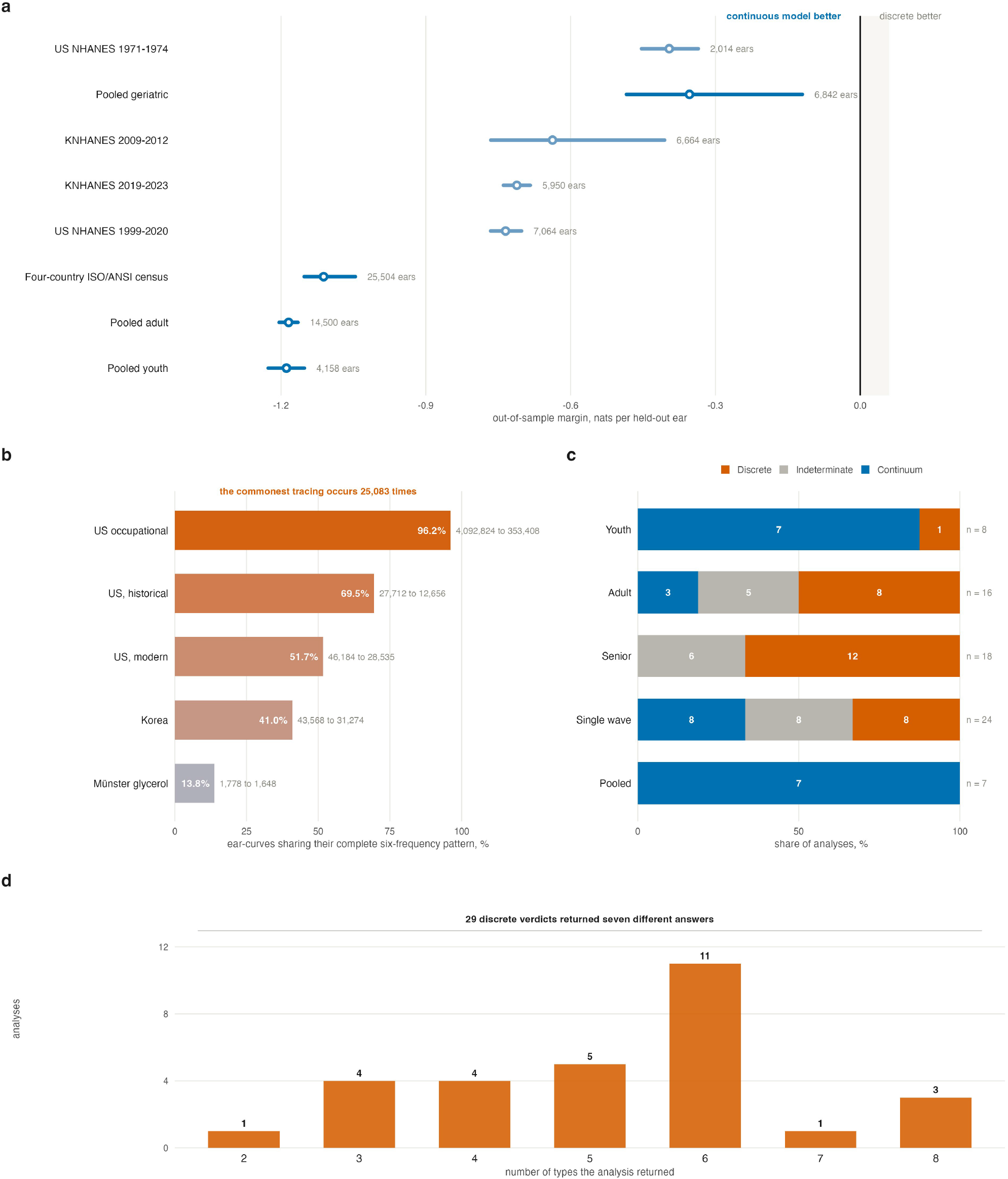
| Audiogram types are artefacts of measurement and sampling, not biological kinds.(a) Out-of-sample comparison of a discrete and a continuous model of the audiogram, scored on locked holdouts of participants that took no part in fitting, model selection or component-count selection. The margin is the discrete minus continuous predictive score per held-out ear, so negative favours the continuum. Intervals are 95% percentile intervals from 10,000 participant-grouped bootstrap resamples.(b) Clinical 5 dB recording concentrates distinct ears onto identical tracings. Bars give the percentage of ear-curves sharing their complete six-frequency pattern with at least one other ear; the figure at the right of each bar gives ear-curves and the distinct patterns they collapse onto. In the occupational archive 4,092,824 ear-examinations collapse onto 353,408 distinct recorded patterns, and the commonest single tracing occurs 25,083 times. Families whose grid cannot carry the canonical six frequencies are excluded.(c) Verdicts across all 73 analyses, by stratum. A verdict is continuum when the margin and its entire interval lie below zero, discrete when both lie above zero, and indeterminate when the interval contains zero. Discrete verdicts concentrate in age-restricted strata: no senior stratum returned a continuum. Every one of the seven pooled analyses did.(d) The number of types recovered by the analyses that did return a discrete verdict. Twenty-nine discrete verdicts returned seven different answers, from two to eight; across participant-grouped outer folds, 1,569 of 2,884 evaluable fold fits (54.4%) selected a count different from the reported value.

**Supplementary Figure S2.**
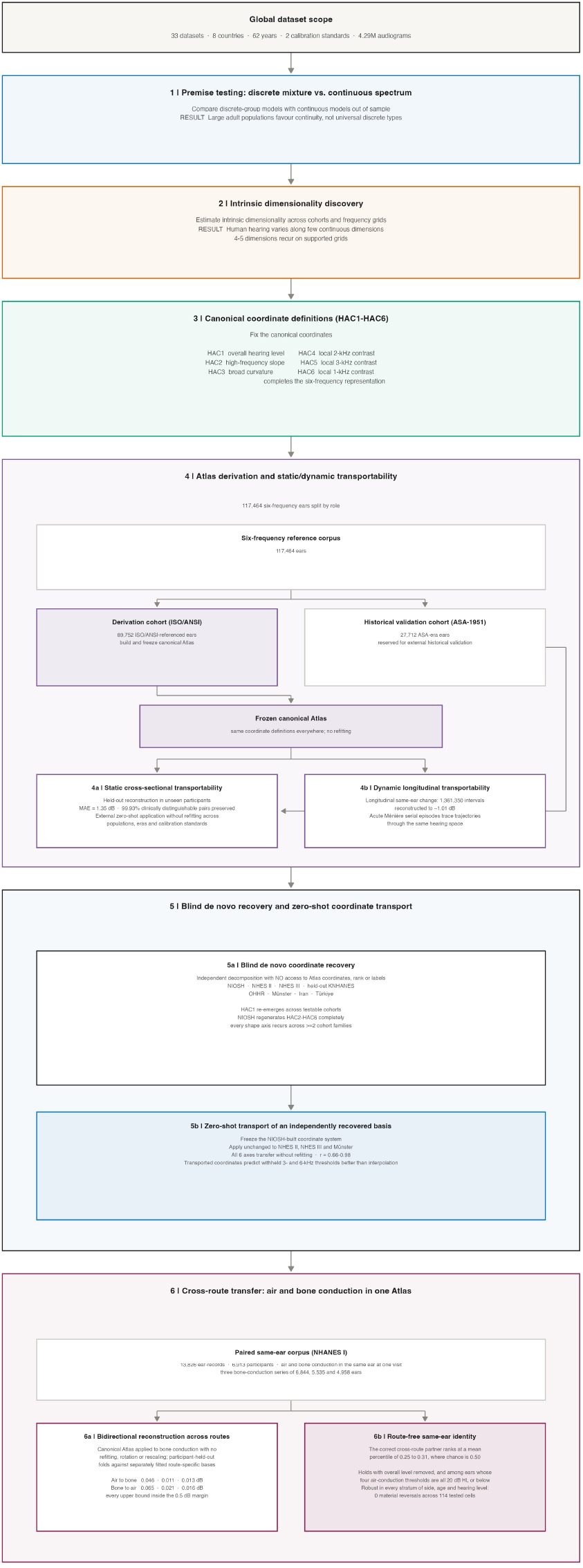
| Study design and validation architecture of the Human Hearing Atlas.Six sequential stages, read top to bottom. Each block gives the question asked, the data used and the result. The header states the study scale: 33 datasets, eight countries, 62 years, two calibration standards and 4.29 million audiograms.Stage 1, premise testing: discrete mixture models against a continuous spectrum, scored out of sample on locked holdouts. Stage 2, intrinsic dimensionality estimated from raw thresholds across audiometric grids. Stage 3, definition of the canonical coordinates HAC1 to HAC6 on the frozen derivation frame of 89,752 ear-curves. The block shows the full six-frequency corpus of 117,464 ears split by role: the remaining 27,712 are the ASA-1951 ears, withheld from derivation for historical external validation.Stage 4, Atlas derivation and transportability: 4a, static validation by participant-held-out reconstruction, reaching a mean absolute error of 1.35 dB with 99.93% of clinically distinguishable pairs preserved, followed by transfer to cohorts that contributed nothing to derivation; 4b, dynamic longitudinal transfer to repeated audiograms of the same ear, with no longitudinal representation fitted.Stage 5, independence: 5a, blind de novo coordinate recovery in NIOSH, NHES II, NHES III, held-out KNHANES, OHHR, Münster, Iran and Türkiye, each decomposed on its own native grid without access to Atlas coordinates, rank or labels; 5b, zero-shot transport of an independently recovered basis, in which all six axes transferred without refitting (r = 0.66 to 0.98).Stage 6, cross-route transfer: 6a, bidirectional reconstruction between air and bone conduction in 13,826 paired ear-records from 6,913 participants of the first US National Health and Nutrition Examination Survey, in whom both routes were measured in the same ear at one visit. The canonical Atlas was applied to bone conduction without refitting, rotation or rescaling. Against models permitted to fit separate route-specific representations, the shared Atlas cost 0.046, 0.011 and 0.013 dB reconstructing bone conduction from air conduction across the three bone-conduction series, and 0.065, 0.021 and 0.016 dB in the reverse direction, every upper confidence bound falling inside the prespecified 0.5 dB noninferiority margin; 6b, route-free same-ear identity, in which the correct cross-route partner ranked at a mean percentile of 0.25 to 0.31 against a chance value of 0.50, with the pattern persisting after overall hearing level was removed and among ears whose four air-conduction thresholds were all 20 dB HL or below.

**Supplementary Figure S3.**
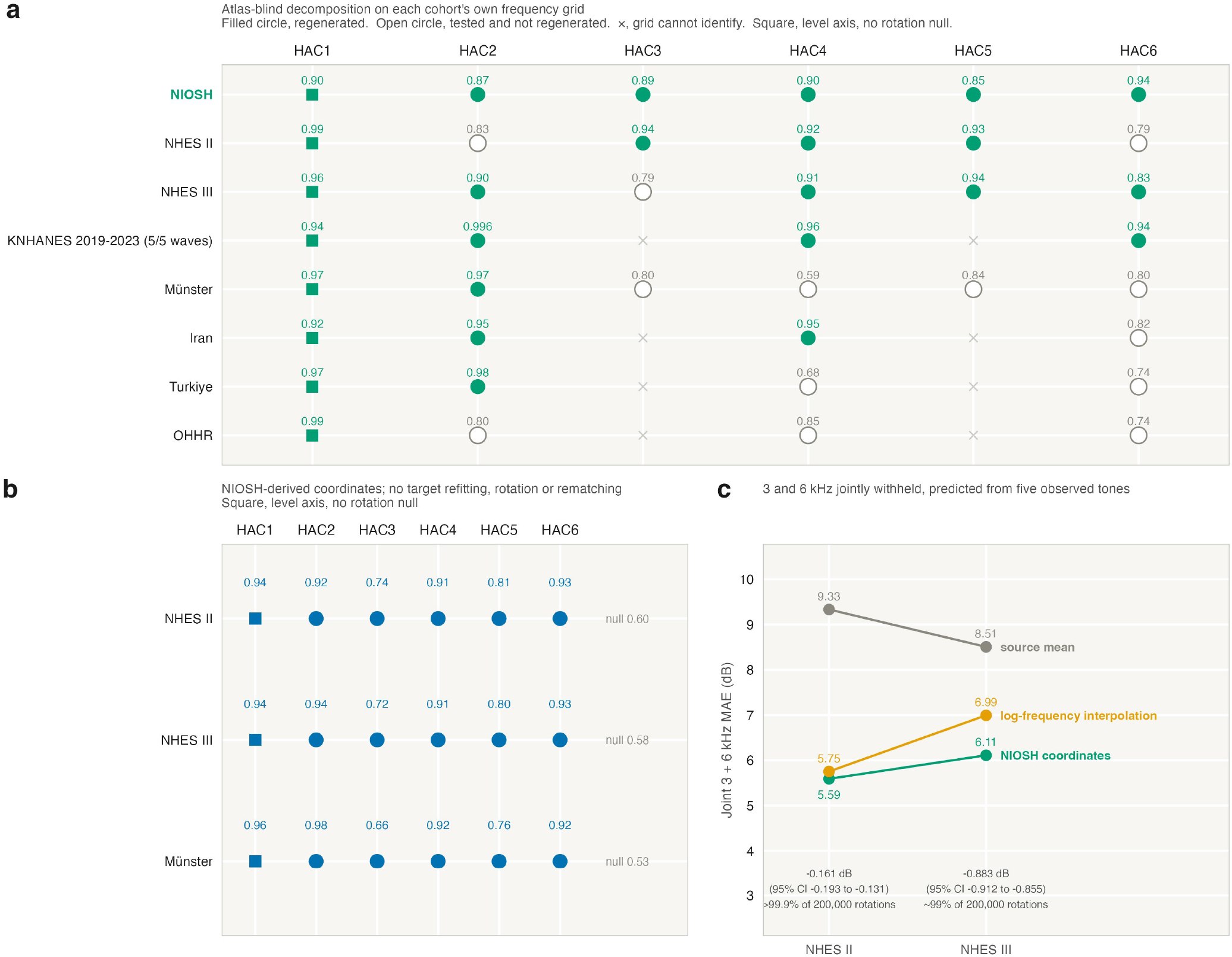
| Independent populations reconstruct and transport the Atlas coordinates.(a) Independent regeneration. Each cohort was decomposed on its own native grid in raw decibels without access to the Atlas basis, rank, axes or labels. Values are absolute congruence after Hungarian matching. Filled circles are axes whose 2.5th bootstrap percentile over 2,000 cluster resamples exceeded that comparison’s own 99.9th-percentile null, built from 200,000 random orthonormal bases. Open circles were tested and did not clear that null. Crosses are coordinates the cohort’s grid cannot identify, excluded in advance. Squares are HAC1, the level axis, which has no rotation null and is not part of the recovery test. The highlighted row marks the complete HAC2 to HAC6 regeneration in 1,365,737 NIOSH workers.(b) Zero-shot coordinate transfer. The coordinate system reconstructed from NIOSH alone was applied unchanged to NHES II, NHES III and Münster, with no target fitting, centring, rotation or rematching. Values are the Pearson correlation between transported and Atlas scores; the number at the right of each row is that cohort’s 99.9th-percentile null. HAC2 to HAC6 exceed that null in every cohort. (c) Zero-shot functional prediction. Five tones were observed and 3 and 6 kHz jointly withheld, then predicted from the frozen NIOSH covariance, from log-frequency interpolation, and from the NIOSH source mean. Annotations give the paired difference against interpolation with its 95% confidence interval from 2,000 paired cluster bootstraps, and the rank of the transported system among 200,000 spectrum-preserving random rotations.

**Supplementary Figure S4.**
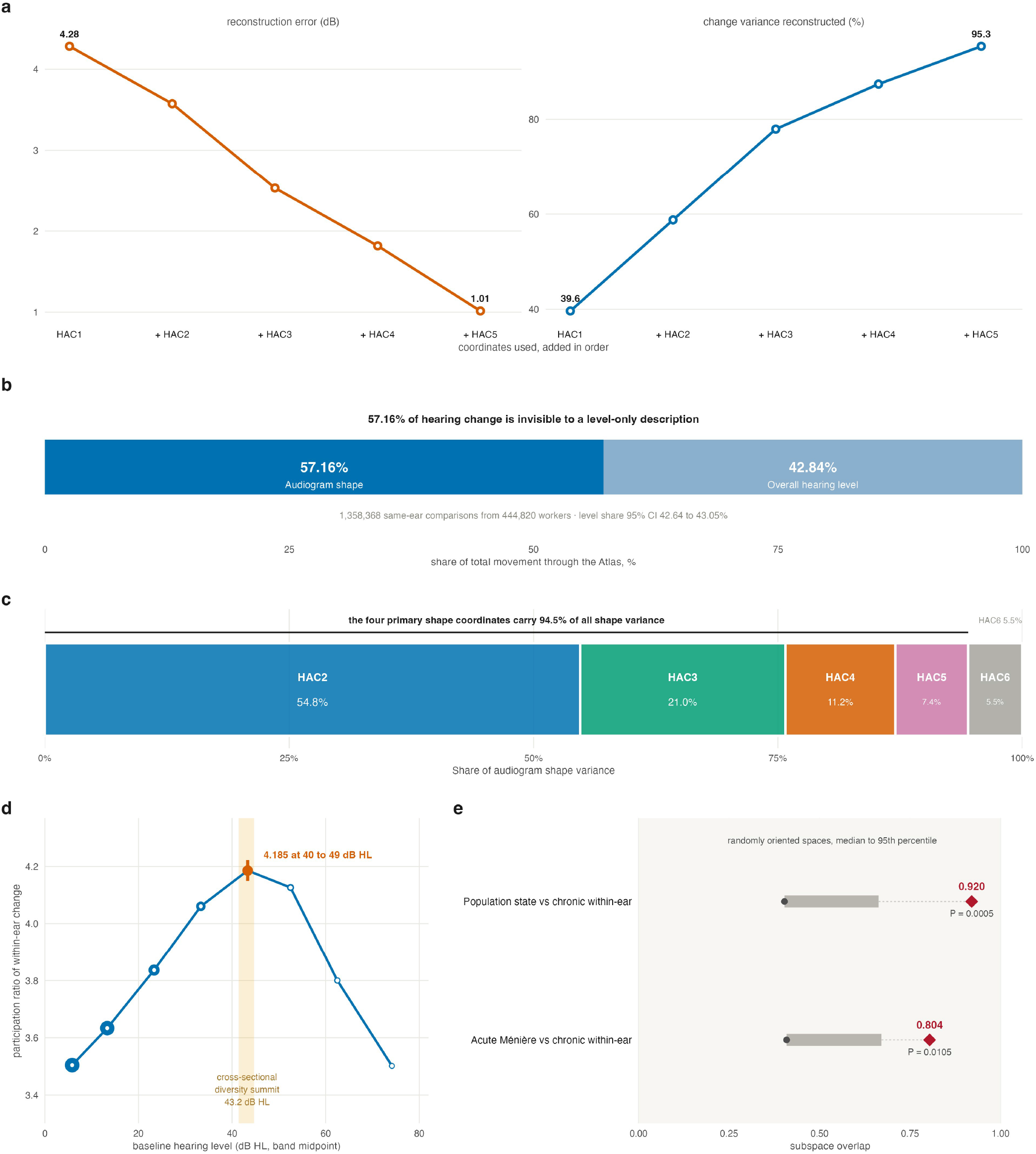
| Hearing change is multidimensional movement through the Atlas.Across 1,361,350 successive same-ear comparisons from 446,146 workers, coordinates derived cross-sectionally were applied unchanged; no longitudinal representation was fitted.(a) Frozen-coordinate reconstruction of within-ear change. Error falls from 4.28 dB with overall level alone to 1.01 dB with five coordinates, while reconstructed variance rises from 39.6% to 95.3%.(b) Decomposition of 1,358,368 comparisons from 444,820 workers. Overall level accounts for 42.84% of longitudinal movement (95% CI 42.64-43.05) and audiogram shape for 57.16% in the isometric embedding.(c) Cross-sectional shape-variance partition in 89,752 derivation ears after removal of HAC1: HAC2, 54.8%; HAC3, 21.0%; HAC4, 11.2%; HAC5, 7.4%; HAC6, 5.5%. The four primary shape coordinates carry 94.5% of residual shape variance. These percentages describe cross-sectional shape variance and are distinct from the longitudinal level-shape partition in (b). Block widths are proportional to variance share.(d) Complexity of longitudinal change by baseline hearing level, measured by participation ratio, peaks at 4.185 (95% CI 4.150-4.222) in the 40-49 dB HL band (median, 43.3 dB HL). Marker area reflects sample size; shading marks the independently estimated cross-sectional dimensionality summit at 43.2 dB HL (Fig. 6e).(e) Between-person and chronic within-ear shape spaces overlap by 0.920 (95% CI 0.918-0.922) across 1,294,932 changes in 421,868 workers; acute Ménière and chronic directions overlap by 0.804. Diamonds show observed overlaps; grey bars span the median to 95th percentile of randomly oriented spaces.

