## Supplementary Material for "The Human Hearing Atlas: a canonical map of human hearing"

---

### S1. Model specification for the discreteness test

For each cohort, a model assuming a fixed number of audiogram groups was compared with a model allowing audiograms to vary continuously; both were developed on one set of participants and evaluated in participants not used to develop either model, with both ears of each participant assigned to the same side of every split.

Unit of analysis and grouping. The unit is the ear. Both ears of a person are perfectly correlated for the purposes of this test, so any split that separates them lets one ear predict its fellow and manufactures apparent structure. All partitions were therefore formed at the person level, and every reported comparison is person-grouped.

Fixed-group model. A finite mixture over the ear's threshold vector, fitted for a ladder of component counts from two to thirty. The component family is a multivariate skew-normal, broad enough to accommodate skewness; a full-covariance Gaussian mixture is available as a cross-check family. Skew-normal components absorb the marginal skewness of audiometric loss, so a smooth but skewed population is not divided into spurious axis-aligned pieces. The degeneracy guard rejects any fitted component covariance that is near-singular, and a solution violating it is rejected. The component-weight floor is the inverse-square-root policy, one over the square root of the development-sample size.

Continuous model. The continuous comparator comprised a family of candidate densities over the same threshold vector, each representing variation along a continuous latent structure rather than membership in mutually exclusive groups. The candidates spanned a single density of the same family, linear factor analysers, a Gaussian copula, principal-curve models across a range of curve flexibility and occupancy bandwidth, and a penalised-spline curve density, so that nonlinear continuous structure could be represented as well as linear. The winning continuous model was selected exclusively within the development sample using the same nested validation procedure used to select the categorical component count.

Observation model. Clinical thresholds are recorded on a 5 dB grid, so a recorded value denotes an interval. Every candidate, categorical and continuous alike, is therefore scored by the probability it assigns to the observed event : for an ordinary recorded value v at each frequency, the probability of the box spanning v ± 2.5 dB; for a value at the recording ceiling, the corresponding one-sided upper tail; and for a value at a floor code, the corresponding one-sided lower tail. Applying the identical observation model to all candidates is essential, because a density comparison on quantised data otherwise rewards whichever family happens to place more mass at lattice points. Box probabilities were evaluated by quasi-Monte Carlo integration, except for the penalised-spline family, whose exact box probability has no closed form and was approximated. Integration tolerance was verified by doubling the number of integration points on the calibration cohort and confirming that the margin moved by far less than the smallest margin of scientific interest.

Estimation under the observation model. Correcting the score alone would leave parameters estimated under the uncorrected measurement model. For the full-covariance Gaussian cross-check family, both estimation and evaluation use the interval likelihood: an exact interval-censored expectation-maximisation procedure whose expectation step uses box probabilities and whose maximisation step uses importance-weighted truncated moments. For the skew-normal mixtures and the curve families, exact interval-likelihood estimation is not tractable at the scale of the nested cross-validation, and parameters are estimated by point-likelihood maximisation and then scored under the interval observation model; these families are labelled as point-fitted throughout and are never described as fully interval-corrected. The prespecified robustness route for them is a matched repeated-dequantisation analysis, in which the recorded value at each frequency is replaced by a uniform draw within its 5 dB interval, using identical draws across all competing candidates, with the analysis repeated across replicates.

Calibration of the discreteness test. The comparison was calibrated against six synthetic populations with known structure before application to any study cohort: a smooth continuum; the same continuum recorded on a 5 dB grid; the same continuum with ceiling censoring; the continuum with both effects combined; separated groups arranged along a common direction; and separated groups differing in audiogram shape. Four scenarios required a continuous verdict and two required a discrete verdict, preventing selection of a configuration that systematically favoured either conclusion.

The final configuration was selected by a two-sided maximin criterion over the prespecified ladder of curve flexibility, regularisation and occupancy bandwidth. It returned the correct verdict in all six synthetic scenarios, with no false-continuous, false-discrete or indeterminate outcomes. The occupancy density of the curve models was constrained to remain smooth and unimodal, preventing the continuous comparator from representing separated groups as multiple peaks along the curve. The simpler comparator containing only the single density, factor analysers and copula classified all four synthetic continua as discrete, whereas the penalised-spline family alone classified both grouped scenarios as continuous. Both the curve-based and componentless continuous families were therefore required for calibrated discrimination in both directions.

Decision rule. Each cohort was first divided at the participant level into a development sample of 85% of participants and a locked holdout of the remaining 15%, with both ears of a participant kept on the same side and the split seed fixed in advance. All model comparison, component-count selection and continuous-variant selection took place inside the development sample. Once every modelling choice was frozen, the selected discrete and continuous models were fitted once on the full development sample and scored once on the locked holdout. The predictive score is the held-out log-likelihood, and the categorical-minus-continuous margin is divided by the number of held-out ears and reported in nats per ear. The verdict is the sign of the holdout margin together with its interval: a cohort is discrete when the interval lies entirely above zero, continuous when it lies entirely below zero, and inconclusive when it contains zero. The holdout was evaluated exactly once per cohort. The component count was selected within the development sample only, using ten inner person-grouped folds nested inside each outer fold and choosing the count with the lowest mean inner test negative log-likelihood; the selected count and the selected continuous variant were then refitted on the full outer-training sample and scored on the outer-test sample, which took no part in selection. Because the selected count may differ between outer folds, the verdict is the margin rather than any single count. The reported component count was the modal interval-arm count across evaluable outer folds and was used for the full-development refit scored on the locked holdout.

Fold counts. Twenty-four of the twenty-five waves ran at 100 outer person-grouped folds with 10 inner folds. The Danish BEAR cohort[14] ran at 50 outer folds with the same 10 inner folds, because it contributes 172 ears from 86 participants after bilateral-complete filtering and a person-grouped design cannot form more folds than it has participants. Fewer outer folds give a noisier margin and a wider standard error, so the BEAR verdict is not comparable to the other waves at equal statistical power and should be read as low-confidence in either direction. The cohort was retained rather than excluded because dropping it would bias the panel toward large surveys. Because model selection runs inside the 85% development sample, the number of outer folds that can carry test ears is bounded by the development-sample participant count rather than by the cohort participant count. In the smallest cohorts the frozen fold count exceeds that bound, and the excess folds contain no held-out ears. Such folds are structurally empty: no per-ear margin is defined for them, they are excluded from the fold-level descriptive summaries, and the number excluded is reported for every affected cohort. They are never imputed, and never counted as ties or as fold-level wins for either model.

Minimum group size. A recovered subtype was required to contain at least the square root of the development-sample size, applied in ears: the threshold is one over the square root of the number of ear rows in the development sample of the fold in which the fit is scored, not the number of participants. Folds are formed by participant, but the floor is evaluated on the ear count.

Admissibility and the three non-discrete outcomes. The degeneracy rules are admissibility rules, applied before any comparison and independently of which model they favour: a component count is admissible on an outer fold only if no inner fold produced a degenerate component, whether by vanishing weight or by a near-singular covariance, and an inadmissible fit is discarded. Three outcomes must be distinguished and are reported separately. A cohort is continuous when both families were evaluable and the holdout interval lies below zero. A cohort is inconclusive when both families were evaluable and the holdout interval contains zero. A cohort is not evaluable when the prespecified discrete family had no admissible candidate on any outer fold; in that case no margin exists, no verdict is assigned, and the cohort is excluded from the discrete-versus-continuous tally rather than counted as continuous. Not-evaluable outcomes arise in the smallest cohorts, where the weight floor scales as one over the square root of the development-sample ear count and no mixture component can satisfy it. Reporting such a cohort as continuous would let an admissibility rule masquerade as evidence about hearing.

Uncertainty for the discreteness test. Inference is based on the locked holdout. The interval is a 95% percentile interval from 10,000 bootstrap resamples of holdout participants, resampling participants with replacement and carrying both ears of a participant together; this is valid because the holdout scores come from one fixed pair of fitted models and therefore carry no dependence on model fitting. Participant counts are reported alongside every interval. The outer-fold margins from the nested cross-validation are retained as descriptive summaries of model-selection stability and are reported as a mean and a standard error across folds; they are not used inferentially. Fold-level paired t statistics and exact binomial sign tests were not used for inference because overlapping training samples make outer folds statistically dependent and neither statistic is valid under that dependence. The 5,000 cohort-cluster bootstrap replicates were used only to estimate uncertainty in the severity maximum.

All models used the complete set of thresholds for each ear. Pure-tone average was used only to group ears by severity and as a comparator, never as a model input.

#### S2. Why a recovered component count is not a phenotype count

Statistical models used to estimate subtype counts can divide even a homogeneous population into multiple apparent groups, particularly as the dataset grows. Two distinct formal results support that statement, and they fail in different ways.

Dirichlet-process mixtures. Miller and Harrison give the sharpest form. In the simplest possible setting, a Dirichlet-process mixture with normal components of unit variance applied to data generated from a single standard normal component, the posterior probability that there is one cluster converges in probability to zero rather than to one. The model does not merely estimate the count imprecisely; it becomes certain of the wrong answer as the sample grows, because the process prefers to introduce additional small components that the data do not require.

Finite mixtures. Cai, Campbell and Broderick address the other family. Consistency of the component-count posterior requires the component likelihoods to be correctly specified. Under even the slightest misspecification the posterior diverges: the posterior probability of any particular finite number of components converges to zero in the limit of infinite data. Because misspecification is effectively unavoidable with real data, inferred counts can change substantially with the size of the dataset.

The two results identify distinct and complementary failure modes. A Dirichlet-process mixture can invent groups where the population is homogeneous; a finite mixture can fail to settle on any count at all once the component family is even slightly wrong. These results do not imply that clustering is uninformative; they establish specific limitations of component-count inference. What our data add is the demonstration that audiogram subtype counts behave in exactly this way: the recovered count changed under resampling of the same population. Holding population, frequency grid and model family fixed, component selection was repeated independently within each of 100 participant-grouped outer folds, with any two fold-specific training samples differing in approximately 2.0% of participant membership. In all 29 analyses with a discrete locked-holdout verdict, the selected count varied across folds (median six distinct values, range 2–10; observed counts 2–12). The reported final count was selected in a median of 43% of folds, and 1,569 of 2,884 evaluable fold fits (54.4%) selected a different count.

Accordingly, the primary outcome was whether a cohort contained distinct groups, rather than the component count returned by an individual fit.

The same instability was tested at the level of individual ears rather than component counts alone. For each of the 29 analyses, the discrete model selected in each evaluable outer fold was refitted on that fold's training partition and applied, together with a taxonomy fitted on the complete development set, to the same locked-holdout ears never used in fitting or selection. Partition agreement used the adjusted Rand index (ARI), which is invariant to component relabelling and remains defined when fold and full-development solutions differ in component count; ear-level agreement used a label-invariant statistic comparing each ear's co-assigned holdout neighbours across taxonomies (mean Jaccard 0.49 against the full-development taxonomy). A matched concordance (0.68) is reported only as a secondary, matching-dependent descriptive statistic. Taking every pair of fold-derived taxonomies within a cohort, mean pairwise ARI across all such pairs, pooled over the 29 analyses, was 0.50 (range by cohort 0.29–0.77). Of these fold-taxonomy pairs, 32% had ARI below 0.5 while differing by under 0.02 nats/ear in interval predictive score on the identical holdout, scored under one shared quadrature draw.

#### S3. Contribution of ears to the Atlas decomposition

The canonical Atlas coordinates were derived from the 89,752 ears recorded under the ISO/ANSI reference. This single frozen construction supplies every reconstruction, transfer, longitudinal and acute result reported in this work and is applied without refitting throughout. The 27,712 ASA-era ears contributed nothing to it and were reserved for external historical validation. A decomposition of the full six-frequency corpus of 117,464 ears, formed by adding those ears after conversion to the ANSI-1969 reference, is reported as a sensitivity analysis where stated; it recovers the same axes and changes five-coordinate reconstruction of longitudinal change from 1.0148 to 1.0036 dB.

Level extraction. HAC1 is defined as the mean threshold across an ear's measured frequencies and is extracted rather than fitted. It is separated before the shape coordinates are derived, so that the shape coordinates describe variation in audiogram shape independently of overall level and are not dominated by the population mean or by the arbitrary decibel origin.

Derivation. The canonical Atlas was fitted using all 89,752 ISO/ANSI-referenced ears: 46,184 ears from the US post-1999 waves and 43,568 ears from KNHANES 2009-2012. The 27,712 ASA-era ears contributed nothing to it and were reserved for historical validation. The frozen coordinate frame used throughout the reported analyses was derived from this corpus. For held-out reconstruction and preservation analyses, the coordinate frame was re-derived within each participant-level development sample and evaluated only in the corresponding held-out participants, as described in S5. The reported mean absolute error of 1.35 dB, the proportions of thresholds and complete audiograms reconstructed within 5 dB, and the preservation of 99.93% of clinically distinguishable audiogram pairs were obtained from these split-specific development frames.

Contribution of ears. Every eligible ear contributed once, at ear level rather than as a catalogue of distinct threshold patterns. Age-stratified derivatives and pooled supersets were excluded to prevent duplicate contribution.

Duplicate handling. Repeated threshold patterns were retained when fitting both coordinate constructions. Deduplication was used only for distance-based dimensionality analyses, where identical vectors contain no information about separation.

### S4. Correlation-dimension implementation

The estimator determines how rapidly the number of similar audiograms increases as the permitted difference between them is widened. Formally, the correlation integral counts the proportion of audiogram pairs separated by less than a given distance, and the correlation dimension is the slope of that proportion against distance on logarithmic axes.

Distances are Euclidean in decibels across the frequencies a cohort measured. Because the estimate cannot exceed the number of frequencies tested, cohorts were compared only against cohorts measured on the same frequencies, and age comparisons paired senior and working-age groups drawn from the same survey wave.

Fitting range and diagnostics. The correlation integral was evaluated over 40 logarithmically spaced radii spanning 1-150 dB, with the slope fitted by ordinary least squares over the prespecified 9-28 dB window. A radius contributed to the fit when it lay within that window and its cumulative pair count was non-zero, and at least five such radii were required. The pair-count floor of 200 and saturation ceiling of 0.60 were used only in the separate synthetic procedure that selected the fitting window and were not applied to individual cohort estimates. The window rule itself was fixed against a synthetic battery of known dimension before any cohort was analysed, and never by inspecting which window preserved a cohort result: a window is admissible only if its median bias is at most 0.35 and its 90th-percentile absolute bias at most 0.75 on that battery, and every cohort estimate carried forward must additionally meet a log-log fit R-squared of at least 0.99. A local-slope dispersion diagnostic was also computed but was not used as an inclusion criterion, because it does not distinguish a scaling region from a saturating correlation integral.

The deduplicated analysis included 65 cohorts and 284 cohort-band observations meeting the fit criterion and placed the vertex at 43.2003 dB, with a 95% confidence interval of 41.3532 to 44.7152 from 5,000 cohort-cluster bootstrap replicates.

Inclusion criteria. A cohort-band estimate entered the primary severity analysis only if it satisfied all of the following conditions: at least 150 observations in the band, at least five non-zero correlation-integral points inside the 9-28 dB fitting interval, and a log-log fit R-squared of at least 0.99. The local-slope flatness criterion of 0.45 was not applied to the primary analysis.

#### S5. Held-out reconstruction and preservation of audiogram distinctions.

The reconstruction results were evaluated in participants who contributed nothing to coordinate derivation. The 89,752-ear ISO/ANSI corpus was divided at the participant level into a 75% development sample of 67,314 ears and a 25% held-out sample of 22,438 ears from 11,219 participants, with both ears of each participant kept on the same side of the split.

Repeated validation. The procedure was repeated across five fixed participant-level splits. The primary split generated the reported reconstruction values, and the remaining four assessed stability. Mean absolute error across the five splits ranged from 1.3434 to 1.3526 dB.

Reconstruction. HAC1 was the mean threshold across the six measured frequencies. After overall level was separated, an ear was represented by the first four shape coordinates, HAC2-HAC5, derived exclusively from the development sample; the audiogram was then reconstructed from those four coordinates together with HAC1.

$$\hat{X} = (X - HAC1)B \quad B^{T} + HAC1,$$

where B contains the four retained shape-loading vectors. The five-coordinate representation therefore comprises overall hearing level and four shape coordinates.

Reconstruction outcomes. Mean absolute error was calculated across all reconstructed thresholds in the held-out sample. Threshold-level accuracy was the proportion of individual thresholds reconstructed within 5 dB. Whole-ear accuracy required all six reconstructed thresholds of an ear to fall within 5 dB of their recorded values. In the primary split, the five-coordinate representation produced a mean absolute error of 1.3495 dB, reconstructed 95.662% of individual thresholds within 5 dB and reconstructed all six frequencies within 5 dB in 81.358% of ears.

Preservation of audiogram distinctions. Two audiograms were defined as clinically distinguishable when they differed by more than 5 dB at one or more measured frequencies. Two million pairs were sampled independently and with replacement from the held-out ears of the primary split. Sampling ears rather than unique threshold patterns made the draw proportional to population frequency rather than to the catalogue of distinct shapes. Self-pairs and pairs that were not clinically distinguishable before reconstruction were excluded.

For the primary split, 1,967,953 sampled pairs met the eligibility definition. Each pair was reconstructed from HAC1-HAC5 and counted as merged when the reconstructed audiograms no longer differed by more than 5 dB at any frequency. Of the eligible pairs, 1,398 were merged, giving preservation of 99.9290% of clinically distinguishable audiogram pairs.

#### S6. Duplicate audiograms under 5 dB recording

Clinical pure-tone thresholds are ordinarily measured and reported on a 5 dB grid, so a recorded threshold represents an interval of possible underlying values rather than an exact one. Different underlying thresholds are therefore recorded as identical audiograms, and at low severity a large number of ears share exactly the same set of recorded thresholds.

Identical audiograms carry no information about the separation between different audiogram shapes. Every distance-based estimate in this study was therefore computed twice: once on the recorded data and once with repeated threshold patterns counted only once. The proportion of exact duplicates was calculated for every cohort. The steep quantisation-driven rise in apparent dimensionality did not survive this correction, although the deduplicated analysis retained a non-linear severity trajectory that peaked in moderate hearing loss.

Finite-precision results from dynamical systems do not apply to this analysis because an audiogram population is a static point cloud rather than an iterated trajectory. The relevant support is the audiometric literature establishing the 5 dB step and its interval interpretation, together with the duplicate fractions measured here.

Recommendation for the field. Analyses of audiogram clustering, density or dimensionality should report exact-duplicate fractions and demonstrate that their findings persist once repeated threshold patterns are accounted for. Where the analysis compares competing density models, the recording interval should additionally be carried in the likelihood itself, as described in S1, rather than handled only by deduplication: a model comparison conducted on point densities over lattice-recorded data can be decided by how each family distributes mass at lattice points rather than by the structure under test.

#### S7. Age and severity in the dimensionality catalogue

Senior and working-age subsets drawn from the same survey wave were compared descriptively because they share equipment, calibration, measured frequencies, recruitment and study population. This comparison does not hold hearing severity constant. That distinction is important because correlation dimension itself varies non-linearly with hearing level, increasing toward moderate hearing loss and declining again at greater loss.

The dimensionality catalogue therefore permits an unadjusted within-wave age comparison but does not identify an independent effect of age. No age effect independent of hearing severity is inferred from these comparisons.

#### S8. Frozen-frame transfer: the portability protocol

Portability was evaluated by applying coordinate frames to independent populations that contributed nothing to their derivation. A decomposition of a single corpus may reflect corpus-specific structure rather than transferable audiogram geometry. Transfer to independent populations determines whether the axes represent recurring human audiogram variation rather than corpus-specific structure.

Why congruence alone is insufficient. Tucker's congruence coefficient states that two sets of loadings point in similar directions. It does not state what a clinician loses by using one frame in place of the other, it has no units, and it saturates: values above 0.95 are routinely obtained between frames that still differ measurably in reconstruction. Congruence describes geometric similarity but does not quantify portability.

The penalty. Portability was quantified as a transfer penalty in decibels. A donor frame is fitted in one population, frozen, and applied without any refitting, rescaling or per-coordinate adjustment to a receiving population that contributed nothing to its derivation. A reference frame is fitted locally within the receiving population under an identical protocol. The penalty is the receiving-population reconstruction error under the frozen donor frame minus the error under the locally fitted frame. It is bounded below by zero only in expectation; a small negative penalty is possible by sampling variation and is reported as such rather than truncated.

Reading a penalty. Clinical thresholds are recorded in 5 dB steps, so a penalty is interpreted against that resolution. An interval spanning zero was interpreted as indicating no detectable difference. An interval spanning zero indicates no detectable difference at the present sample size, not exact equality between frames.

Directions run. US to Korea and Korea to US on the full-corpus frames; the ISO/ANSI-restricted US frame to Korea; the canonical frame to the external German, Danish, Turkish, UK, Australian and Iranian cohorts[9,14,17,18,19,20]; and the canonical frame to the external ASA-era ears. Transfer was completed in both ISO-restricted directions: the US-ISO frame applied to held-out Korean ears carried a penalty of 0.0059 dB (95% CI -0.0031 to 0.0148), and the Korean frame applied to held-out US-ISO ears a penalty of 0.0519 dB (95% CI 0.0431 to 0.0607). The asymmetry concerned mean absolute error, not whether the frame transferred.

Interval method and bounds. Intervals are 95 per cent percentile intervals from 2,000 participant-grouped bootstrap resamples of the paired per-ear difference, resampling participants with replacement and carrying both ears of a participant together. The following bounds were obtained from the full-corpus four-country frame and are distinct from the canonical ISO/ANSI matched-frame penalties. On the six-frequency canonical grid with five coordinates, US to Korea gave a penalty of 0.0340 dB (95% CI 0.0220 to 0.0453) on 10,892 target ears from 5,446 participants, and Korea to US 0.0348 dB (0.0271 to 0.0436) on 18,474 ears from 9,237 participants. Both receiving populations contributed to the canonical pool. On the reduced 0.5, 1, 2 and 4 kHz grid, the external German cohort took a penalty of 0.3116 dB (0.1376 to 0.4838) with two coordinates and -0.0343 dB (-0.1214 to 0.0546) with three, on 284 ears from 142 participants. The penalty ceased to be detectable only after the second reduced-grid shape coordinate was added, and the negative point estimate at three coordinates was retained without truncation. Neither external cohort ever entered the canonical pool.

Four external cohorts are reported without a transfer penalty because fitting a local comparator frame within a development split and scoring it on the remaining participants leaves too few held-out ears to estimate a paired per-ear difference under the protocol applied to every other cohort, and no Danish transfer penalty is therefore estimated. Absolute zero-shot reconstruction in Denmark is reported below; it does not require a paired per-ear comparison and is therefore unaffected.

Applied at full scale in Germany, where a local frame can be fitted, the frozen coordinates were not merely acceptable but preferred: against a comparator fitted in all 569 German participants and scored on 570 held-out ears, the transfer penalty was -0.1158 dB. The corresponding figure in the Turkish cohort was -0.1326 dB on 324 held-out ears. Negative penalties of this size indicate that local refitting on a cohort of a few hundred participants overfits relative to the frozen frame, not that the frozen frame is superior in expectation.

What transfer does not establish. Global transfer was evaluated across eight countries. These analyses do not establish portability to populations, audiometric protocols or frequency grids not represented among the evaluated datasets.

Global external transfer. The transfer matrix described in this section is one component of the validation programme and not its whole extent: the occupational archive and the Iranian clinical cohort were evaluated outside it and are included in the external evaluations tabulated in Supplementary Table S3. Within the matrix, portability was evaluated across 17 cohort or survey evaluations, of which ten contributed no ears to the derivation and seven are held-out evaluations within the NHANES and KNHANES derivation families. Each receiving dataset was encoded using the frozen canonical Atlas without local modification whenever its measured frequency grid supported the required coordinates. Eleven evaluations measured the complete canonical six-frequency grid, with frozen reconstruction errors from 1.0869 dB in the 1963-1965 US wave to 1.5709 dB in the UK Cadenza panel. Six measured reduced grids and reconstructed the frequencies they did record to between 0.9755 and 1.8552 dB; these reduced-grid values are not comparable with the full-grid values and are never averaged with them. Fourteen evaluations supported a locally fitted comparator and yielded a transfer penalty; three cohorts of 172, 106 and 121 units respectively were evaluated zero-shot only, and are reported without a penalty rather than with one estimated from a sample too small to support it. Coordinates requiring frequencies absent from a receiving protocol were treated as unavailable rather than imputed, as described in S9.

Large-scale occupational validation. The largest receiving dataset was the US National Institute for Occupational Safety and Health longitudinal audiometry archive[8]. After quality control it contained 4,092,824 ear-examinations from 2,046,412 examinations in 1,365,737 workers between 1981 and 2010. The canonical Atlas was applied zero-shot: no loading, scaling parameter or coordinate definition was re-estimated from NIOSH before reconstruction. With HAC1-HAC5, the mean absolute reconstruction error was 1.018 dB and 98.19% of recorded thresholds were reconstructed within 5 dB.

Era stability. The same frozen encoding was evaluated separately for examinations recorded in 1981-1989, 1990-1999 and 2000-2010. The corresponding mean absolute errors were 1.0003,

1.0035 and 1.0232 dB. Worker counts in these era-specific analyses are not additive because a worker may contribute examinations to more than one era.

Local-refit comparison in NIOSH. To determine how much population-specific refitting could improve reconstruction, a coordinate frame was fitted directly within NIOSH and compared with the frozen Atlas on matched target audiograms. Mean absolute error was 0.8064 dB for the local frame and 0.9963 dB for the frozen frame, a difference of 0.1900 dB (95% CI 0.1891 to 0.1909). Because the standard clinical threshold grid is 5 dB, this improvement corresponds to approximately 3.8% of one recording step. The local frame was closely congruent with, but not identical to, the canonical Atlas: Tucker congruence after optimal matching was 0.9897, 0.9800, 0.8998, 0.8477 and 0.9488, and the principal angles between the two shape subspaces were 0, $2.6 \times 10^{-8}$, $3.0 \times 10^{-8}$ and 0.3214 radians. Three of the four shape axes were therefore recovered to numerical identity by a frame fitted independently in 1,365,737 workers who contributed nothing to the derivation of the Atlas, and only the fourth axis differed, by 18.4 degrees.

#### S9. Observability: which coordinates a frequency grid can recover

Each shape coordinate is a contrast among particular frequencies, so a coordinate whose defining frequencies were not tested has not been measured.

The reduced decomposition. When a cohort measures fewer frequencies, a decomposition fitted on that grid returns its own shape coordinates. On the four-frequency grid these are the reduced coordinates R3 and R4. They are not the canonical HAC3 and HAC4 recorded under a different name, they are not interchangeable with them, and a four-frequency cohort is never assigned canonical HAC3 or HAC4 values.

The masking test. The separate question is which canonical coordinates a reduced grid can recover. This was tested in ears whose complete six-frequency audiograms were known by masking the audiogram to 0.5, 1, 2 and 4 kHz and estimating the complete-audiogram coordinates from those four thresholds using two independently specified linear estimators. Both were fitted exclusively in source-country development participants and evaluated in held-out participants, with both ears of each participant kept together. Model selection and tuning were performed entirely within the development data. The two estimators produced materially identical recoverability conclusions. Overall hearing level and the local 2 kHz contrast were recoverable in both populations. High-frequency slope met the 0.80 criterion in the United States and fell just below it in Korea; broad curvature and the local 3 kHz contrast did not meet the criterion in either population.

The recovery statistic and its threshold. Recovery was measured as variance reduction: one minus the ratio of residual variance of the coordinate estimated from the masked audiogram to its variance in the source population. A coordinate was considered reliably recoverable when variance reduction reached the prespecified threshold of 0.80. On US development ears applied to held-out US ears the values are 0.9688 for overall hearing level, 0.8035 for high-frequency slope, 0.2346 for broad curvature, 0.9048 for the local 2 kHz contrast and 0.1849 for the local 3 kHz contrast. The same ordering holds when the estimator is derived in Korea and applied to held-out Korean ears, at 0.9703, 0.7893, 0.2646, 0.8985 and 0.1750. Curvature and the 3 kHz contrast therefore fall below the criterion in both populations, and by a wide margin rather than marginally.

Consequence. The routine four-frequency audiogram is not a uniformly coarser version of the six-frequency audiogram. It is selectively blind, and the blindness is specific and predictable rather than diffuse. Curvature and the 3 kHz contrast were therefore treated as unavailable on four-frequency grids rather than estimated.

Recommendation for the field. Frequency selection has been governed largely by convention, test time and clinical practicality. A study can now select frequencies by the coordinates it needs to observe: a study of curvature or of the 3 kHz contrast must measure 3 and 6 kHz directly, and cannot recover either from the four-frequency grid however large its sample.

#### S10. Full-spectrum validation across normal and impaired hearing

The unchanged five-coordinate ISO/ANSI Atlas was evaluated across the complete normal-impaired range. Strict-normal hearing was defined as thresholds of 20 dB HL or better at all six canonical frequencies. The impaired group contained ears with at least one threshold above 20 dB HL. Participants, rather than individual ears, were used for all development-validation partitions and resampling procedures, with both ears of each participant always carried together.

Absolute reconstruction within normal hearing. The prespecified Atlas was applied without refitting to strict-normal ears in the held-out participant sample. Reconstruction accuracy was assessed as mean absolute error across thresholds, the proportion of individual thresholds reconstructed within 5 dB, and the proportion of complete six-frequency audiograms for which every threshold was reconstructed within 5 dB. Mean absolute error was 1.1986 dB (95% CI 1.1792-1.2178); 97.094% of individual thresholds and all six thresholds in 85.856% of ears were reconstructed within 5 dB.

Relative retention of audiogram-shape variation. Overall hearing level, HAC1, was removed from each audiogram before the comparison. Within the normal and impaired groups separately, retained shape variation was calculated as the proportion of variation in the de-levelled threshold vectors accounted for by HAC2-HAC5. The primary statistic was the difference between the retained proportion in normal and impaired hearing, calculated as normal minus impaired. Equivalence required the lower 95% confidence bound on this difference to exceed -5 percentage points. HAC2-HAC5 retained 87.882% of shape variation in normal-hearing ears and 97.078% in impaired ears, a difference of -9.196 percentage points with a lower 95% confidence bound of -9.499 percentage points. The equivalence criterion was therefore not met.

The retained-variation analysis was repeated across all prespecified participant-level stability analyses and under two descriptive sensitivity definitions. In the first sensitivity analysis, an ear was classified as normal-hearing when the maximum of its six air-conduction thresholds at 0.5, 1, 2, 3, 4 and 6 kHz was 25 dB HL or lower, and as impaired otherwise, with no ears excluded. In the buffered sensitivity analysis, an ear was classified as normal-hearing when that maximum was 15 dB HL or lower and as impaired when it was 25 dB HL or higher; ears whose maximum threshold fell between these values were excluded. Both sensitivity definitions used the same retained-variation statistic, participant-grouped resampling procedure and one-sided 95% lower confidence bound as the primary analysis. The equivalence criterion remained unmet under both definitions.

Continuity across clinical hearing-loss cutoffs. We tested whether reconstruction error changed abruptly at the conventional boundary between normal and impaired hearing. The primary cutoff was 20 dB HL, with 15 and 25 dB HL evaluated as prespecified additional cutoffs. Because thresholds are recorded on a discrete 5 dB lattice, the abrupt-change estimate was a prespecified adjacent-shell contrast rather than a regression discontinuity or piecewise-regression model. At each cutoff c, the estimate was the difference in mean per-ear HAC1-HAC5 reconstruction error between ears whose maximum threshold equalled c dB HL and ears whose maximum threshold equalled c + 5 dB HL. One-sided 95% upper confidence bounds were obtained from 2,000 participant-grouped bootstrap resamples drawn independently within the two shells, with both ears of each participant carried together. Acceptability required the upper confidence bound to remain below 0.5 dB. At 20 dB HL, the estimated abrupt change was -0.0047 dB and the upper 95% confidence bound was +0.0442 dB. The corresponding estimates were +0.0877 dB with an upper bound of +0.1297 dB at 15 dB HL and +0.0552 dB with an upper bound of +0.1152 dB at 25 dB HL. All three cutoffs met the prespecified criterion.

Transfer within normal hearing. Transfer was evaluated within strict-normal hearing in both directions between the United States and Korea and between the ISO/ANSI and ASA calibration eras. In each comparison, a coordinate representation derived in the donor population was applied without refitting to held-out target ears and compared on the same target ears with a representation fitted locally within the receiving population. The transfer penalty was the paired difference in mean absolute reconstruction error, calculated as donor-derived minus locally fitted error. Confidence intervals were obtained by participant-grouped resampling, carrying both ears of each participant together. Acceptability required the complete confidence interval to remain within ±0.5 dB.

The United States-to-Korea comparison included 4,332 target ears from 2,654 participants and produced a transfer penalty of +0.0779 dB. Korea-to-United States included 6,153 ears from

3,568 participants and produced a penalty of -0.0236 dB. ISO/ANSI-to-ASA included 4,866 ears from 2,841 participants and produced a penalty of -0.0592 dB. ASA-to-ISO/ANSI included 10,485 ears from 6,222 participants and produced a penalty of +0.0935 dB. Every confidence interval remained within the prespecified 0.5 dB acceptability limit.

Composite criterion. The primary full-spectrum conclusion required all three prespecified criteria to be met: equivalent relative retention of audiogram-shape variation in normal and impaired hearing, continuity across the clinical hearing-loss boundary, and acceptable transfer within normal hearing across populations and calibration eras. The continuity and transfer criteria were met. The relative-retention equivalence criterion was not met; the prespecified composite criterion was therefore not met. Absolute reconstruction within normal hearing was a supporting analysis and did not override the failed relative-retention criterion.

#### S11. NIOSH longitudinal cohort and edge construction

The occupational longitudinal analyses used audiograms from the US National Institute for Occupational Safety and Health hearing-conservation archive[8]. The source contained 2,944,670 examinations from 1,816,812 workers. After application of the frozen ingestion and quality-control rules, the analytic dataset contained 2,046,412 examinations from 1,365,737 workers, corresponding to 4,092,824 ear-examination rows.

Threshold values were processed according to the documented conventions of the source dataset using prespecified quality criteria applied consistently across analyses. Recorded values were not rounded or reinterpreted.

All primary Atlas encoding and longitudinal reconstruction in NIOSH used the six canonical frequencies 0.5, 1, 2, 3, 4 and 6 kHz. The 8-kHz threshold was excluded from coordinate construction and was reserved for the separate extended-frequency analyses.

For longitudinal analyses, examinations were ordered within worker and ear. Successive eligible examinations defined longitudinal edges. Each edge therefore represents the observed change between two measurements of the same ear in the same worker. Analyses requiring a minimum elapsed time applied that interval after edge construction; otherwise all successive eligible edges were retained.

The principal longitudinal reconstruction dataset contained 1,361,350 same-ear edges from 446,146 workers. Analyses requiring at least 0.5 years between examinations contained 1,358,368 edges from 444,820 workers. Analyses of the longitudinal covariance and geometry used 1,294,932 eligible edges from 421,868 workers after their corresponding prespecified completeness requirements were applied. These three edge sets are distinct and are named explicitly wherever a quantity is reported.

Follow-up structure. Of the 1,365,737 workers in the analytic dataset, 919,591 contributed one examination, 289,301 two, 100,167 three, 39,114 four, 14,122 five and 3,442 six. The interval between successive examinations had a median of 4.30 years and a mean of 4.32 years, with a maximum of 28.04 years. Examination dates ran from 1 January 1981 to 31 December 2010 with no unparseable or implausible dates, and no duplicate worker-date-ear key was present. Overall hearing level across the 4,092,824 ear-examinations had a mean of 14.25 dB, a median of 11.67 dB and a 95th percentile of 36.67 dB, so the archive is weighted toward mild loss and the longitudinal results should be read as characterising an occupational surveillance population rather than a clinic population.

#### S12. Reconstruction of longitudinal audiogram change

For two successive audiograms X and X from the same ear, the observed threshold change was $\Delta X = X - X$. Each audiogram was encoded using the unchanged canonical Atlas, producing coordinate vectors z and z. Coordinate change was $\Delta z = z - z$.

For each reconstruction depth, the observed threshold change was reconstructed from the corresponding coordinate differences using the same frozen basis used for cross-sectional audiogram reconstruction. No longitudinal coordinate system was fitted for this analysis. The test therefore asks whether coordinates derived from population audiograms also span the changes observed when an individual ear is measured repeatedly.

Using HAC1-HAC5, mean absolute error between the observed and reconstructed threshold change was 1.0148 dB (95% CI 1.0132 to 1.0164). Overall, 98.21% of reconstructed threshold changes were within 5 dB and 99.94% were within 10 dB. The HAC1-HAC5 reconstruction accounted for 95.32% of the observed variance in threshold change. Inclusion of HAC6 completes the six-frequency basis and reconstructs the six measured threshold changes to numerical precision.

Error by measured frequency. The residual of the five-coordinate representation is not distributed evenly across the audiogram, and the longitudinal profile closely follows the cross-sectional one for the same archive. Because HAC6 is defined as the local 1 kHz contrast, the cost of stopping at five coordinates is concentrated predominantly at 1 kHz, and the representation is effectively exact at the higher frequencies.

Error by follow-up interval and by baseline severity. Mean absolute error of the five-coordinate longitudinal reconstruction was 0.9428 dB for intervals under one year, 0.9544 at one to two years, 0.9846 at two to three, 1.0101 at three to five, 1.0494 at five to ten and 1.1872 at ten years or more. Stratified instead by the baseline overall hearing level of the ear, it was 0.9743 dB below 20 dB, 1.1106 at 20-34 dB, 1.2271 at 35-49 dB, 1.2924 at 50-64 dB, 1.3186 at 65-79 dB and 1.4810 at 80 dB or above. Accuracy therefore degrades gradually with both elapsed time and severity and remains below one third of the 5 dB recording step in every stratum, including the 143 edges beginning above 80 dB.

#### S13. Metric, level-versus-shape decomposition and longitudinal covariance

HAC1 is the arithmetic mean of the six thresholds, whereas HAC2-HAC6 are coefficients along unit-norm shape vectors. A one-unit change in HAC1 therefore changes all six threshold values by one decibel and cannot be compared directly with a one-unit change in a shape coordinate. Distances and variance partitions that compare level with shape consequently used the isometric coordinate vector

$$z = (\sqrt{6}\ \text{HAC1}, \text{HAC2}, \text{HAC3}, \text{HAC4}, \text{HAC5}, \text{HAC6}).$$

Under this transformation, squared Euclidean distance in coordinate space equals squared Euclidean distance between the corresponding six-frequency audiograms. Raw HAC1 values were retained for clinical reporting, but raw HAC1 variance was never compared directly with HAC2-HAC6 variance.

Among 1,358,368 longitudinal edges separated by at least 0.5 years, 42.84% of total squared displacement occurred along overall hearing level and 57.16% along the five shape coordinates. This partition is a geometric description of observed audiogram change and is not a decomposition into biological causes.

The raw longitudinal covariance matrix was estimated from coordinate differences across 1,358,368 eligible edges from 444,820 workers, the same minimum-interval edge set used for the level-versus-shape partition above. The marginal raw variances were 23.0505 for HAC1, 61.1381 for HAC2, 64.5776 for HAC3, 32.0079 for HAC4, 26.8612 for HAC5 and 15.8527 for HAC6, with worker-cluster bootstrap intervals of 22.85-23.26, 60.79-61.45, 64.34-64.80, 31.88-32.12, 26.75-26.99 and 15.80-15.90 respectively from 1,000 resamples. After the isometric transformation, the HAC1 variance is $6 \times 23.0505 = 138.303$. The eigenspectrum reported in S15 is derived from the separate 1,294,932-edge geometry set and is not interchangeable with the variances given here.

Total observed movement increased monotonically with the interval between examinations. The trace of the coordinate-difference covariance was 172.09 below one year, 182.98 at one to two years, 198.94 at two to three years, 214.23 at three to five years, 245.17 at five to ten years and 382.43 at ten years or more. Displacement in the Atlas metric therefore accumulates with elapsed time, as a distance in a state space should.

The prespecified measurement-error correction required at least 200 same-ear repeat pairs within 30 days. Only 44 such pairs were available, so no measurement-error covariance was subtracted. A 90-day sensitivity set contained 484 pairs but may contain genuine biological change and therefore cannot isolate measurement error. All reported longitudinal covariance results therefore describe observed within-ear change rather than noise-corrected biological change.

An independent bound on measurement variability was nevertheless available without repeat pairs. Regressing the squared per-frequency threshold difference on the elapsed interval across 1,342,404 edges and extrapolating to zero elapsed time gives a frequency-specific intercept of 28.45 $dB^2$ at 0.5 kHz, 20.54 at 1 kHz, 20.57 at 2 kHz, 23.07 at 3 kHz, 32.16 at 4 kHz and 67.30 at 6 kHz, with slopes of 1.98, 2.39, 4.67, 7.96, 10.27 and 10.27 $dB^2$ per year respectively. These intercepts bound the variance not attributable to elapsed time. They were not subtracted from any reported covariance, because the extrapolation assumes a linear variance-time relationship that the low coefficients of determination of these fits do not establish, and they are reported only as an order-of-magnitude bound.

### S14. Correspondence between population variation and within-ear change

The central longitudinal test asked whether the principal shape variations separating different ears are the same variations expressed when an individual ear changes. Overall hearing level was removed before this comparison so that correspondence could not be driven by severity alone.

Between-person shape covariance was estimated across 1,365,737 NIOSH workers, one qualifying audiogram per worker. Within-ear covariance was estimated independently from 1,294,932 longitudinal change vectors contributed by 421,868 workers. Both covariance matrices were expressed in the five-dimensional shape space HAC2-HAC6 and eigendecomposed independently.

Subspace correspondence was quantified from the principal angles between the leading eigenspaces. For two orthonormal basis matrices U and V, the singular values of $U^{T}V$ are the cosines of the principal angles. The primary overlap statistic was the mean squared canonical correlation between the leading two-dimensional subspaces, bounded from zero for orthogonal spaces to one for coincident spaces. Formally, for leading k-dimensional orthonormal bases U_k and V_k, O_k = (1/k)||U_k^T V_k||_F^2 = (1/k) sum_j cos^2(theta_j). The primary comparison used k = 2, giving the reported value of 0.920151. Longitudinal edges required at least one year between examinations.

The observed leading-subspace overlap was 0.920151, with a 95% worker-cluster bootstrap interval of 0.9184-0.9219. The first two principal angles were 0.04434 rad (2.54°) and 0.40842 rad (23.40°). Of the observed within-ear shape variance, 92.12% lay within HAC2-HAC5 and 7.88% along the remaining HAC6 direction.

A random-orientation null was generated by comparing the observed eigensystem with independently sampled orthogonal orientations of the same dimensionality. The null used 2,000 independently sampled random orthogonal orientations. The null overlap had a median of 0.403 and a 95th percentile of 0.663; the observed value was 0.920 (Monte Carlo p=0.0005). Bootstrap intervals were 95% intervals from 2,000 worker-cluster resamples. The corresponding maximum-principal-angle null likewise placed the observed alignment far outside random orientation.

A separate examination-order permutation was performed within worker. Reordering an individual's examinations changes the temporal direction of successive differences while preserving the collection of audiograms available to that worker. This permutation did not reduce the geometric correspondence. The permutation overlap median was 0.937, greater rather than smaller than the observed 0.920; the corresponding one-sided permutation test therefore provided no evidence for chronological-direction-specific correspondence. The analysis supports correspondence of the space in which hearing varies, but does not establish a universal chronological direction of progression.

Axis-by-axis comparisons were treated as secondary because independently estimated eigenvectors can exchange order when eigenvalues are similar. After optimal one-to-one matching, the congruence coefficients between independently derived within-ear and between-person shape axes were 0.849, 0.818, 0.845, 0.920 and 0.988. These values support the subspace result but are not used to define the primary conclusion.

### S15. Longitudinal complexity, severity and categorical transitions

The spectrum of observed longitudinal change was summarized from the eigenvalues of the covariance of Δz. Participation ratio was calculated as $(\Sigma\lambda_i)^2 / \Sigma\lambda_i^2$ and used as a descriptive effective-rank measure; it is distinct from the Grassberger-Procaccia correlation dimension used for the cross-sectional severity analysis in S4.

Estimated on the 1,294,932-edge geometry set from 421,868 workers, the five HAC2-HAC6 eigenvalues were 81.71, 50.70, 31.69, 23.36 and 14.95, corresponding to variance shares of 40.37%, 25.05%, 15.66%, 11.54% and 7.39%. The participation ratio was 3.718; four coordinates were required to retain 90% of observed shape-change variance and all five to retain 95%. Including the level coordinate increased the participation ratio to 4.274, with five coordinates required for 90% and six for 95%. Because no prespecified measurement-error covariance was available, these are descriptive properties of observed longitudinal change and are not interpreted as noise-corrected biological dimensionality.

The same quantity computed on subsets behaved consistently. Restricting to edges separated by one to three years gave a shape participation ratio of 3.665 and by three to ten years 3.719, so the estimate is not an artifact of a particular follow-up length. Splitting instead by baseline severity gave 3.663 for edges beginning below 35 dB, on 1,276,914 edges, against 4.163 for edges beginning at 35 dB or above, on 84,436 edges from 39,257 workers. This severity contrast is an independent observation of the same direction reported below, obtained on a different stratification and a different subset, and is descriptive rather than a test.

To examine variation with baseline severity, longitudinal edges were grouped according to the starting HAC1 value. Shape participation ratio was 3.505 below 10 dB, 3.634 at 10-19 dB, 3.836 at 20-29 dB, 4.061 at 30-39 dB, 4.185 at 40-49 dB, 4.126 at 50-59 dB, 3.800 at 60-69 dB and 3.502 at 70 dB or greater. The maximum therefore occurred in the 40-49 dB band; the median baseline HAC1 among observations in that band was 43.33 dB.

This longitudinal participation-ratio analysis and the cross-sectional correlation-dimension analysis in S4 estimate different quantities and were not pooled or treated as interchangeable estimators. Their similar location in severity was interpreted only as concordance of two independently defined observations.

Conventional configuration labels were assigned independently to each examination using label definitions locked before any result was inspected. Severity was graded from the four-frequency pure-tone average at 0.5, 1, 2 and 4 kHz with band edges at 20, 35, 50, 65, 80 and 95 dB. A configuration was called flat when thresholds varied by no more than 15 dB across the measured frequencies; a notch required a depth of at least 15 dB with a recovery of at least 10 dB. Among 1,361,350 successive longitudinal edges, 27.5006% changed at least one label. Median isometric Atlas displacement was 19.36 when a label changed and 13.23 when it did not. The small-movement region was defined in advance as an isometric Atlas displacement of 5.0 dB or less. Nevertheless, 3.68% of label changes occurred despite displacement remaining within that small-movement region, demonstrating that categorical transitions can occur by crossing a classification boundary without a correspondingly large change in the underlying continuous representation.

#### S16. State dependence of longitudinal hearing change

To test whether subsequent hearing change depended on current hearing state, the longitudinal Atlas was partitioned into prespecified rectangular cells of baseline coordinate space. Cells were defined on three coordinates: overall hearing level HAC1, with bin edges at 15, 25, 35, 45 and 60 dB, and the two leading shape coordinates HAC2 and HAC3, each with bin edges at -10, 0 and 10. Values falling on a boundary were assigned to the upper bin, and the outermost bins were left unbounded. No smoothing or interpolation across cells was permitted. Longitudinal edges required at least one year between examinations, and a cell contributed to the primary analysis only when it contained at least 200 eligible edges.

The analysis contained 1,294,932 edges from 421,868 workers. Ninety-five state cells were defined and 61 met the minimum-count criterion. For each evaluable cell, the distribution of subsequent change vectors was estimated independently. The between-cell share of subsequent-change variance was then calculated as the proportion of total change variance attributable to differences between starting-state cells.

Starting state accounted for 0.0491 of subsequent-change variance, with a 95% worker-cluster bootstrap interval of 0.0487-0.0496. Bootstrap intervals were 95% intervals from 1,000 worker-cluster resamples. Statistical significance was assessed by permuting the association between starting state and subsequent movement while preserving the longitudinal observations, using 1,000 permutations with the worker as the permutation unit, giving $p=0.000999$.

The result is therefore evidence of modest state dependence rather than deterministic flow. Approximately 95% of variation in subsequent change remains unexplained by the current Atlas cell, and no individual-level trajectory or prognosis is inferred from this analysis.

#### S17. Acute Ménière trajectories in Atlas space

The acute analysis comprised 356 unilateral-ear series[11,12] with audiograms recorded at five successive protocol timepoints labelled hours 0, 1, 2, 3 and 4. A series was eligible when it contained at least two observations, and thresholds were accepted between -10 and 120 dB. Hours 0-3 contained 356 series and hour 4 contained 354. The frozen canonical Atlas was applied without refitting.

For each timepoint and coordinate, the group mean was calculated across the available unilateral-ear series. Across the five timepoints the group mean moved simultaneously in overall hearing level and high-frequency slope, rather than as a uniform shift of the whole audiogram.

From hour 0 to the end of follow-up, mean HAC1 changed by -2.818 dB (95% CI -3.359 to -2.313), HAC2 by +3.714 (2.761 to 4.671) and HAC6 by +0.779 (0.225 to 1.339). Intervals for HAC3-HAC5 included zero. The modal time of maximum displacement was hour 3.

Trajectory displacement was calculated in the same isometric coordinate metric used for the longitudinal NIOSH analyses. Net displacement was 15.72 Atlas units and peak displacement 18.91. The variance shares across HAC1-HAC6 were 45.7%, 22.0%, 13.4%, 6.9%, 5.9% and 6.2%, respectively. The leading acute shape direction was dominated by HAC2, with loading approximately 0.952.

These statistics describe the group-level trajectory of the measured acute series. They do not imply that every individual Ménière episode follows the mean path or that the observed path identifies a unique underlying cochlear lesion.

#### S18. Cross-timescale comparison of acute and chronic hearing change

To compare acute hearing change over hours with chronic change over years, overall hearing level was removed from both datasets and the dominant shape-change eigenspaces were derived independently. The acute eigensystem was estimated from the Ménière series described in S17 and the chronic eigensystem from the NIOSH longitudinal changes described in S11-S15.

The primary subspace-overlap statistic was calculated using the same principal-angle construction as in S14. Acute and chronic shape spaces had an overlap of 0.80395. The first two principal angles were 0.20386 rad (11.68°) and 0.63422 rad (36.34°).

Against randomly oriented shape spaces of the same dimensionality, the overlap null had a median of 0.4094 and a 95th percentile of 0.6710. The observed overlap was greater than this null distribution (Monte Carlo $p=0.0105$). The leading-axis congruence was 0.743; secondary one-to-one axis congruences were treated descriptively because axis order is not stable when neighboring eigenvalues are similar.

The analysis therefore tests whether the two timescales occupy significantly shared directions of audiogram-shape change. It does not test whether acute and chronic trajectories are identical, whether their biological causes are the same, or whether they proceed in the same temporal direction.

To test whether the chronic eigenspace predicts acute change without adaptation, the frozen NIOSH chronic shape eigenspace (S11-S15) was applied unchanged to 1,422 successive hourly transitions from the 356 Ménière series (356, 356, 356 and 354 transitions across the four successive one-hour intervals). Variance captured by the frozen frame ($R_N = 0.55018$) was compared with the variance captured by a rank-2 frame fitted directly to the acute transitions ($R_M = 0.58205$, an in-sample geometric ceiling rather than an unbiased or holdout comparator), giving a transfer efficiency $R_N/R_M$ of 0.94525, that is 94.53% of the locally achievable rank-2 variance capture. Reconstruction of shape-change after overall-level removal gave a mean absolute error of 1.793 dB for the frozen frame against 1.790 dB for the locally fitted ceiling and 2.874 dB for a zero-change predictor.

Significance was assessed against 2,000 random rank-2 orientations generated by QR decomposition of 5 by 2 matrices of independent standard normal values, separately for variance captured and reconstruction error. The null median variance captured was 0.3996 (95th percentile 0.4843) against the observed 0.5502 (P = 0.0015); the null median reconstruction error was 2.186 dB (5th percentile 2.011 dB) against the observed 1.793 dB (P = 0.0010). Neither value reached the Monte Carlo resolution floor (1/2001 = 0.0005).

Stability under resampling was assessed by 2,000 series-level bootstrap replicates of the overlap statistic (median 0.7968, 95% CI 0.7359-0.8586). Because the source publication reports 347 patients against 356 identifiable ear-series, with patient-level linkage unavailable in the deposited data, a sensitivity analysis resampled 347 series per replicate; the result was materially unchanged (median 0.7982, 95% CI 0.7282-0.8632). Splitting the acute cohort into two independent halves of 178 series gave a half-to-half overlap of 0.9621 (95% CI 0.9002-0.9899) and half-to-NIOSH overlaps of 0.7964 and 0.7995, both closely tracking the full-cohort estimate.

The frozen-frame overlap was evaluated separately within each of the four successive one-hour intervals: 0.8617 (hour 0-1), 0.8687 (hour 1-2), 0.8499 (hour 2-3) and 0.9283 (hour 3-4). These per-interval values are descriptive and are not four independent tests; they are read against the frozen acute-to-chronic random-orientation null 95th percentile of 0.6710 reported above, used here only as a fixed reference point. All four point estimates lay above that reference. The bootstrap lower bounds for the first three intervals were 0.748, 0.720 and 0.715, whereas the fourth interval was wider (0.4814 to 0.9716); the per-interval estimates are therefore reported descriptively as consistent in magnitude rather than as four independent tests of significance.

As a descriptive diagnostic only, not a primary endpoint, the acute successive-transition eigenspace used in this transfer analysis overlapped the acute peak-displacement eigenspace of the primary comparison above at 0.97747, and overlapped the frozen chronic frame at 0.87805 (95% CI 0.8027-0.9383). These figures relate the two acute representations and do not alter the interpretation of the primary overlap reported above.

356 ear-series are analysed against 347 patients reported in the source publication; patient-level linkage is unavailable in the deposited data, so at most 9 of 356 series (2.53%) may share an unidentified second-ear counterpart, and the 347-series sensitivity analysis above addresses resampled-unit count rather than reconstructing true bilateral clustering. Two series lack an hour-4 observation. The chronic covariance used for the frozen frame is uncorrected for measurement noise. Rank k = 2 was fixed a priori and never searched.

#### S19. Robustness of the intermediate-severity dimensionality maximum

The severity maximum reported in S4 was re-evaluated using the same historical Grassberger-Procaccia estimator and the same quadratic cohort-adjusted severity model. The purpose was to distinguish robustness of the intermediate-severity pattern from portability of an exact numerical peak.

Reproduction of the correlation-dimension estimator. Correlation dimension was calculated directly from each cohort's native threshold vector, with no coordinate transformation or standardisation. Euclidean distance was measured in raw decibels on the frequencies available in that cohort. The correlation integral was evaluated at 40 logarithmically spaced radii from 1 to 150 dB and the slope fitted over the fixed 9-28 dB interval. A band required at least 150 observations, at least five non-zero correlation-integral points in the fitting interval and $R^2 \geq 0.99$. Severity was the arithmetic mean of the frequencies present in that cohort. Ten-decibel severity bands were used; the open 90-dB-and-greater band was excluded from the quadratic fit because it has no finite midpoint.

The severity model was $D = \alpha c + \beta 1S + \beta 2S^2$, where $\alpha c$ is a cohort-specific intercept and S the severity-band midpoint. Computationally this was fitted after within-cohort demeaning. The vertex was $-\beta 1/(2\beta 2)$ when $\beta 2 < 0$. Uncertainty was obtained by cohort-level resampling.

External native-grid analysis. The same estimator was then applied to independent datasets using each cohort's native frequency axis. Because correlation dimension is bounded by embedding dimension, cohorts with equal numbers of tones but different frequency sets were not automatically pooled. Grid-stratified analyses were reported only when sufficient admissible observations existed and sparse incompatible grids were not rescued by post hoc pooling.

The pooled external analysis contained 24 cohorts, 104 admissible cohort-band cells and 183,706 ears from 91,853 participants, together with 1,778 Münster ear-series observations[11]. Its vertex was 44.6474 dB (95% CI 43.0794-46.1342).

Three native grids contained sufficient information for a separate quadratic estimate: 500|1000|2000|4000|8000 Hz, vertex 44.1168 dB (42.4353-44.8071); 500|1000|2000|3000|4000|6000 Hz, 46.7383 dB (44.2096-48.7951); and 500|1000|2000|3000|4000|6000|8000 Hz, 43.1501 dB (39.4530-46.1204). All evaluable grid-specific fits were concave with an interior maximum. Other prespecified grid strata were retained as not evaluable when they contained too few observations or no admissible severity bands.

The grid-stratified external analysis and the grid-by-severity curvature diagnostic were separate calculations. Three native grids contained sufficient observations for standalone external quadratic estimates. The curvature diagnostic, applied to the available frozen band estimates, was evaluable in four grids and found negative curvature with an interior maximum in all four. Grid-specific curvature was heterogeneous. A model allowing grid-specific severity curvature was preferred to one imposing a common curvature after cohort fixed effects ($F=27.93$ on 9 and 208 degrees of freedom, $p=2.2\times10^{-31}$). In addition, the mean embedding dimension of admissible observations decreased with severity, from approximately 5.95 at 10-20 dB to 4.00 at 80-90 dB ($r=-0.8025$). These findings indicate that the intermediate-severity maximum is reproducible while its exact location is influenced by the audiometric frequency grid and should not be interpreted as a universal constant.

Local-slope flatness diagnostic. Among the 284 historically admissible bands, local-slope flatness ranged from 0.0714 to 0.4048, so no band was additionally excluded by the later 0.45 criterion. Two other evaluable bands exceeded 0.45 but had already failed the historical R-squared criterion.

Within the single fixed seven-frequency NIOSH grid, dimensionality showed the same non-monotonic severity pattern. At N=100,000 workers, correlation dimension was 4.155, 4.800, 5.066, 5.037, 4.930 and 4.913 across severity-band midpoints of 15, 25, 35, 45, 55 and 65 dB, respectively, with the empirical maximum at 35 dB. Quadratic fits at the two evaluable upper sample-size rungs were concave, but their fitted vertices shifted from 44.31 to 46.93 dB as sample size doubled. The within-grid analysis therefore supports an intermediate-severity maximum but does not identify a precise universal peak location.

Quantisation sensitivity. Frequency-specific lattice pitch and phase were estimated for each dataset rather than assumed globally. The historical ASA waves used 5 dB pitch with frequency-specific offsets, while Macquarie[19] used a 2.5 dB lattice and the remaining principal datasets were recorded on the conventional 5 dB lattice. Correcting the ASA lattice phase changed pairwise distance geometry by zero to numerical precision and shifted the pooled severity vertex by only 0.011 dB. A separate deterministic dequantisation-jitter analysis moved the pooled vertex by 2.76 dB, while grid-specific vertices moved by approximately 1 dB or less, further supporting interpretation of the exact pooled location as protocol-dependent rather than as a fixed biological threshold.

#### S20. Directional coherence of successive within-ear change

The covariance analyses in S13 to S15 describe how much an ear moves and along which directions, but not whether successive movements of the same ear point the same way. A representation in which hearing change accumulated in a consistent direction would behave differently from one in which change partly reverses, and the distinction matters clinically because it determines whether a single observed interval can be extrapolated.

Construction. For each ear with at least three eligible examinations, successive change vectors were formed as in S12 and consecutive pairs of those vectors were compared. The statistic is the cosine of the angle between one change vector and the next change vector of the same ear. A positive mean cosine indicates persistent drift, zero indicates directionless movement and a negative mean indicates reversal. The statistic was computed both in the full isometric coordinate metric and with overall hearing level removed, because a shared level drift alone can create apparent coherence that has nothing to do with audiogram shape.

Result. The analysis contained 468,712 successive step pairs from 156,728 workers. The mean cosine was -0.2195 in the full isometric metric (95% CI -0.2211 to -0.2179) and -0.2957 with overall hearing level removed (95% CI -0.2973 to -0.2942). Successive movements of the same ear are therefore negatively correlated, and the reversal is stronger in audiogram shape than in overall level. Nulls comprised cross-worker step permutation, random sign flipping and time reversal.

Interpretation and limits. Observed reversal of this magnitude is consistent with two contributions that this analysis does not separate: genuine physiological fluctuation of threshold around a slowly moving position, and measurement variability, which is negatively autocorrelated in successive differences by construction because adjacent change vectors share an examination. Because no measurement-error covariance was available (S13), the observed coherence is reported as a property of serial audiometry as recorded rather than as an estimate of biological reversal. The claim supported is the conservative one: successive recorded changes do not accumulate in a consistent direction, so a single interval of change is not a trajectory.

#### S21. Generalization to frequencies outside the canonical grid

The canonical Atlas is defined on 0.5, 1, 2, 3, 4 and 6 kHz. Many protocols additionally record 0.125, 0.25, 1.5 or 8 kHz. We asked whether the canonical coordinates carry information about those frequencies, which is a different question from the observability analysis in S9: there we asked which canonical coordinates a reduced grid can recover, here we ask whether the canonical coordinates predict a threshold that was never part of their construction.

Design. For each cohort measuring an extra frequency, that threshold was regressed on the Atlas coordinates of the same ear, fitted in a development split and evaluated in held-out participants with both ears of a participant kept together. The Atlas was never refitted. The comparator was a prespecified log-frequency baseline computed from the neighbouring measured thresholds: interpolation for interior frequencies, extrapolation for edge frequencies, with an affine variant fitted on the development split only. All baseline rules were fixed before results were inspected, and the reported contrast uses whichever baseline achieved the best held-out coefficient of determination, a conservative choice that can only shrink any Atlas advantage.

Results above the canonical range. At 8 kHz the Atlas outperformed the baseline in both large cohorts. In the occupational archive, across 2,046,160 held-out examinations, five coordinates improved the coefficient of determination by 0.0198 (95% CI 0.0194 to 0.0202) and reduced root-mean-square error by 0.312 dB. In US NHANES 2011-2012, across 3,818 held-out ear-examinations, the improvement was 0.0153 (0.0089 to 0.0222) with a root-mean-square reduction of 0.291 dB. Adding the sixth coordinate changed neither result materially.

Results at an interior frequency. At 1.5 kHz, evaluated in the Münster series[11] on 889 held-out observations, Atlas prediction and log-frequency interpolation were statistically indistinguishable: the difference in coefficient of determination was 0.0015 (-0.0013 to 0.0042) with five coordinates. Both approaches explained about 94% of the variance. Where a frequency lies between two measured neighbours, simple interpolation is already close to optimal and the Atlas adds nothing.

Results below the canonical range. Below 0.5 kHz the Atlas performed worse than the baseline. At 250 Hz the five coordinates lost 0.0238 of explained variance (-0.0410 to -0.0080) and at 125 Hz they lost 0.0301 (-0.0488 to -0.0118), in both cases increasing root-mean-square error by about 0.5 dB. Adding the sixth coordinate reversed the sign of both contrasts, to +0.0084 and +0.0125 respectively, but the gains are small. In the same cohort the Atlas also lost at 8 kHz, by 0.0115, in contrast to the two large cohorts; the Münster series comprises 356 unilateral ear-series and its extra-frequency contrasts are correspondingly imprecise.

Conclusion. The canonical coordinates extrapolate upward but not downward. This is consistent with the construction of the frame, whose highest-frequency loadings are well determined by 4 and 6 kHz while nothing in the canonical grid constrains the low-frequency edge. Thresholds below 0.5 kHz should be measured rather than inferred from Atlas coordinates.

#### S22. Severity dependence of Atlas-coordinate diversity, and its divergence from the correlation-dimension result

S4 and S19 locate a maximum of correlation dimension in moderate hearing loss, and S15 locates a maximum of the longitudinal participation ratio in the 40-49 dB band. We separately computed the severity dependence of the cross-sectional Atlas-coordinate participation ratio, using overall hearing level as the stratifier and taking the per-bin covariance over the identifiable shape coordinates only, so that the stratifying coordinate never enters the quantity being stratified. Cohorts were pooled by severity bin only within an identical grid signature, and a bin below the occupancy floor was left not evaluable rather than rescued by pooling with a neighbouring bin, another grid or another cohort.

Seven arms were prespecified: all eligible cohorts, population surveys only, occupational archive excluded, occupational archive only, and three leave-one-cohort-family-out arms removing the Korean, US and clinical families in turn. Eleven of thirteen arm-grid strata were evaluable. The occupational-only arm selected no eligible cohort under its membership rule and is reported as a prespecified empty arm rather than as a failure.

In every evaluable arm and on both grid signatures, this estimator placed its maximum in the lowest severity bin, not in moderate loss. On the six-frequency grid the peak participation ratio was 4.109 at a median overall hearing level of 5.83 dB in the all-eligible arm, and the leave-one-family-out arms gave 4.142, 4.015 and 4.109 with the same peak location. The bootstrap interval on the peak bin index did not extend beyond the lowest bin on the six-frequency grid in any arm.

This is a genuine divergence between estimators and we report it as such. Correlation dimension is computed on raw threshold vectors in the native frequency space of each cohort and measures how the number of similar audiograms grows with permitted distance; the Atlas-coordinate participation ratio is computed after projection onto a fixed five-dimensional frame and measures how evenly variance is distributed across that frame's shape axes. They are different quantities and were never pooled or treated as interchangeable. The intermediate-severity maximum reported in S4, S19 and S15 is reproducible within the estimators that produce it, including across leave-one-family-out resampling and across independent native grids, but it is not a property that every measure of audiogram diversity shares. We therefore do not present it as an estimator-independent biological constant, and the main text states this limitation explicitly.

#### S23. Glossary of Atlas terms

To ensure consistency and interoperability across clinical practice, audiological research and therapeutic trials, this study establishes a standardized glossary for the HHA.

The HHA (The Canonical Map): The standardized reference map of human hearing. A continuous, 5-dimensional geometric representation of clinical pure-tone thresholds.

Universal reference system: The clinical framework introduced by the Atlas. It supplements conventional severity scales by allowing uniform, quantitative measurement and comparison of the complete audiogram across global health systems.

HHA coordinate space (or human hearing space): The continuous multidimensional geometric space in which the audiograms are located. Every possible human audiogram occupies a single, mathematically defined location within this space.

Universal coordinates (HA coordinates): The set of five numerical values, HAC1 through HAC5, assigned to a patient's ear, representing their exact location within the human hearing space. These coordinates reconstruct the clinical audiogram to an accuracy of 1.35 dB.

Hearing state: the state of hearing function, defined by the HAC coordinates in the Human Hearing Atlas.

5-Dimensions: The five independent, continuous axes of human hearing measured by the universal coordinates: HAC1, overall impairment magnitude; HAC2, high-frequency decline (slope); HAC3, broad overarching curvature; HAC4, localized contrast at 2000 Hz; and HAC5, localized contrast at 3000 Hz. A sixth coordinate, HAC6, a localized contrast at 1000 Hz, completes the six-frequency representation.

Interoperability: The ability of the universal reference system to transfer and translate patient audiograms across different historical eras, national populations, calibration standards and measurement routes, including air and bone conduction, without requiring researchers to derive new, cohort-specific categorizations.

The Categorical Framework (The Conventional Paradigm): The conventional approach in audiology and otological research that utilizes discrete groupings and summary metrics. While foundational to clinical practice over the past century, large-scale predictive analyses indicate these methods omit geometric detail and are highly sensitive to analytical specifications.

Subtype taxonomy: The conventional approach in otological research that classifies human audiograms into a fixed catalog of distinct disease groups. The present predictive analyses suggest that a universal taxonomy is not supported by out-of-sample testing across populations.

Audiogram subtypes: Categorical groups, previously referred to in the literature as phenotypes, classes or clusters. The current findings indicate these behave as outputs of the sample and analytical specification rather than stable, reproducible biological entities across different cohorts.

Discrete model: A statistical formulation that partitions continuous data into mutually exclusive groups.

Continuous model: A statistical and conceptual representation that allows patient data to vary naturally along a multidimensional spectrum, accommodating individual variation without requiring assignment to isolated categories.

Pure-tone average (PTA): A widely used clinical summary metric that measures the absolute magnitude of impairment, HAC1. While highly practical, it inherently omits independently varying dimensions such as slope, curvature and localized frequency contrasts.

WHO severity scale: The conventional six-level clinical grading system, mild to profound, based on the PTA. The Atlas demonstrates that patients within the same severity grade can possess distinct audiometric configurations relevant to therapeutic management.

ASHA configuration: Conventional clinical descriptors for audiogram shapes, such as flat, sloping, notched and cookie-bite. The Atlas maps these terms to contiguous, heavily overlapping regions of a continuous spectrum, rather than mutually exclusive biological states.

Methodological artifacts: Analytical phenomena and data characteristics that contribute to the statistical recovery of distinct categorical groups, influencing how continuous variation is algorithmically interpreted.

Finite mixture model specification: The use of symmetric Gaussian mixture models to represent skewed clinical distributions. A clustering algorithm will inherently stack multiple symmetric components to absorb natural skewness, which can inflate the estimated subtype count.

Dependent observations (data leakage): Treating a single patient's left and right ears as independent observations during model training and evaluation. This practice inflates apparent structure and can lead to over-optimistic model performance, as the model learns one ear and is scored on its highly correlated partner.

Sample-dependent subtype counts: The statistical phenomenon, established in mixture model theory, where the recovered number of groups changes as a function of sample size, random sampling or measured frequencies, rather than reflecting a fixed biological property.

5 dB quantisation artifact: A structural effect arising from standard clinical audiometers recording thresholds in 5 dB rounding steps. This rounding creates heavy concentrations of mathematically identical audiograms that clustering algorithms may interpret as dense groups rather than measurement characteristics.

Cohort pooling: An analytical effect observed when pooling datasets collected across different generations, decades or eras. This aggregation smooths continuous variation and can erase distinct local cohort structures, altering the recovered classifications.

#### S24. Independent recovery of the Atlas architecture from unlabelled cohorts

Question. The canonical coordinates were derived once and frozen. A separate question is whether cohorts that contributed nothing to that derivation recover the same directions from their own data. This section reports that test. It concerns the orientation of the axes, not reconstruction accuracy, which is addressed in S5 and S8.

Cohorts. Four cohorts entered the core panel: two US national surveys of the 1960s recorded under ASA-1951 calibration, a German clinical cohort[9] measured on a low-frequency grid, and an occupational archive[8] of 2,731,474 ear-examinations from 1,365,737 workers. The remaining cohorts, all analysed identically, were a Turkish clinical cohort[17] of 648 ears from 324 patients; an Iranian clinical cohort[20] of 962 ears from 495 participants; five held-out Korean survey waves totalling 18,908 persons, whose 2009 to 2012 derivation waves were never touched and whose grid differs from them; the Münster series[11] of 356 independent ear-series; and three four-frequency historical US surveys of 6,713, 5,692 and 6,931 persons, none of which is in any derivation pool.

Blinding. Each cohort was decomposed without access to the Atlas frame, using only that cohort's own thresholds on its native grid. All comparisons used the coordinate system derived exclusively from the 89,752 ISO/ANSI-referenced ears.

Matching and criterion. Each cohort axis was matched to an Atlas axis objectively and one to one. Congruence is the absolute Tucker coefficient. Recovery required the 2.5th percentile of a 2,000-replicate cluster bootstrap to exceed the 99.9th percentile of a random-orientation null of 200,000 draws evaluated identically and computed per comparison rather than shared. Point congruence alone never determined recovery. Split-half stability over 200 cluster-level splits had to reach 0.90 for a comparison to be admissible.

Centrings. Two centrings were declared in advance. The primary arm is column-centred. The secondary arm is de-levelled, because the Atlas shape axes are constructed orthogonal to overall level and a like-for-like comparison of shape directions requires the level dimension to be removed on both sides. The secondary arm may explain the primary result and may never replace it. The two are reported separately in Supplementary Tables S4b and S4c and are never merged.

Identifiability. Axes that a cohort's native grid cannot carry were excluded in advance rather than estimated. The German cohort measures neither 3 nor 6 kHz and therefore admits only three axes; the Iranian grid likewise admits three. Fewer identifiable axes raise the random-basis null, so these cohorts face a stricter threshold than cohorts on the canonical grid. S9 sets out which coordinates each grid can carry.

Prespecified verdict. The five-part conjunctive rule was not satisfied. It failed two independent conditions: no core family returned full recovery in the primary arm, and the German cohort returned no recovery. Either alone is sufficient for the rule to fail. This verdict is a conclusion based on the joint satisfaction of all prespecified criteria and is reported separately from the axis-level results below; the two are not interchangeable.

Level. In all six cohorts the leading native axis was overall hearing level, carrying 55.5% and 56.0% in the 1960s surveys, 78.3% in the German cohort, 72.0% in the occupational archive, 78.4% in the Turkish cohort and 82.2% in the Iranian cohort, at split-half stability of 0.998 or above throughout. No Atlas information was available to any of these decompositions.

Shape, primary arm. Axes clearing the null were HAC3, HAC4 and HAC5 in the 1963-1965 survey; HAC4 and HAC5 in the 1966-1970 survey; HAC2, HAC3 and HAC6 in the occupational archive; HAC2 in the Turkish cohort, at a congruence of 0.983 with a bootstrap lower bound of 0.954 against a null of 0.902, the strongest single correspondence in the panel; HAC4 in the Iranian cohort; and none in the German cohort. The Turkish and Iranian verdicts are partial recovery, one of three identifiable axes each.

Shape, de-levelled arm. Axes clearing the null were HAC3, HAC4 and HAC5 in the 1963-1965 survey; HAC2, HAC4, HAC5 and HAC6 in the 1966-1970 survey; all five of HAC2 to HAC6 in the occupational archive; HAC2 in the Iranian cohort; and none in the Turkish or German cohorts. All five held-out Korean waves recovered HAC2, HAC4 and HAC6 in this arm, with HAC2 at 0.994 to 0.997, lower bounds 0.988 to 0.995, and every split-half at or above 0.995; the pooled Korean cohort and its five waves are one cohort family, not six independent replications.

Each of HAC2 to HAC6 was recovered by at least two distinct cohort families spanning 1963 to 2010, two calibration standards and five countries.

German cohort. Its local 2 kHz equivalent reached a point congruence of 0.848 with split-half stability of 0.992 in the de-levelled arm, but a bootstrap lower bound of 0.814 against a null of 0.868. Because the verdict reads the lower bound, this is a failure to clear the threshold rather than a marginal pass. The cohort's three-axis identifiability and correspondingly raised null are the operative constraint.

Resolution control. The same workers, examinations and ears from the occupational archive were projected onto a four-frequency grid and decomposed again. Native axes present on the seven-tone grid have no four-tone counterpart and disappear. Every Atlas comparison in this control carries the verdict not evaluable on rank grounds, because four observed dimensions cannot identify five axes. The control contributes to no recovery verdict and is descriptive only.

Sample size does not explain the German result. One hundred replicates were drawn at 569 persons, the German cohort's person count, from the held-out Korean waves, preserving both ears and the clustering, under the same prespecified analysis with Wilson intervals. In the de-levelled arm all three target axes were recovered in 100 of 100 replicates, Wilson interval 0.963 to 1.000, with median lower bounds of 0.986, 0.933 and 0.926 against a null of 0.885. In the primary arm HAC4 and HAC6 recovered in 91 and 92 of 100 replicates and HAC2 in none. At the same person count the German cohort's de-levelled HAC4 lower bound was 0.814 against a null of 0.868, while the Korean median was 0.933 and the fifth percentile 0.912. Frequency support differs between the two cohorts, so this establishes only that 569 persons is a sufficient sample size for this estimand.

Frequency sampling determines recovery with the cohort held fixed. In the Münster series the same 356 independent ear-series returned no recovery on a near-German grid and at three separate eight-tone rungs, and partial recovery once 3 and 6 kHz completed the canonical set. Its HAC2 rose from 0.812 with a lower bound of 0.749 against a null of 0.908, to 0.970 with a lower bound of 0.952 against a null of 0.813: the estimate rose and the threshold fell. The outcome is not monotone in tone count, since three eight-tone rungs fail while the six canonical tones recover. Within this ladder the information carried at 3 and 6 kHz cannot be separated from canonical completeness itself; what the ladder tracks is canonical completeness rather than the number of tones.

Four-frequency historical surveys. Four observed coordinates cannot identify five axes, so the five-axis verdict is not evaluable on rank grounds and was not attempted. In a four-coordinate signature analysis against the frozen within-archive four-tone reference, the first three coordinates reached 0.970, 0.937 and 0.967 in the 1971 to 1974 survey, 0.843, 0.737 and 0.885 in the 1976 to 1980 survey, and 0.940, 0.904 and 0.958 in the 1982 to 1984 survey, with every split-half at or above 0.994. The fourth coordinate is the orthogonal complement of the first three within four dimensions, so its congruence of 0.995 to 0.999 is largely determined by agreement of the three-dimensional subspace and is not an independently replicated axis; the level-orthogonal null for that comparison is 0.996. Pooled combinations are sensitivity analyses and are never counted as independent confirmations.

Interpretation and limits. Independent recovery is constrained by instrument resolution rather than by population identity. The test establishes that cohorts which saw no Atlas information rediscover the same shape directions; it does not establish that any coordinate corresponds to a mechanism, and it is not a test of reconstruction accuracy.

#### S25. Measurement-route invariance: air and bone conduction in one Atlas

Every other analysis in this study concerns hearing measured through the ear canal. A coordinate system recovered only from air-conduction audiograms could in principle describe the behaviour of that measurement rather than the hearing state of the patient. Bone conduction bypasses the outer and middle ear and delivers the stimulus through the skull, so agreement between the two routes in one ear is a test of what the Atlas represents, not of how well it fits one instrument.

Cohort. Paired air- and bone-conduction thresholds came from the first US National Health and Nutrition Examination Survey, 13,826 ear-records from 6,913 participants, both routes measured in the same ears at one visit at 0.5, 1, 2 and 4 kHz. The source carries parallel bone-conduction series whose relationship is undocumented. They were frozen as separate series, never averaged, ranked, imputed between or pooled; every analysis ran independently in each, and a finding was required to hold in all of them. Complete four-tone paired ears numbered 6,844, 5,535 and 4,958 in the three series, from 3,522, 2,972 and 2,736 participants. Because the grid carries no 3 or 6 kHz thresholds, the coordinates requiring them were treated as unavailable throughout and were never synthesised, as specified in S9.

The frozen Atlas. The canonical air-conduction Atlas was applied unchanged to bone conduction, without refitting, rotation, rescaling or realignment. Bone-conduction coordinates were obtained directly from the fixed Atlas transformation.

Same-ear correspondence. For each ear, the bone-conduction coordinates were compared with the air-conduction coordinates of the same ear against a candidate set drawn from other participants matched on side, age within five years and hearing level, with shape matching prohibited. The endpoint is the percentile rank of the true cross-route partner among candidates, for which chance is 0.50. Mean percentile was 0.2496, 0.2853 and 0.3057 in the three series.

Severity removal. Two independent controls established that the correspondence does not rest on hearing loss severity. In the first, the level term was structurally deleted, so that no residual difference in overall hearing level could carry a correspondence, and the same-ear relationship was re-estimated on the level-free coordinates alone. In the second, the analysis was restricted to ears whose measured air-conduction thresholds were all 20 dB HL or lower. That restriction is defined at the level of the ear within a bone-conduction series, from the four measured air-conduction frequencies; it is not bilateral six-frequency normal hearing, the counts are ears rather than participants, and they are not additive across series. The correspondence held under both controls.

One representation against two. A two-representation model fitting a separate air-conduction basis, a separate bone-conduction basis and the mapping between them was compared against a one-representation model in which the frozen Atlas contributes no fitted basis parameters and only a route operator is fitted. The two models were scored on identical folds, identical test ears and identical regularisation, and the two-representation model never received fewer fitted parameters than its comparator, so it cannot lose for want of flexibility. The endpoint is the difference in held-out mean absolute error, shared minus separate, against a noninferiority margin of 0.5 dB fixed before results. Differences were 0.0191 dB (95% CI 0.0137 to 0.0242), 0.0066 dB (0.0036 to 0.0095) and 0.0025 dB (0.0001 to 0.0050) in the three series, all far inside the margin. A bone-specific representation adds nothing.

Saturation. The one-representation comparison was repeated across subsample sizes to establish that the result is not an artefact of the available sample. The estimated difference remained inside the margin at every subsample size in every series, with upper confidence bounds of 0.0464, 0.0115 and 0.0133 dB, and reached its full-sample value at 100 participants.

Bidirectional reconstruction. Reconstruction was evaluated in held-out participants in both directions under participant-clustered cross-validation. Reconstructing air conduction from bone conduction, the shared Atlas differed from route-specific representations by 0.0651, 0.0213 and 0.0158 dB, again inside the 0.5 dB margin. In the same analysis the frozen Atlas slightly outperformed a representation fitted within the cohort itself, by 0.0079, 0.0214 and 0.0043 dB. Bone conduction therefore carries enough patient-specific information to reconstruct the air-conduction audiogram, and not merely to be predicted from it. Inference is confined to the level-plus-shape rank at which the comparison is informative; full-rank agreement is algebraically forced by four thresholds and is quarantined as a control.

The scalar gap as comparator. The conventional air-bone gap entered only as the reduced comparator. Held-out reconstruction of air conduction from the full cross-route displacement was compared with reconstruction from the scalar gap on the same folds. The full relationship was better in every series. The air-bone gap captures only part of the air-bone relationship and does not preserve all clinically relevant information contained in the paired hearing state. This is a statement about what the scalar summary discards, and no claim is made here that the Atlas recovers the discarded information, nor any claim of clinical misclassification, which would require an external adjudicator.

Resampling and intervals. The unit of analysis is the ear, and every split, bootstrap resample and permutation carries both ears of a participant together. Cross-validation is five-fold by participant. Intervals are 95 per cent participant-clustered bootstrap percentile intervals from 2,000 resamples; permutation p values are reported at their replicate floor.

Prespecified verdict. The hypothesis, null, acceptance criterion and margin for each endpoint were fixed before execution. The verdict across the three series is that the same patient-specific hearing pattern survives the change of measurement route, that it survives the removal of hearing level, that one Atlas represents both routes, and that the relationship between the routes is bidirectional.

What this does not establish. The paired cohort is a single national survey measured on a four-frequency grid. These analyses establish invariance of the Atlas coordinates observable on that grid across air and bone conduction. They do not extend the Atlas to modalities outside pure-tone audiometry, and they make no claim regarding the anatomical or physiological origin of any particular hearing state.

**[20]** kavehmcsd. Audiogram Analysis for Tinnitus Detection [dataset]. Kaggle. https://www.kaggle.com/datasets/kavehmcsd/audiogram-analysis-for-tinnitus-detection
