## Supplementary Tables for "The Human Hearing Atlas: a canonical map of human hearing"

### Supplementary Table S1. Out-of-sample comparison of discrete and continuous audiogram models across populations and strata

*Each analysis compares a model assigning audiograms to a fixed number of mutually exclusive groups with a model allowing continuous variation, scored on a locked holdout of participants that took no part in fitting, model selection or component-count selection. Both ears of a participant are always assigned to the same side of every split. A negative margin favours the continuous model.*

| Analysis | Holdout participants | Holdout ears | Margin, nats per ear | 95% CI | Verdict | Selected components |
|---|---|---|---|---|---|---|
| KNHANES 2019-2023 | 2,975 | 5,950 | -0.7114 | -0.7394 to -0.6835 | Continuum | 11 |
| KNHANES 2009-2012 | 3,332 | 6,664 | -0.6374 | -0.7653 to -0.4052 | Continuum | 8 |
| US NHANES 1999-2020 | 3,532 | 7,064 | -0.7348 | -0.7657 to -0.7017 | Continuum | 10 |
| US NHANES 1971-1974 | 1,007 | 2,014 | -0.3956 | -0.4532 to -0.3355 | Continuum | 8 |
| Four-country ISO/ANSI census | 12,752 | 25,504 | -1.1117 | -1.1519 to -1.0460 | Continuum | 11 |
| Pooled adult | 7,250 | 14,500 | -1.1842 | -1.2038 to -1.1646 | Continuum | 15 |
| Pooled geriatric | 3,421 | 6,842 | -0.3537 | -0.4844 to -0.1196 | Continuum | 12 |
| Pooled youth | 2,079 | 4,158 | -1.1887 | -1.2266 to -1.1515 | Continuum | 2 |

Panel a. Margin is the discrete minus continuous predictive score per held-out ear. Intervals are 95% percentile intervals from 10,000 participant-grouped bootstrap resamples of the locked holdout.

*Panel b. Distribution of verdicts across all 73 analyses.*

| Stratum | Analyses | Continuum | Discrete | Indeterminate |
|---|---|---|---|---|
| Youth | 8 | 7 | 1 | 0 |
| Adult | 16 | 3 | 8 | 5 |
| Senior | 18 | 0 | 12 | 6 |
| Pooled | 7 | 7 | 0 | 0 |
| Wave | 24 | 8 | 8 | 8 |
| All analyses | 73 | 25 | 29 | 19 |

A verdict is continuum when the margin and its entire interval lie below zero, discrete when both lie above zero, and indeterminate when the interval contains zero.

*Panel c. Selected component counts among analyses receiving a discrete verdict.*

| Components | Analyses | Youth | Working age | Senior | Whole wave |
|---|---|---|---|---|---|
| 2 | 1 | 1 | 0 | 0 | 0 |
| 3 | 4 | 0 | 0 | 4 | 0 |
| 4 | 4 | 0 | 0 | 4 | 0 |
| 5 | 5 | 0 | 3 | 2 | 0 |
| 6 | 11 | 0 | 4 | 2 | 5 |
| 7 | 1 | 0 | 0 | 0 | 1 |
| 8 | 3 | 0 | 1 | 0 | 2 |

Recovered component counts ranged from 2 to 8 and were not stable across cohorts or survey waves.

*Panel d. Exact duplicate audiogram patterns under the 5 dB recording lattice.*

| Dataset family | Ear-curves | Distinct patterns | Largest cluster | In a duplicate cluster, % |
|---|---|---|---|---|
| Münster glycerol series | 1,778 | 1,648 | 4 | 13.8 |
| KNHANES | 43,568 | 31,274 | 30 | 41.0 |
| NIOSH occupational archive | 4,092,824 | 353,408 | 25,083 | 96.2 |
| US NHANES 1999-2020 | 46,184 | 28,535 | 51 | 51.7 |
| US historical surveys (pre-1999) | 27,712 | 12,656 | 66 | 69.5 |

Counted over the complete six-frequency threshold pattern. Families whose grid cannot carry the canonical six frequencies are excluded.

*Panel e. Reproducibility of the selected component count across participant-grouped outer folds among the 29 analyses receiving a discrete locked-holdout verdict.*

| Analysis | Stratum | Reported k | Evaluable folds | Observed k range | Distinct k values | Folds selecting reported k, n | Folds selecting reported k, % |
|---|---|---|---|---|---|---|---|
| KNHANES 2009 | Adult | 6 | 100 | 4 to 8 | 5 | 44 | 44.0% |
| KNHANES 2011 | Adult | 8 | 100 | 3 to 11 | 9 | 37 | 37.0% |
| KNHANES 2012 | Adult | 6 | 100 | 4 to 9 | 6 | 30 | 30.0% |
| KNHANES 2020 | Adult | 6 | 100 | 3 to 9 | 6 | 42 | 42.0% |
| KNHANES 2022 | Adult | 6 | 100 | 3 to 9 | 7 | 39 | 39.0% |
| NHANES 1999-2000 | Adult | 5 | 100 | 2 to 7 | 5 | 43 | 43.0% |
| NHANES 2001-2002 | Adult | 5 | 100 | 4 to 7 | 4 | 45 | 45.0% |
| NHANES 2003-2004 | Adult | 5 | 100 | 4 to 8 | 5 | 43 | 43.0% |
| KNHANES 2009 | Senior | 3 | 100 | 2 to 4 | 3 | 61 | 61.0% |
| KNHANES 2010 | Senior | 4 | 100 | 2 to 8 | 7 | 39 | 39.0% |
| KNHANES 2012 | Senior | 5 | 100 | 3 to 8 | 6 | 44 | 44.0% |
| KNHANES 2019 | Senior | 4 | 100 | 3 to 6 | 4 | 51 | 51.0% |
| KNHANES 2020 | Senior | 6 | 100 | 2 to 8 | 7 | 43 | 43.0% |
| KNHANES 2021 | Senior | 5 | 100 | 3 to 7 | 5 | 48 | 48.0% |
| KNHANES 2023 | Senior | 6 | 100 | 2 to 10 | 9 | 35 | 35.0% |
| NHANES 1999-2000 | Senior | 3 | 100 | 2 to 4 | 3 | 72 | 72.0% |
| NHANES 2005-2006 | Senior | 3 | 100 | 3 to 8 | 5 | 58 | 58.0% |
| NHANES 2009-2010 | Senior | 4 | 100 | 3 to 5 | 3 | 72 | 72.0% |
| NHANES 2011-2012 | Senior | 3 | 100 | 3 to 5 | 3 | 47 | 47.0% |
| NHANES 2015-2016 | Senior | 4 | 100 | 3 to 8 | 5 | 58 | 58.0% |
| KNHANES 2012 | Whole wave | 7 | 100 | 6 to 11 | 6 | 40 | 40.0% |
| NHANES 1999-2000 | Whole wave | 6 | 100 | 2 to 8 | 6 | 43 | 43.0% |
| NHANES 2001-2002 | Whole wave | 6 | 100 | 3 to 10 | 7 | 34 | 34.0% |
| NHANES 2003-2004 | Whole wave | 6 | 100 | 3 to 8 | 5 | 39 | 39.0% |
| NHANES 2005-2006 | Whole wave | 6 | 100 | 3 to 8 | 6 | 43 | 43.0% |
| NHANES 2009-2010 | Whole wave | 6 | 100 | 2 to 9 | 8 | 33 | 33.0% |
| NHANES 2011-2012 | Whole wave | 8 | 100 | 3 to 12 | 10 | 28 | 28.0% |
| NHANES 2017-2020 | Whole wave | 8 | 100 | 3 to 11 | 9 | 23 | 23.0% |
| KNHANES 2023 | Youth | 2 | 84 | 2 to 3 | 2 | 81 | 96.4% |

Reported k is the modal interval-arm component count across evaluable outer folds. Observed k range and distinct k values summarize independently selected component counts across the same participant-grouped outer folds used for model comparison. Folds with no admissible discrete candidate are excluded rather than imputed.

*Panel f. All 73 analyses.*

| Analysis | Stratum | Holdout participants | Holdout ears | Locked-holdout margin, nats per ear | 95% CI | Verdict | Reported k | Outer-fold mean margin | Outer-fold SE | Evaluable outer folds | Structurally empty outer folds |
|---|---|---|---|---|---|---|---|---|---|---|---|
| BEAR 2017-2018 | Whole wave | 13 | 26 | -0.2598 | -0.6350 to 0.0860 | Indeterminate | 2 | -0.3578 | 0.2094 | 50 | 0 |
| BEAR 2017-2018, senior | Senior | 11 | 22 | 0.2465 | -0.6899 to 1.3373 | Indeterminate | 2 | 0.3719 | 0.3321 | 61 | 11 |

| Analysis | Stratum | Holdout participants | Holdout ears | Locked-holdout margin, nats per ear | 95% CI | Verdict | Reported k | Outer-fold mean margin | Outer-fold SE | Evaluable outer folds | Structurally empty outer folds |
|---|---|---|---|---|---|---|---|---|---|---|---|
| Four-country ISO/ANSI census | Pooled | 12,752 | 25,504 | -1.1117 | -1.1519 to -1.0460 | Continuum | 11 | -1.0893 | 0.0150 | 100 | 0 |
| Hispanic HANES | Whole wave | 1,040 | 2,080 | -1.0094 | -1.0659 to -0.9539 | Continuum | 5 | -1.0034 | 0.0117 | 100 | 0 |
| KNHANES 2009 | Whole wave | 567 | 1,134 | 0.6128 | -0.1138 to 1.9374 | Indeterminate | 6 | 0.2739 | 0.1571 | 100 | 0 |
| KNHANES 2009-2012 | Pooled | 3,332 | 6,664 | -0.6374 | -0.7653 to -0.4052 | Continuum | 8 | -0.6625 | 0.0369 | 100 | 0 |
| KNHANES 2009, adult | Adult | 365 | 730 | 0.0741 | 0.0253 to 0.1245 | Discrete | 6 | 0.2727 | 0.1671 | 100 | 0 |
| KNHANES 2009, senior | Senior | 139 | 278 | 0.1132 | 0.0152 to 0.2160 | Discrete | 3 | 0.0957 | 0.0250 | 100 | 0 |
| KNHANES 2009, youth | Youth | 64 | 128 | -0.2331 | -0.4607 to -0.0114 | Continuum | 2 | 0.0431 | 0.0592 | 100 | 0 |
| KNHANES 2010 | Whole wave | 948 | 1,896 | -0.1926 | -0.2909 to -0.0801 | Continuum | 10 | 0.1679 | 0.1838 | 100 | 0 |
| KNHANES 2010, adult | Adult | 619 | 1,238 | 0.2998 | -0.3393 to 1.5139 | Indeterminate | 4 | -0.1789 | 0.0270 | 100 | 0 |
| KNHANES 2010, senior | Senior | 238 | 476 | 0.3240 | 0.1871 to 0.4728 | Discrete | 4 | 0.3976 | 0.2711 | 100 | 0 |
| KNHANES 2010, youth | Youth | 91 | 182 | -0.5176 | -0.7129 to -0.3458 | Continuum | 2 | -0.5433 | 0.0458 | 100 | 0 |
| KNHANES 2011 | Whole wave | 914 | 1,828 | 0.2323 | -0.2138 to 1.0544 | Indeterminate | 6 | 0.0624 | 0.0905 | 100 | 0 |
| KNHANES 2011, adult | Adult | 577 | 1,154 | 0.1336 | 0.0807 to 0.1938 | Discrete | 8 | 0.1567 | 0.0126 | 100 | 0 |
| KNHANES 2011, senior | Senior | 254 | 508 | 0.0551 | -0.0184 to 0.1315 | Indeterminate | 3 | 0.0811 | 0.0171 | 100 | 0 |
| KNHANES 2011, youth | Youth | 83 | 166 | -0.3649 | -0.5420 to -0.2179 | Continuum | 2 | -0.4494 | 0.0431 | 100 | 0 |
| KNHANES 2012 | Whole wave | 839 | 1,678 | 0.1315 | 0.0922 to 0.1768 | Discrete | 7 | 0.2199 | 0.0742 | 100 | 0 |
| KNHANES 2012, adult | Adult | 513 | 1,026 | 0.1307 | 0.0854 to 0.1776 | Discrete | 6 | 0.1688 | 0.0180 | 100 | 0 |
| KNHANES 2012, senior | Senior | 255 | 510 | 0.2523 | 0.1160 to 0.4280 | Discrete | 5 | 0.0983 | 0.0205 | 100 | 0 |
| KNHANES 2012, youth | Youth | 72 | 144 | -5.4551 | -15.2105 to -0.5048 | Continuum | 2 | 0.4628 | 1.0211 | 100 | 0 |
| KNHANES 2019 | Whole wave | 274 | 548 | -0.1106 | -0.2229 to 0.0019 | Indeterminate | 7 | 0.0860 | 0.0190 | 100 | 0 |
| KNHANES 2019-2023 | Pooled | 2,975 | 5,950 | -0.7114 | -0.7394 to -0.6835 | Continuum | 11 | -0.6931 | 0.0220 | 100 | 0 |
| KNHANES 2019, adult | Adult | 150 | 300 | -0.2499 | -0.3613 to -0.1420 | Continuum | 4 | 0.6825 | 0.8035 | 100 | 0 |
| KNHANES 2019, senior | Senior | 125 | 250 | 0.0819 | 0.0095 to 0.1556 | Discrete | 4 | 0.1136 | 0.0214 | 100 | 0 |
| KNHANES 2020 | Whole wave | 552 | 1,104 | 0.6967 | -0.0276 to 2.0714 | Indeterminate | 8 | 0.0225 | 0.0157 | 100 | 0 |
| KNHANES 2020, adult | Adult | 277 | 554 | 0.1492 | 0.0519 to 0.2756 | Discrete | 6 | 0.1382 | 0.0173 | 100 | 0 |
| KNHANES 2020, senior | Senior | 275 | 550 | 0.1348 | 0.0861 to 0.1845 | Discrete | 6 | 0.1316 | 0.0175 | 100 | 0 |
| KNHANES 2021 | Whole wave | 535 | 1,070 | 0.7024 | -0.0175 to 2.0819 | Indeterminate | 8 | 0.1699 | 0.1195 | 100 | 0 |
| KNHANES 2021, adult | Adult | 258 | 516 | -0.0977 | -0.2131 to 0.0332 | Indeterminate | 6 | -0.0303 | 0.0270 | 100 | 0 |
| KNHANES 2021, senior | Senior | 276 | 552 | 0.1081 | 0.0680 to 0.1472 | Discrete | 5 | 0.1299 | 0.0267 | 100 | 0 |
| KNHANES 2022 | Whole wave | 747 | 1,494 | -0.4245 | -0.4991 to -0.3408 | Continuum | 3 | -0.3960 | 0.0154 | 100 | not recorded |
| KNHANES 2022, adult | Adult | 407 | 814 | 0.1334 | 0.0818 to 0.1879 | Discrete | 6 | 0.1439 | 0.0133 | 100 | 0 |
| KNHANES 2022, senior | Senior | 300 | 600 | -0.0389 | -0.0927 to 0.0119 | Indeterminate | 2 | -0.0117 | 0.0151 | 100 | 0 |
| KNHANES 2023 | Whole wave | 728 | 1,456 | -0.3263 | -0.3812 to -0.2707 | Continuum | 5 | -0.2605 | 0.0170 | 100 | 0 |
| KNHANES 2023, adult | Adult | 424 | 848 | -0.1603 | -0.2356 to -0.0867 | Continuum | 7 | -0.0769 | 0.0200 | 100 | 0 |
| KNHANES 2023, senior | Senior | 260 | 520 | 0.1101 | 0.0487 to 0.1724 | Discrete | 6 | 0.1164 | 0.0179 | 100 | 0 |
| KNHANES 2023, youth | Youth | 44 | 88 | 8.2255 | 0.0569 to 24.3573 | Discrete | 2 | 0.2309 | 0.0750 | 100 | 0 |
| NHANES 1999-2000 | Whole wave | 248 | 496 | 0.3290 | 0.1933 to 0.4900 | Discrete | 6 | 0.2198 | 0.0264 | 100 | 0 |
| NHANES 1999-2000, adult | Adult | 195 | 390 | 0.1295 | 0.0286 to 0.2267 | Discrete | 5 | 0.2356 | 0.0374 | 100 | 0 |
| NHANES 1999-2000, senior | Senior | 54 | 108 | 0.2129 | 0.0511 to 0.3814 | Discrete | 3 | 0.1180 | 0.0433 | 100 | 0 |
| NHANES 2001-2002 | Whole wave | 279 | 558 | 0.2157 | 0.1480 to 0.2891 | Discrete | 6 | 0.2066 | 0.0177 | 100 | 0 |
| NHANES 2001-2002, adult | Adult | 233 | 466 | 0.1157 | 0.0330 to 0.1993 | Discrete | 5 | 0.2228 | 0.0303 | 100 | 0 |
| NHANES 2001-2002, senior | Senior | 46 | 92 | 0.1976 | -0.0434 to 0.4389 | Indeterminate | 3 | 0.1475 | 0.0441 | 100 | 0 |
| NHANES 2003-2004 | Whole wave | 262 | 524 | 0.1859 | 0.0929 to 0.2868 | Discrete | 6 | 0.1781 | 0.0187 | 100 | 0 |
| NHANES 2003-2004, adult | Adult | 212 | 424 | 0.2552 | 0.1634 to 0.3548 | Discrete | 5 | 0.2602 | 0.0532 | 100 | 0 |

| Analysis | Stratum | Holdout participants | Holdout ears | Locked-holdout margin, nats per ear | 95% CI | Verdict | Reported k | Outer-fold mean margin | Outer-fold SE | Evaluable outer folds | Structurally empty outer folds |
|---|---|---|---|---|---|---|---|---|---|---|---|
| NHANES 2003-2004, senior | Senior | 51 | 102 | 0.1337 | -0.0320 to 0.3028 | Indeterminate | 3 | 0.1509 | 0.0505 | 100 | 0 |
| NHANES 2005-2006 | Whole wave | 399 | 798 | 0.0722 | 0.0187 to 0.1359 | Discrete | 6 | 0.1363 | 0.0102 | 100 | 0 |
| NHANES 2005-2006, senior | Senior | 99 | 198 | 0.1620 | 0.0393 to 0.2837 | Discrete | 3 | 0.1555 | 0.0312 | 100 | 0 |
| NHANES 2005-2006, youth | Youth | 225 | 450 | -0.6817 | -0.7867 to -0.5740 | Continuum | 2 | -0.3353 | 0.3037 | 100 | 0 |
| NHANES 2007-2008 | Whole wave | 170 | 340 | -0.7127 | -0.8455 to -0.5960 | Continuum | 3 | -0.6224 | 0.0501 | 100 | 0 |
| NHANES 2007-2008, adult | Adult | 40 | 80 | 0.1067 | -0.0192 to 0.2337 | Indeterminate | 2 | 0.3118 | 0.0638 | 100 | 0 |
| NHANES 2007-2008, youth | Youth | 130 | 260 | -0.4733 | -0.6833 to -0.1778 | Continuum | 2 | 0.2731 | 0.5097 | 100 | 0 |
| NHANES 2009-2010 | Whole wave | 317 | 634 | 0.1481 | 0.0970 to 0.1979 | Discrete | 6 | 0.1564 | 0.0197 | 100 | 0 |
| NHANES 2009-2010, senior | Senior | 131 | 262 | 0.1389 | 0.0178 to 0.2753 | Discrete | 4 | 0.1247 | 0.0247 | 100 | 0 |
| NHANES 2009-2010, youth | Youth | 144 | 288 | -0.6090 | -0.7026 to -0.5194 | Continuum | 2 | -0.2177 | 0.3974 | 100 | 0 |
| NHANES 2011-2012 | Whole wave | 573 | 1,146 | 0.1813 | 0.1334 to 0.2300 | Discrete | 8 | 0.1474 | 0.0115 | 100 | 0 |
| NHANES 2011-2012, adult | Adult | 466 | 932 | 0.0327 | -0.0625 to 0.1313 | Indeterminate | 8 | 0.0187 | 0.0306 | 100 | 0 |
| NHANES 2011-2012, senior | Senior | 107 | 214 | 0.1667 | 0.0411 to 0.2999 | Discrete | 3 | 0.1224 | 0.0287 | 100 | 0 |
| NHANES 2015-2016 | Whole wave | 639 | 1,278 | 0.0483 | -0.1131 to 0.2755 | Indeterminate | 5 | 0.0395 | 0.0192 | 100 | 0 |
| NHANES 2015-2016, adult | Adult | 513 | 1,026 | -0.1626 | -0.2755 to -0.0191 | Continuum | 7 | -0.1081 | 0.0214 | 100 | 0 |
| NHANES 2015-2016, senior | Senior | 127 | 254 | 0.2379 | 0.1094 to 0.3826 | Discrete | 4 | 0.1749 | 0.0327 | 100 | 0 |
| NHANES 2017-2020 | Whole wave | 576 | 1,152 | 0.0830 | 0.0449 to 0.1255 | Discrete | 8 | 0.1205 | 0.0103 | 100 | 0 |
| NHANES I | Whole wave | 1,007 | 2,014 | -0.3956 | -0.4532 to -0.3355 | Continuum | 8 | -0.4025 | 0.0122 | 100 | 0 |
| NHES Cycle II | Whole wave | 1,064 | 2,128 | -0.9474 | -1.0070 to -0.8851 | Continuum | 7 | -0.8226 | 0.0848 | 100 | 0 |
| NHES Cycle III | Whole wave | 1,015 | 2,030 | -0.5352 | -0.6005 to -0.4680 | Continuum | 8 | -0.5060 | 0.0652 | 100 | 0 |
| OHHR 2013-2015 | Whole wave | 85 | 170 | 0.1218 | -0.0579 to 0.3206 | Indeterminate | 3 | 0.1496 | 0.0465 | 100 | 0 |
| OHHR 2013-2015, adult | Adult | 12 | 24 | 0.2445 | -0.7065 to 1.2208 | Indeterminate | 2 | 0.1607 | 0.9893 | 68 | 12 |
| OHHR 2013-2015, senior | Senior | 73 | 146 | 0.0598 | -0.0852 to 0.2026 | Indeterminate | 3 | 0.1474 | 0.0405 | 100 | 0 |
| Pooled adult | Pooled | 7,250 | 14,500 | -1.1842 | -1.2038 to -1.1646 | Continuum | 15 | -1.1703 | 0.0207 | 100 | 0 |
| Pooled geriatric | Pooled | 3,421 | 6,842 | -0.3537 | -0.4844 to -0.1196 | Continuum | 12 | -0.4182 | 0.0273 | 100 | 0 |
| Pooled youth | Pooled | 2,079 | 4,158 | -1.1887 | -1.2266 to -1.1515 | Continuum | 2 | -1.2110 | 0.0076 | 100 | 0 |
| US NHANES 1999-2020 | Pooled | 3,532 | 7,064 | -0.7348 | -0.7657 to -0.7017 | Continuum | 10 | -0.6681 | 0.0319 | 100 | 0 |

The locked-holdout margin and its interval determine the verdict. The outer-fold mean and standard error summarise model-selection stability across participant-grouped outer folds and are descriptive only; they are never used inferentially and no paired or binomial fold test is performed.

Holdout participants and holdout ears are the locked-holdout denominators, not total cohort size. Structurally empty outer folds contain no held-out ears, leave the per-ear margin undefined, and are neither ties nor failures. For one analysis, the number of structurally empty folds was not recorded in the original result schema; its 100 evaluable folds are reported without imputation.

An em dash marks a quantity that is not scientifically defined for that analysis; no component count is shown where none was selected.

*Panel g. Sensitivity of discreteness classification to the 5 dB measurement model.*

| Point-arm verdict | Interval: Continuum | Interval: Discrete | Interval: Indeterminate |
|---|---|---|---|
| Continuum | 24 | 18 | 0 |
| Discrete | 0 | 31 | 0 |
| Indeterminate | 0 | 0 | 0 |

The point arm treats recorded thresholds as exact values, whereas the interval-aware arm treats each recorded threshold as representing its 5 dB measurement interval. Counts are paired verdicts across all 73 analyses under otherwise identical cohort, stratum, model-family and validation designs. Interval-aware treatment shifted 18 analyses from continuum to discrete and none from discrete to continuum. This sensitivity analysis quantifies dependence of the statistical verdict on the measurement model and is distinct from Panel d, which quantifies concentration of observed audiograms on the 5 dB recording lattice.

#### Supplementary Table S2. Reconstruction of held-out audiograms as a function of the number of Human Hearing Atlas coordinates

*Reconstruction accuracy in 22,438 held-out ears from 11,219 participants who contributed nothing to the derivation of the coordinates.*

| Coordinates | Number | MAE, dB | RMSE, dB | 95th percentile, dB | Thresholds within 5 dB, % | Whole audiograms within 5 dB, % | Pairs preserved, % |
|---|---|---|---|---|---|---|---|
| HAC1 | 1 | 7.6417 | 10.7905 | 23.333 | 50.712 | 7.871 | not applicable |
| HAC1-HAC2 | 2 | 4.9578 | 6.5584 | 13.280 | 60.898 | 15.514 | not applicable |
| HAC1-HAC3 | 3 | 3.1985 | 4.4348 | 9.118 | 78.678 | 38.671 | not applicable |
| HAC1-HAC4 | 4 | 2.2491 | 3.2459 | 6.793 | 88.483 | 61.235 | not applicable |
| HAC1-HAC5 | 5 | 1.3495 | 2.1592 | 4.743 | 95.662 | 81.358 | 99.929 |
| HAC1-HAC6 | 6 | 0.0000 | 0.0000 | 0.000 | 100.000 | 100.000 | not applicable |

Whole-audiogram accuracy requires all six measured frequencies of an ear to fall within 5 dB. Pair preservation is the percentage of clinically distinguishable audiogram pairs that remain distinguishable after reconstruction, evaluated over 1,967,953 sampled pairs differing by more than 5 dB at one or more frequencies, of which 1,398 merged.

Accuracy across five independent participant-level splits ranged from 1.3434 to 1.3526 dB.

#### Supplementary Table S3. External validation of the frozen Human Hearing Atlas across populations, calibration standards and audiometric frequency grids

*Cohorts that contributed nothing to the derivation of the coordinates. The frozen coordinates were applied without any refitting, rescaling or per-coordinate adjustment.*

| Cohort | Country | Years | Grid and calibration standard | Audiograms or ear-examinations | Frozen MAE, dB | Within 5 dB, % | Local-fit penalty, dB |
|---|---|---|---|---|---|---|---|
| NIOSH occupational archive | USA | 1981-2010 | 7 tone, ISO/ANSI | 4,092,824 | 1.018 | 98.19 | 0.190 |
| US NHES Cycle II | USA | 1963-1965 | 6 tone canonical, ASA-1951 | 14,184 | 1.0869 | 97.71 | 0.127 |
| US NHES Cycle III | USA | 1966-1970 | 6 tone canonical, ASA-1951 | 13,528 | 1.1317 | 97.55 | 0.037 |
| Münster glycerol series (Lütkenhöner and Basel) | Germany | 2013 | 10 tone; 6 canonical analysed, ISO/ANSI | 1,778 serial (356 ear-series) | 1.4082 | 94.69 | 0.248 |
| Cadenza CAD1 | UK | 2023 | 6 tone canonical, ISO/ANSI | 106 | 1.5709 | 94.50 | Frozen Atlas only; no local comparator |
| Iranian clinical cohort | Iran | 2023 | 6 canonical frequencies, eligible subset, ISO/ANSI | 158 | 1.2269 | 96.20 | Frozen Atlas only; no local comparator |
| KNHANES 2019 | South Korea | 2019 | 4 tone reduced, ISO/ANSI | 3,660 | 1.8435 | 91.55 | -0.048 |
| KNHANES 2020 | South Korea | 2020 | 4 tone reduced, ISO/ANSI | 7,364 | 1.8552 | 91.19 | -0.058 |
| OHHR | Germany | 2013-2015 | 4 tone reduced, ISO/ANSI | 1,138 | 1.7627 | 91.83 | -0.116 |
| Meltem Hospital | Türkiye | 2024 | 4 tone reduced, ISO/ANSI | 648 | 1.5858 | 93.90 | -0.133 |
| BEAR | Denmark | 2017-2018 | 4 tone reduced, ISO/ANSI | 172 | 1.4476 | 93.75 | Frozen Atlas only; no local comparator |
| Macquarie | Australia | 2024 | 4 tone reduced, ISO/ANSI | 121 | 0.9755 | 99.17 | Frozen Atlas only; no local comparator |

"Frozen Atlas only; no local comparator" indicates application of the frozen Atlas without a locally fitted comparator. All external evaluations used the frozen Atlas without refitting.

Penalty is the paired per-ear difference between the frozen representation and one fitted locally within the receiving population. A negative penalty indicates the frozen representation reconstructed better than the local one. Cohorts too small to support a locally fitted comparator are reported with the frozen Atlas only, without a penalty.

Held-out transfer experiments within the NHANES and KNHANES derivation families are reported separately in the Supplementary Information and are not listed here as independent external cohorts.

Twelve rows are shown for eleven evaluations: the 2019 and 2020 KNHANES waves are one evaluation of one cohort on one reduced grid, reported wave by wave here. Eight rows therefore correspond to the seven evaluations that supported a locally fitted comparator, and four rows to the four evaluated with the frozen Atlas only.

For the occupational archive the two accuracy figures come from different evaluation subsets and are not differences of one another: the frozen mean absolute error of 1.018 dB and 98.19% within 5 dB are measured over the complete archive of 4,092,824 ear-examinations, whereas the 0.190 dB local-fit penalty is the paired frozen-minus-local difference on the matched transfer subset of 2,731,474 ear-examinations from 1,365,737 workers, on which the frozen representation reconstructed to 0.9963 dB and a locally fitted one to 0.8064 dB.

The Iranian cohort was evaluated on the 158 of 1,004 source ears that carried all six canonical frequencies. The 846 excluded ears were excluded solely for missing canonical frequencies and not on any quality criterion: 725 lacked 3 kHz and 609 lacked 6 kHz, the two frequencies the reduced clinical grid most often omits. Coordinates requiring unmeasured frequencies were treated as unavailable and were not imputed.

The glycerol series contributes 1,778 serial ear-observations from 356 ear-series in 347 patients, each ear measured before and 1, 2, 3 and 4 hours after glycerol administration; the observations are repeated measurements of the same ears and are not independent audiograms. Patient-level linkage is unavailable in the deposited data. Ten frequencies were recorded and the six canonical frequencies were analysed.

*Panel b. Which coordinates a four-frequency audiogram can recover, measured as proportional reduction in the variance of each coordinate.*

| Coordinate | United States | Korea | Status under the 0.80 criterion |
|---|---|---|---|
| Overall hearing level | 0.9688 | 0.9703 | Recoverable |
| High-frequency slope | 0.8035 | 0.7893 | Recoverable in the United States only |
| Broad curvature | 0.2346 | 0.2646 | Not recoverable |
| Local 2 kHz contrast | 0.9048 | 0.8985 | Recoverable |
| Local 3 kHz contrast | 0.1849 | 0.1750 | Not recoverable |

The 0.80 criterion was fixed before analysis. Missing frequencies produce selective loss of specific coordinates rather than uniformly poorer precision.

*Panel c. Bamford–Bench historical findings, as reported in the primary papers.*

| Study | N | Frequencies, kHz | PC1, % | PC2, % | PC1+PC2, % |
|---|---|---|---|---|---|
| Bamford et al. 1980 | 201 | 0.25, 0.5, 1, 2, 4, 8 | 60.07 | 25.65 | 85.72 |
| Bench 1983 | 391 | 0.25, 0.5, 1, 2, 4, 8 | 68 | 19 | 87 |

Third component: 8.21% (Bamford et al.) and 6% (Bench). Neither study treated it as meaningful structure. Bench reports percentages rounded to whole figures.

These are the historical principal components of each study. They are not identified with any Human Hearing Atlas coordinate; no loading comparison was performed.

*Panel d. Bamford–Bench frequency-grid replication. Held-out NHES II and III ears (1963–1970) restricted to the historical grid of 0.25, 0.5, 1, 2, 4 and 8 kHz, withholding 3 and 6 kHz relative to the canonical Atlas grid, with the Atlas frozen. N = 11,084 held-out ears.*

| Coordinate | Interpretation | Variance reduction | Status under the 0.80 criterion |
|---|---|---|---|
| HAC1 | Overall hearing level | 0.933334 | Recoverable |
| HAC2 | High-frequency slope | 0.666048 | Below criterion |
| HAC3 | Broad curvature | 0.295897 | Not recoverable |
| HAC4 | Local 2 kHz contrast | 0.893626 | Recoverable |
| HAC5 | Local 3 kHz contrast | 0.281889 | Not recoverable |

Reconstruction error at the withheld frequencies was 4.467911 dB at 3 kHz and 6.460046 dB at 6 kHz.

The 0.80 recoverability threshold was prespecified. The two withheld frequencies were removed jointly; their individual contributions were not separately identified.

*Panel e. Independent NIOSH frequency-grid replication. 3 and 6 kHz jointly withheld from 1,638,338 held-out ear-examinations, evaluated under the same frozen-frame criterion.*

| Coordinate | Interpretation | Variance reduction | Status under the 0.80 criterion |
|---|---|---|---|
| HAC1 | Overall hearing level | 0.960994 | Recoverable |
| HAC2 | High-frequency slope | 0.888106 | Recoverable |
| HAC3 | Broad curvature | 0.445002 | Not recoverable |
| HAC4 | Local 2 kHz contrast | 0.903101 | Recoverable |
| HAC5 | Local 3 kHz contrast | 0.090136 | Not recoverable |

The same two coordinates fell below criterion as in the NHES replication.

#### Supplementary Table S4. Independent recovery of the Human Hearing Atlas coordinates

*Panel a. Spontaneous emergence of the overall-level axis in six independent cohorts, each decomposed on its native grid without access to the Atlas basis, rank, axes or labels.*

| Cohort | Tones | Ears | Clusters | A1 variance share | A1 split-half | phi to level | 2.5% lower | 97.5% upper |
|---|---|---|---|---|---|---|---|---|
| NHES Cycle II | 8 | 14184 | 7092 | 0.555432 | 0.999559 | 0.985654 | 0.981398 | 0.989112 |
| NHES Cycle III | 8 | 13528 | 6764 | 0.560446 | 0.999325 | 0.961309 | 0.950534 | 0.970001 |
| OHHR | 8 | 1138 | 569 | 0.783387 | 0.999293 | 0.991552 | 0.986978 | 0.994505 |
| NIOSH | 7 | 2731474 | 1365737 | 0.720077 | 0.999999 | 0.895789 | 0.895307 | 0.896278 |
| Türkiye (Meltem) | 6 | 648 | 324 | 0.783598 | 0.998416 | 0.971845 | 0.956426 | 0.983239 |
| Iran (Urmia) | 6 | 962 | 495 | 0.821521 | 0.999244 | 0.916049 | 0.898944 | 0.930722 |

Congruence is between the cohort's own leading axis and the uniform direction built from its frequency grid alone. The level axis carries no rotation null and is not part of the shape-recovery test; it is reported for its congruence and split-half stability.

*Panel b. Primary column-centred arm, one result per independent cohort family. Every identifiable shape axis is shown, including those that did not clear their null.*

| Cohort | HAC axis | Cohort axis | Congruence | 2.5% lower | 97.5% upper | Split-half | Comparable | Null 99.9% | Exceeds null | Mode |
|---|---|---|---|---|---|---|---|---|---|---|
| NHES Cycle II | HAC2 | A4 | 0.865567 | 0.778738 | 0.898331 | 0.982211 | Yes | 0.824797 | Does not clear null | direct |
| NHES Cycle II | HAC3 | A3 | 0.937752 | 0.882970 | 0.973227 | 0.982156 | Yes | 0.824797 | Clears null | direct |
| NHES Cycle II | HAC4 | A5 | 0.919919 | 0.900178 | 0.933937 | 0.992487 | Yes | 0.824797 | Clears null | direct |
| NHES Cycle II | HAC5 | A6 | 0.928710 | 0.868968 | 0.951092 | 0.983470 | Yes | 0.824797 | Clears null | direct |
| NHES Cycle II | HAC6 | A7 | 0.822621 | 0.766834 | 0.861854 | 0.985291 | Yes | 0.824797 | Does not clear null | direct |
| NHES Cycle III | HAC2 | A2 | 0.749375 | 0.709767 | 0.787316 | 0.997279 | Yes | 0.824128 | Does not clear null | direct |
| NHES Cycle III | HAC3 | A4 | 0.811268 | 0.751675 | 0.878389 | 0.990718 | Yes | 0.824128 | Does not clear null | direct |
| NHES Cycle III | HAC4 | A5 | 0.914733 | 0.893681 | 0.929428 | 0.993423 | Yes | 0.824128 | Clears null | direct |
| NHES Cycle III | HAC5 | A6 | 0.941311 | 0.902911 | 0.967988 | 0.990703 | Yes | 0.824128 | Clears null | direct |
| NHES Cycle III | HAC6 | A8 | 0.830869 | 0.807764 | 0.852748 | 0.997494 | Yes | 0.824128 | Does not clear null | direct |
| OHHR | HAC2 | A3 | 0.708291 | 0.648116 | 0.757351 | 0.992804 | Yes | 0.875529 | Does not clear null | interpolated |
| OHHR | HAC4 | A4 | 0.828637 | 0.789543 | 0.858623 | 0.991090 | Yes | 0.875529 | Does not clear null | interpolated |
| OHHR | HAC6 | A5 | 0.714263 | 0.617971 | 0.783899 | 0.978934 | Yes | 0.875529 | Does not clear null | interpolated |
| NIOSH | HAC2 | A3 | 0.867876 | 0.866863 | 0.868870 | 0.999979 | Yes | 0.820250 | Clears null | direct |
| NIOSH | HAC3 | A4 | 0.888391 | 0.887484 | 0.889247 | 0.999984 | Yes | 0.820250 | Clears null | direct |
| NIOSH | HAC4 | A5 | 0.759880 | 0.757278 | 0.762407 | 0.999980 | Yes | 0.820250 | Does not clear null | direct |
| NIOSH | HAC5 | A6 | 0.746505 | 0.743860 | 0.749145 | 0.999988 | Yes | 0.820250 | Does not clear null | direct |
| NIOSH | HAC6 | A7 | 0.942327 | 0.941629 | 0.942995 | 0.999998 | Yes | 0.820250 | Clears null | direct |
| Türkiye (Meltem) | HAC2 | A2 | 0.983267 | 0.953576 | 0.996254 | 0.991606 | Yes | 0.902227 | Clears null | interpolated |
| Türkiye (Meltem) | HAC4 | A4 | 0.705536 | 0.610871 | 0.820379 | 0.967288 | Yes | 0.902227 | Does not clear null | interpolated |
| Türkiye (Meltem) | HAC6 | A5 | 0.747784 | 0.647857 | 0.801361 | 0.979506 | Yes | 0.902227 | Does not clear null | interpolated |
| Iran (Urmia) | HAC2 | A2 | 0.928059 | 0.864192 | 0.968671 | 0.989255 | Yes | 0.903700 | Does not clear null | interpolated |
| Iran (Urmia) | HAC4 | A4 | 0.948509 | 0.906822 | 0.965144 | 0.980073 | Yes | 0.903700 | Clears null | interpolated |
| Iran (Urmia) | HAC6 | A5 | 0.836609 | 0.789541 | 0.869050 | 0.989949 | Yes | 0.903700 | Does not clear null | interpolated |
| KNHANES pooled 2019-2023 | HAC2 | A2 | 0.876085 | 0.862522 | 0.887838 | 0.999644 | Yes | 0.902352 | Does not clear null | interpolated |
| KNHANES pooled 2019-2023 | HAC4 | A4 | 0.959755 | 0.955618 | 0.963217 | 0.999615 | Yes | 0.902352 | Clears null | interpolated |
| KNHANES pooled 2019-2023 | HAC6 | A5 | 0.943721 | 0.942845 | 0.944252 | 0.999826 | Yes | 0.902352 | Clears null | interpolated |

Recovery requires the 2.5th bootstrap percentile to exceed that comparison's own 99.9th-percentile null, built from a 200,000-draw random-basis null pushed through the identical pipeline. Point congruence alone never determines recovery.

Axes absent from a cohort's rows were not identifiable on its native grid and were excluded in advance: OHHR carries HAC2, HAC4 and HAC6 only.

KNHANES is represented by its pooled 2019-2023 result. The five individual waves are within-family sensitivity analyses and appear in Panel d-i.

The prespecified five-part conjunctive criterion was not supported. It failed two independent conditions: no core family returned full recovery in this primary arm, and the German cohort returned no recovery. Either alone was sufficient for failure. The global conjunctive verdict and the axis-level results address different estimands and are reported separately.

*Panel c. Prespecified secondary de-levelled arm, the same seven cohort families. Successes and failures are both shown.*

| Cohort | HAC axis | Cohort axis | Congruence | 2.5% lower | 97.5% upper | Split-half | Comparable | Null 99.9% | Exceeds null | Mode |
|---|---|---|---|---|---|---|---|---|---|---|
| NHES Cycle II | HAC2 | A3 | 0.830380 | 0.797845 | 0.867937 | 0.985427 | Yes | 0.805294 | Does not clear null | direct |
| NHES Cycle II | HAC3 | A2 | 0.932152 | 0.874416 | 0.971908 | 0.985847 | Yes | 0.805294 | Clears null | direct |
| NHES Cycle II | HAC4 | A4 | 0.923296 | 0.903078 | 0.937776 | 0.992069 | Yes | 0.805294 | Clears null | direct |
| NHES Cycle II | HAC5 | A5 | 0.916499 | 0.834890 | 0.948002 | 0.979785 | Yes | 0.805294 | Clears null | direct |
| NHES Cycle II | HAC6 | A6 | 0.792721 | 0.753944 | 0.836597 | 0.983174 | Yes | 0.805294 | Does not clear null | direct |
| NHES Cycle III | HAC2 | A1 | 0.901716 | 0.872729 | 0.926528 | 0.998330 | Yes | 0.804585 | Clears null | direct |
| NHES Cycle III | HAC3 | A2 | 0.790683 | 0.770651 | 0.844312 | 0.993008 | Yes | 0.804585 | Does not clear null | direct |
| NHES Cycle III | HAC4 | A4 | 0.907372 | 0.886411 | 0.921619 | 0.993726 | Yes | 0.804585 | Clears null | direct |
| NHES Cycle III | HAC5 | A5 | 0.937408 | 0.892139 | 0.967845 | 0.987837 | Yes | 0.804585 | Clears null | direct |
| NHES Cycle III | HAC6 | A7 | 0.831236 | 0.807114 | 0.852951 | 0.997472 | Yes | 0.804585 | Clears null | direct |
| OHHR | HAC2 | A2 | 0.802506 | 0.755548 | 0.837958 | 0.992602 | Yes | 0.867569 | Does not clear null | interpolated |
| OHHR | HAC4 | A3 | 0.847584 | 0.813976 | 0.874877 | 0.992153 | Yes | 0.867569 | Does not clear null | interpolated |
| OHHR | HAC6 | A4 | 0.744234 | 0.656236 | 0.811032 | 0.979793 | Yes | 0.867569 | Does not clear null | interpolated |
| NIOSH | HAC2 | A1 | 0.980110 | 0.979933 | 0.980280 | 0.999996 | Yes | 0.790937 | Clears null | direct |
| NIOSH | HAC3 | A3 | 0.893252 | 0.892089 | 0.894304 | 0.999987 | Yes | 0.790937 | Clears null | direct |
| NIOSH | HAC4 | A4 | 0.895037 | 0.893009 | 0.896978 | 0.999979 | Yes | 0.790937 | Clears null | direct |
| NIOSH | HAC5 | A5 | 0.845546 | 0.843388 | 0.847617 | 0.999988 | Yes | 0.790937 | Clears null | direct |
| NIOSH | HAC6 | A6 | 0.947818 | 0.947153 | 0.948467 | 0.999998 | Yes | 0.790937 | Clears null | direct |
| Türkiye (Meltem) | HAC2 | A1 | 0.894916 | 0.851691 | 0.929657 | 0.995773 | Yes | 0.893316 | Does not clear null | interpolated |
| Türkiye (Meltem) | HAC4 | A3 | 0.683838 | 0.599797 | 0.804719 | 0.970360 | Yes | 0.893316 | Does not clear null | interpolated |
| Türkiye (Meltem) | HAC6 | A4 | 0.744713 | 0.639076 | 0.798058 | 0.978169 | Yes | 0.893316 | Does not clear null | interpolated |
| Iran (Urmia) | HAC2 | A1 | 0.953557 | 0.934874 | 0.966264 | 0.997693 | Yes | 0.893977 | Clears null | interpolated |
| Iran (Urmia) | HAC4 | A3 | 0.879774 | 0.828567 | 0.907961 | 0.981468 | Yes | 0.893977 | Does not clear null | interpolated |
| Iran (Urmia) | HAC6 | A4 | 0.821473 | 0.774758 | 0.856404 | 0.992358 | Yes | 0.893977 | Does not clear null | interpolated |
| KNHANES pooled 2019-2023 | HAC2 | A1 | 0.995875 | 0.994731 | 0.996822 | 0.999920 | Yes | 0.884674 | Clears null | interpolated |
| KNHANES pooled 2019-2023 | HAC4 | A3 | 0.962468 | 0.959207 | 0.965362 | 0.999639 | Yes | 0.884674 | Clears null | interpolated |
| KNHANES pooled 2019-2023 | HAC6 | A4 | 0.945410 | 0.944194 | 0.946017 | 0.999833 | Yes | 0.884674 | Clears null | interpolated |

This arm was declared in advance as secondary and explanatory. It may explain the primary arm but never replaces it, and the two arms are never merged.

*Panel d-i. KNHANES wave-by-wave sensitivity, both arms.*

| Wave | Arm | HAC axis | Cohort axis | Congruence | 2.5% lower | Null 99.9% | Split-half | Exceeds null | Verdict |
|---|---|---|---|---|---|---|---|---|---|
| KNHANES 2019 | Primary | HAC2 | A2 | 0.882528 | 0.825751 | 0.902352 | 0.994889 | Does not clear null | Partial recovery |
| KNHANES 2019 | Primary | HAC4 | A4 | 0.950381 | 0.934122 | 0.902352 | 0.996131 | Clears null | Partial recovery |
| KNHANES 2019 | Primary | HAC6 | A5 | 0.943541 | 0.936444 | 0.902352 | 0.997887 | Clears null | Partial recovery |
| KNHANES 2019 | De-levelled | HAC2 | A1 | 0.994068 | 0.987778 | 0.884674 | 0.998754 | Clears null | Full recovery |
| KNHANES 2019 | De-levelled | HAC4 | A3 | 0.956945 | 0.943233 | 0.884674 | 0.996279 | Clears null | Full recovery |
| KNHANES 2019 | De-levelled | HAC6 | A4 | 0.944711 | 0.936297 | 0.884674 | 0.998078 | Clears null | Full recovery |
| KNHANES 2020 | Primary | HAC2 | A2 | 0.881123 | 0.848536 | 0.902352 | 0.997544 | Does not clear null | Partial recovery |
| KNHANES 2020 | Primary | HAC4 | A4 | 0.957987 | 0.947655 | 0.902352 | 0.997979 | Clears null | Partial recovery |
| KNHANES 2020 | Primary | HAC6 | A5 | 0.942382 | 0.939812 | 0.902352 | 0.999179 | Clears null | Partial recovery |

| Wave | Arm | HAC axis | Cohort axis | Congruence | 2.5% lower | Null 99.9% | Split-half | Exceeds null | Verdict |
|---|---|---|---|---|---|---|---|---|---|
| KNHANES 2020 | De-levelled | HAC2 | A1 | 0.996310 | 0.993139 | 0.884674 | 0.999453 | Clears null | Full recovery |
| KNHANES 2020 | De-levelled | HAC4 | A3 | 0.961805 | 0.953774 | 0.884674 | 0.998528 | Clears null | Full recovery |
| KNHANES 2020 | De-levelled | HAC6 | A4 | 0.945051 | 0.941814 | 0.884674 | 0.999318 | Clears null | Full recovery |
| KNHANES 2021 | Primary | HAC2 | A2 | 0.910643 | 0.887369 | 0.902352 | 0.998160 | Does not clear null | Partial recovery |
| KNHANES 2021 | Primary | HAC4 | A4 | 0.958570 | 0.948767 | 0.902352 | 0.998237 | Clears null | Partial recovery |
| KNHANES 2021 | Primary | HAC6 | A5 | 0.944356 | 0.941649 | 0.902352 | 0.999054 | Clears null | Partial recovery |
| KNHANES 2021 | De-levelled | HAC2 | A1 | 0.997142 | 0.994877 | 0.884674 | 0.999498 | Clears null | Full recovery |
| KNHANES 2021 | De-levelled | HAC4 | A3 | 0.962508 | 0.954008 | 0.884674 | 0.998593 | Clears null | Full recovery |
| KNHANES 2021 | De-levelled | HAC6 | A4 | 0.945853 | 0.943245 | 0.884674 | 0.999211 | Clears null | Full recovery |
| KNHANES 2022 | Primary | HAC2 | A2 | 0.890943 | 0.870228 | 0.902352 | 0.998833 | Does not clear null | Partial recovery |
| KNHANES 2022 | Primary | HAC4 | A4 | 0.967965 | 0.961099 | 0.902352 | 0.998343 | Clears null | Partial recovery |
| KNHANES 2022 | Primary | HAC6 | A5 | 0.943687 | 0.940833 | 0.902352 | 0.999103 | Clears null | Partial recovery |
| KNHANES 2022 | De-levelled | HAC2 | A1 | 0.996187 | 0.994187 | 0.884674 | 0.999716 | Clears null | Full recovery |
| KNHANES 2022 | De-levelled | HAC4 | A3 | 0.967700 | 0.961636 | 0.884674 | 0.998367 | Clears null | Full recovery |
| KNHANES 2022 | De-levelled | HAC6 | A4 | 0.945070 | 0.941680 | 0.884674 | 0.999090 | Clears null | Full recovery |
| KNHANES 2023 | Primary | HAC2 | A2 | 0.806944 | 0.796766 | 0.902352 | 0.997164 | Does not clear null | Partial recovery |
| KNHANES 2023 | Primary | HAC4 | A4 | 0.955291 | 0.944931 | 0.902352 | 0.998312 | Clears null | Partial recovery |
| KNHANES 2023 | Primary | HAC6 | A5 | 0.943667 | 0.941191 | 0.902352 | 0.999218 | Clears null | Partial recovery |
| KNHANES 2023 | De-levelled | HAC2 | A1 | 0.994246 | 0.991439 | 0.884674 | 0.999648 | Clears null | Full recovery |
| KNHANES 2023 | De-levelled | HAC4 | A3 | 0.959381 | 0.951559 | 0.884674 | 0.998587 | Clears null | Full recovery |
| KNHANES 2023 | De-levelled | HAC6 | A4 | 0.945358 | 0.942476 | 0.884674 | 0.999376 | Clears null | Full recovery |

The five waves are sensitivity analyses within one South Korean cohort family and are not counted as independent replications.

*Panel d-ii. Münster frequency-grid ladder, both arms.*

| Grid variant | Role | Grid (Hz) | Tones | Arm | HAC axis | Congruence | 2.5% lower | Null 99.9% | Split-half | Result |
|---|---|---|---|---|---|---|---|---|---|---|
| Münster, all timepoints | Perturbation sensitivity | 125, 250, 500, 1000, 1500, 2000, 3000, 4000, 6000, 8000 | 10 | Primary | HAC2 | 0.973025 | 0.949707 | 0.832607 | 0.992871 | Partial recovery |
| Münster, all timepoints | Perturbation sensitivity | 125, 250, 500, 1000, 1500, 2000, 3000, 4000, 6000, 8000 | 10 | Primary | HAC3 | 0.719196 | 0.657423 | 0.832607 | 0.885128 | Partial recovery |
| Münster, all timepoints | Perturbation sensitivity | 125, 250, 500, 1000, 1500, 2000, 3000, 4000, 6000, 8000 | 10 | Primary | HAC4 | 0.670904 | 0.566821 | 0.832607 | 0.916798 | Partial recovery |
| Münster, all timepoints | Perturbation sensitivity | 125, 250, 500, 1000, 1500, 2000, 3000, 4000, 6000, 8000 | 10 | Primary | HAC5 | 0.748047 | 0.631759 | 0.832607 | 0.863836 | Partial recovery |
| Münster, all timepoints | Perturbation sensitivity | 125, 250, 500, 1000, 1500, 2000, 3000, 4000, 6000, 8000 | 10 | Primary | HAC6 | 0.943634 | 0.701905 | 0.832607 | 0.844731 | Partial recovery |
| Münster, all timepoints | Perturbation sensitivity | 125, 250, 500, 1000, 1500, 2000, 3000, 4000, 6000, 8000 | 10 | De-levelled | HAC2 | 0.928636 | 0.899439 | 0.822679 | 0.995961 | Partial recovery |
| Münster, all timepoints | Perturbation sensitivity | 125, 250, 500, 1000, 1500, 2000, | 10 | De-levelled | HAC3 | 0.714987 | 0.657493 | 0.822679 | 0.979228 | Partial recovery |

| Grid variant | Role | Grid (Hz) | Tones | Arm | HAC axis | Congruence | 2.5% lower | Null 99.9% | Split-half | Result |
|---|---|---|---|---|---|---|---|---|---|---|
| | | 3000, 4000, 6000, 8000 | | | | | | | | |
| Münster, all timepoints | Perturbation sensitivity | 125, 250, 500, 1000, 1500, 2000, 3000, 4000, 6000, 8000 | 10 | De-levelled | HAC4 | 0.659535 | 0.560788 | 0.822679 | 0.941058 | Partial recovery |
| Münster, all timepoints | Perturbation sensitivity | 125, 250, 500, 1000, 1500, 2000, 3000, 4000, 6000, 8000 | 10 | De-levelled | HAC5 | 0.746425 | 0.635985 | 0.822679 | 0.852398 | Partial recovery |
| Münster, all timepoints | Perturbation sensitivity | 125, 250, 500, 1000, 1500, 2000, 3000, 4000, 6000, 8000 | 10 | De-levelled | HAC6 | 0.944489 | 0.702141 | 0.822679 | 0.848269 | Partial recovery |
| Münster, hour 0 | Primary baseline | 125, 250, 500, 1000, 1500, 2000, 3000, 4000, 6000, 8000 | 10 | Primary | HAC2 | 0.983133 | 0.962193 | 0.832607 | 0.992693 | Axis inconclusive |
| Münster, hour 0 | Primary baseline | 125, 250, 500, 1000, 1500, 2000, 3000, 4000, 6000, 8000 | 10 | Primary | HAC3 | 0.785704 | 0.672293 | 0.832607 | 0.874854 | Axis inconclusive |
| Münster, hour 0 | Primary baseline | 125, 250, 500, 1000, 1500, 2000, 3000, 4000, 6000, 8000 | 10 | Primary | HAC4 | 0.647438 | 0.551141 | 0.832607 | 0.801212 | Axis inconclusive |
| Münster, hour 0 | Primary baseline | 125, 250, 500, 1000, 1500, 2000, 3000, 4000, 6000, 8000 | 10 | Primary | HAC5 | 0.642359 | 0.588319 | 0.832607 | 0.856518 | Axis inconclusive |
| Münster, hour 0 | Primary baseline | 125, 250, 500, 1000, 1500, 2000, 3000, 4000, 6000, 8000 | 10 | Primary | HAC6 | 0.739908 | 0.653505 | 0.832607 | 0.780826 | Axis inconclusive |
| Münster, hour 0 | Primary baseline | 125, 250, 500, 1000, 1500, 2000, 3000, 4000, 6000, 8000 | 10 | De-levelled | HAC2 | 0.941006 | 0.914057 | 0.822679 | 0.995194 | Axis inconclusive |
| Münster, hour 0 | Primary baseline | 125, 250, 500, 1000, 1500, 2000, 3000, 4000, 6000, 8000 | 10 | De-levelled | HAC3 | 0.775565 | 0.671688 | 0.822679 | 0.877162 | Axis inconclusive |
| Münster, hour 0 | Primary baseline | 125, 250, 500, 1000, 1500, 2000, 3000, 4000, 6000, 8000 | 10 | De-levelled | HAC4 | 0.649450 | 0.543978 | 0.822679 | 0.785656 | Axis inconclusive |
| Münster, hour 0 | Primary baseline | 125, 250, 500, 1000, 1500, 2000, 3000, 4000, 6000, 8000 | 10 | De-levelled | HAC5 | 0.641853 | 0.579641 | 0.822679 | 0.836017 | Axis inconclusive |
| Münster, hour 0 | Primary baseline | 125, 250, 500, 1000, 1500, 2000, 3000, 4000, 6000, 8000 | 10 | De-levelled | HAC6 | 0.745812 | 0.648863 | 0.822679 | 0.751540 | Axis inconclusive |
| Münster, hour 0, canonical six | Resolution control | 500, 1000, 2000, 3000, 4000, 6000 | 6 | Primary | HAC2 | 0.969909 | 0.952061 | 0.813498 | 0.994864 | Partial recovery |
| Münster, hour 0, canonical six | Resolution control | 500, 1000, 2000, 3000, 4000, 6000 | 6 | Primary | HAC3 | 0.838796 | 0.757802 | 0.813498 | 0.985563 | Partial recovery |

| Grid variant | Role | Grid (Hz) | Tones | Arm | HAC axis | Congruence | 2.5% lower | Null 99.9% | Split-half | Result |
|---|---|---|---|---|---|---|---|---|---|---|
| Münster, hour 0, canonical six | Resolution control | 500, 1000, 2000, 3000, 4000, 6000 | 6 | Primary | HAC4 | 0.662705 | 0.557944 | 0.813498 | 0.975417 | Partial recovery |
| Münster, hour 0, canonical six | Resolution control | 500, 1000, 2000, 3000, 4000, 6000 | 6 | Primary | HAC5 | 0.823091 | 0.572664 | 0.813498 | 0.866832 | Partial recovery |
| Münster, hour 0, canonical six | Resolution control | 500, 1000, 2000, 3000, 4000, 6000 | 6 | Primary | HAC6 | 0.805195 | 0.630038 | 0.813498 | 0.865505 | Partial recovery |
| Münster, hour 0, canonical six | Resolution control | 500, 1000, 2000, 3000, 4000, 6000 | 6 | De-levelled | HAC2 | 0.980498 | 0.962996 | 0.770236 | 0.996983 | Partial recovery |
| Münster, hour 0, canonical six | Resolution control | 500, 1000, 2000, 3000, 4000, 6000 | 6 | De-levelled | HAC3 | 0.799101 | 0.656910 | 0.770236 | 0.985738 | Partial recovery |
| Münster, hour 0, canonical six | Resolution control | 500, 1000, 2000, 3000, 4000, 6000 | 6 | De-levelled | HAC4 | 0.593315 | 0.498559 | 0.770236 | 0.974711 | Partial recovery |
| Münster, hour 0, canonical six | Resolution control | 500, 1000, 2000, 3000, 4000, 6000 | 6 | De-levelled | HAC5 | 0.835456 | 0.551788 | 0.770236 | 0.888084 | Partial recovery |
| Münster, hour 0, canonical six | Resolution control | 500, 1000, 2000, 3000, 4000, 6000 | 6 | De-levelled | HAC6 | 0.799867 | 0.602350 | 0.770236 | 0.888884 | Partial recovery |
| Ladder, OHHR-like grid | Resolution ladder | 125, 250, 500, 1000, 1500, 2000, 4000 | 7 | Primary | HAC2 | 0.812018 | 0.748605 | 0.907812 | 0.988243 | No recovery |
| Ladder, OHHR-like grid | Resolution ladder | 125, 250, 500, 1000, 1500, 2000, 4000 | 7 | Primary | HAC4 | 0.775959 | 0.714570 | 0.907812 | 0.973549 | No recovery |
| Ladder, OHHR-like grid | Resolution ladder | 125, 250, 500, 1000, 1500, 2000, 4000 | 7 | Primary | HAC6 | 0.826165 | 0.692646 | 0.907812 | 0.950140 | No recovery |
| Ladder, OHHR-like grid | Resolution ladder | 125, 250, 500, 1000, 1500, 2000, 4000 | 7 | De-levelled | HAC2 | 0.854393 | 0.811831 | 0.898233 | 0.990639 | No recovery |
| Ladder, OHHR-like grid | Resolution ladder | 125, 250, 500, 1000, 1500, 2000, 4000 | 7 | De-levelled | HAC4 | 0.779069 | 0.721567 | 0.898233 | 0.977442 | No recovery |
| Ladder, OHHR-like grid | Resolution ladder | 125, 250, 500, 1000, 1500, 2000, 4000 | 7 | De-levelled | HAC6 | 0.823102 | 0.685112 | 0.898233 | 0.954661 | No recovery |
| Ladder, full ten tone | Resolution ladder | 125, 250, 500, 1000, 1500, 2000, 3000, 4000, 6000, 8000 | 10 | Primary | HAC2 | 0.983133 | 0.964227 | 0.832607 | 0.992266 | Axis inconclusive |
| Ladder, full ten tone | Resolution ladder | 125, 250, 500, 1000, 1500, 2000, 3000, 4000, 6000, 8000 | 10 | Primary | HAC3 | 0.785704 | 0.672048 | 0.832607 | 0.885775 | Axis inconclusive |
| Ladder, full ten tone | Resolution ladder | 125, 250, 500, 1000, 1500, 2000, 3000, 4000, 6000, 8000 | 10 | Primary | HAC4 | 0.647438 | 0.558512 | 0.832607 | 0.822385 | Axis inconclusive |
| Ladder, full ten tone | Resolution ladder | 125, 250, 500, 1000, 1500, 2000, 3000, 4000, 6000, 8000 | 10 | Primary | HAC5 | 0.642359 | 0.587483 | 0.832607 | 0.848739 | Axis inconclusive |
| Ladder, full ten tone | Resolution ladder | 125, 250, 500, 1000, 1500, 2000, 3000, 4000, 6000, 8000 | 10 | Primary | HAC6 | 0.739908 | 0.650321 | 0.832607 | 0.794394 | Axis inconclusive |
| Ladder, full ten tone | Resolution ladder | 125, 250, 500, 1000, 1500, 2000, 3000, 4000, 6000, 8000 | 10 | De-levelled | HAC2 | 0.941006 | 0.914918 | 0.822679 | 0.995607 | Axis inconclusive |

| Grid variant | Role | Grid (Hz) | Tones | Arm | HAC axis | Congruence | 2.5% lower | Null 99.9% | Split-half | Result |
|---|---|---|---|---|---|---|---|---|---|---|
| Ladder, full ten tone | Resolution ladder | 125, 250, 500, 1000, 1500, 2000, 3000, 4000, 6000, 8000 | 10 | De-levelled | HAC3 | 0.775565 | 0.670445 | 0.822679 | 0.885143 | Axis inconclusive |
| Ladder, full ten tone | Resolution ladder | 125, 250, 500, 1000, 1500, 2000, 3000, 4000, 6000, 8000 | 10 | De-levelled | HAC4 | 0.649450 | 0.548522 | 0.822679 | 0.820430 | Axis inconclusive |
| Ladder, full ten tone | Resolution ladder | 125, 250, 500, 1000, 1500, 2000, 3000, 4000, 6000, 8000 | 10 | De-levelled | HAC5 | 0.641853 | 0.576462 | 0.822679 | 0.828875 | Axis inconclusive |
| Ladder, full ten tone | Resolution ladder | 125, 250, 500, 1000, 1500, 2000, 3000, 4000, 6000, 8000 | 10 | De-levelled | HAC6 | 0.745812 | 0.642391 | 0.822679 | 0.797060 | Axis inconclusive |
| Ladder, plus 3 kHz | Resolution ladder | 125, 250, 500, 1000, 1500, 2000, 3000, 4000 | 8 | Primary | HAC2 | 0.794272 | 0.740421 | 0.905003 | 0.987628 | No recovery |
| Ladder, plus 3 kHz | Resolution ladder | 125, 250, 500, 1000, 1500, 2000, 3000, 4000 | 8 | Primary | HAC4 | 0.750973 | 0.680049 | 0.905003 | 0.959951 | No recovery |
| Ladder, plus 3 kHz | Resolution ladder | 125, 250, 500, 1000, 1500, 2000, 3000, 4000 | 8 | Primary | HAC6 | 0.828579 | 0.652643 | 0.905003 | 0.869596 | No recovery |
| Ladder, plus 3 kHz | Resolution ladder | 125, 250, 500, 1000, 1500, 2000, 3000, 4000 | 8 | De-levelled | HAC2 | 0.840095 | 0.791249 | 0.895415 | 0.989834 | No recovery |
| Ladder, plus 3 kHz | Resolution ladder | 125, 250, 500, 1000, 1500, 2000, 3000, 4000 | 8 | De-levelled | HAC4 | 0.755194 | 0.686221 | 0.895415 | 0.968738 | No recovery |
| Ladder, plus 3 kHz | Resolution ladder | 125, 250, 500, 1000, 1500, 2000, 3000, 4000 | 8 | De-levelled | HAC6 | 0.831728 | 0.657595 | 0.895415 | 0.868471 | No recovery |
| Ladder, plus 3 and 6 kHz | Resolution ladder | 125, 250, 500, 1000, 1500, 2000, 3000, 4000, 6000 | 9 | Primary | HAC2 | 0.949561 | 0.918563 | 0.830004 | 0.992595 | Partial recovery |
| Ladder, plus 3 and 6 kHz | Resolution ladder | 125, 250, 500, 1000, 1500, 2000, 3000, 4000, 6000 | 9 | Primary | HAC3 | 0.941687 | 0.895644 | 0.830004 | 0.969296 | Partial recovery |
| Ladder, plus 3 and 6 kHz | Resolution ladder | 125, 250, 500, 1000, 1500, 2000, 3000, 4000, 6000 | 9 | Primary | HAC4 | 0.657177 | 0.583318 | 0.830004 | 0.950667 | Partial recovery |
| Ladder, plus 3 and 6 kHz | Resolution ladder | 125, 250, 500, 1000, 1500, 2000, 3000, 4000, 6000 | 9 | Primary | HAC5 | 0.636440 | 0.582175 | 0.830004 | 0.851107 | Partial recovery |
| Ladder, plus 3 and 6 kHz | Resolution ladder | 125, 250, 500, 1000, 1500, 2000, 3000, 4000, 6000 | 9 | Primary | HAC6 | 0.738011 | 0.648128 | 0.830004 | 0.817312 | Partial recovery |
| Ladder, plus 3 and 6 kHz | Resolution ladder | 125, 250, 500, 1000, 1500, 2000, 3000, 4000, 6000 | 9 | De-levelled | HAC2 | 0.874958 | 0.843098 | 0.812091 | 0.995526 | Partial recovery |
| Ladder, plus 3 and 6 kHz | Resolution ladder | 125, 250, 500, 1000, 1500, 2000, 3000, 4000, 6000 | 9 | De-levelled | HAC3 | 0.941768 | 0.896114 | 0.812091 | 0.971760 | Partial recovery |
| Ladder, plus 3 and 6 kHz | Resolution ladder | 125, 250, 500, 1000, 1500, 2000, 3000, 4000, 6000 | 9 | De-levelled | HAC4 | 0.658704 | 0.584077 | 0.812091 | 0.948862 | Partial recovery |

| Grid variant | Role | Grid (Hz) | Tones | Arm | HAC axis | Congruence | 2.5% lower | Null 99.9% | Split-half | Result |
|---|---|---|---|---|---|---|---|---|---|---|
| Ladder, plus 3 and 6 kHz | Resolution ladder | 125, 250, 500, 1000, 1500, 2000, 3000, 4000, 6000 | 9 | De-levelled | HAC5 | 0.636383 | 0.583787 | 0.812091 | 0.801329 | Partial recovery |
| Ladder, plus 3 and 6 kHz | Resolution ladder | 125, 250, 500, 1000, 1500, 2000, 3000, 4000, 6000 | 9 | De-levelled | HAC6 | 0.743512 | 0.647925 | 0.812091 | 0.810558 | Partial recovery |
| Ladder, plus 6 kHz | Resolution ladder | 125, 250, 500, 1000, 1500, 2000, 4000, 6000 | 8 | Primary | HAC2 | 0.884665 | 0.830465 | 0.866892 | 0.992425 | No recovery |
| Ladder, plus 6 kHz | Resolution ladder | 125, 250, 500, 1000, 1500, 2000, 4000, 6000 | 8 | Primary | HAC4 | 0.676764 | 0.620768 | 0.866892 | 0.989188 | No recovery |
| Ladder, plus 6 kHz | Resolution ladder | 125, 250, 500, 1000, 1500, 2000, 4000, 6000 | 8 | Primary | HAC6 | 0.816177 | 0.702206 | 0.866892 | 0.937803 | No recovery |
| Ladder, plus 6 kHz | Resolution ladder | 125, 250, 500, 1000, 1500, 2000, 4000, 6000 | 8 | De-levelled | HAC2 | 0.759723 | 0.730945 | 0.861280 | 0.988307 | No recovery |
| Ladder, plus 6 kHz | Resolution ladder | 125, 250, 500, 1000, 1500, 2000, 4000, 6000 | 8 | De-levelled | HAC4 | 0.656335 | 0.565945 | 0.861280 | 0.956626 | No recovery |
| Ladder, plus 6 kHz | Resolution ladder | 125, 250, 500, 1000, 1500, 2000, 4000, 6000 | 8 | De-levelled | HAC6 | 0.812856 | 0.699347 | 0.861280 | 0.929873 | No recovery |
| Ladder, plus 8 kHz | Resolution ladder | 125, 250, 500, 1000, 1500, 2000, 4000, 8000 | 8 | Primary | HAC2 | 0.897642 | 0.844544 | 0.879727 | 0.992154 | No recovery |
| Ladder, plus 8 kHz | Resolution ladder | 125, 250, 500, 1000, 1500, 2000, 4000, 8000 | 8 | Primary | HAC4 | 0.767718 | 0.716345 | 0.879727 | 0.963203 | No recovery |
| Ladder, plus 8 kHz | Resolution ladder | 125, 250, 500, 1000, 1500, 2000, 4000, 8000 | 8 | Primary | HAC6 | 0.802409 | 0.689037 | 0.879727 | 0.950752 | No recovery |
| Ladder, plus 8 kHz | Resolution ladder | 125, 250, 500, 1000, 1500, 2000, 4000, 8000 | 8 | De-levelled | HAC2 | 0.753675 | 0.720212 | 0.870801 | 0.994519 | No recovery |
| Ladder, plus 8 kHz | Resolution ladder | 125, 250, 500, 1000, 1500, 2000, 4000, 8000 | 8 | De-levelled | HAC4 | 0.769151 | 0.707835 | 0.870801 | 0.963936 | No recovery |
| Ladder, plus 8 kHz | Resolution ladder | 125, 250, 500, 1000, 1500, 2000, 4000, 8000 | 8 | De-levelled | HAC6 | 0.797531 | 0.679450 | 0.870801 | 0.949719 | No recovery |

These rows use the same underlying cohort under different frequency grids and are resolution experiments, not independent population replications.

*Panel d-iii. Rank-limited four-tone controls.*

| Analysis | Reference axis | Matched axis | Congruence | 2.5% lower | Null 99.9% | Split-half | Result |
|---|---|---|---|---|---|---|---|
| NIOSH four-tone resolution control | C1 | A1 | 0.999692 | 0.999679 | — | 0.999999 | Not evaluable: rank limit |
| NIOSH four-tone resolution control | C2 | A3 | 0.999861 | 0.999802 | — | 0.999997 | Not evaluable: rank limit |
| NIOSH four-tone resolution control | C3 | A6 | 0.930223 | 0.928823 | — | 0.999997 | Not evaluable: rank limit |
| NIOSH four-tone resolution control | C4 | A7 | 0.999380 | 0.999348 | — | 0.999999 | Not evaluable: rank limit |
| NHANES I | C1 | A1 | 0.969503 | 0.964735 | 0.956292 | 0.999866 | Partial four-tone signature |
| NHANES I | C2 | A2 | 0.936845 | 0.927368 | 0.956292 | 0.999499 | Partial four-tone signature |
| NHANES I | C3 | A3 | 0.967192 | 0.959919 | 0.956292 | 0.999330 | Partial four-tone signature |

| Analysis | Reference axis | Matched axis | Congruence | 2.5% lower | Null 99.9% | Split-half | Result |
|---|---|---|---|---|---|---|---|
| NHANES I | C4 | A4 | 0.999350 | 0.998086 | 0.956292 | 0.999685 | Partial four-tone signature |
| NHANES I | E1 | A1 | 0.984111 | 0.979560 | 0.996174 | 0.999826 | Partial four-tone signature |
| NHANES I | E2 | A2 | 0.984448 | 0.979906 | 0.996174 | 0.999525 | Partial four-tone signature |
| NHANES I | E3 | A3 | 0.999312 | 0.998256 | 0.996174 | 0.999745 | Partial four-tone signature |
| NHANES II | C1 | A1 | 0.842526 | 0.829112 | 0.956292 | 0.999686 | Partial four-tone signature |
| NHANES II | C2 | A2 | 0.736544 | 0.676502 | 0.956292 | 0.996367 | Partial four-tone signature |
| NHANES II | C3 | A3 | 0.885005 | 0.832072 | 0.956292 | 0.994355 | Partial four-tone signature |
| NHANES II | C4 | A4 | 0.996843 | 0.989076 | 0.956292 | 0.998255 | Partial four-tone signature |
| NHANES II | E1 | A1 | 0.963746 | 0.927289 | 0.996174 | 0.996920 | No four-tone signature |
| NHANES II | E2 | A2 | 0.962578 | 0.924530 | 0.996174 | 0.994672 | No four-tone signature |
| NHANES II | E3 | A3 | 0.998139 | 0.991748 | 0.996174 | 0.998250 | No four-tone signature |
| Hispanic HANES | C1 | A1 | 0.939597 | 0.932140 | 0.956292 | 0.999828 | Partial four-tone signature |
| Hispanic HANES | C2 | A2 | 0.904057 | 0.891514 | 0.956292 | 0.999327 | Partial four-tone signature |
| Hispanic HANES | C3 | A3 | 0.958340 | 0.949312 | 0.956292 | 0.998773 | Partial four-tone signature |
| Hispanic HANES | C4 | A4 | 0.995348 | 0.990870 | 0.956292 | 0.999492 | Partial four-tone signature |
| Hispanic HANES | E1 | A1 | 0.993351 | 0.989460 | 0.996174 | 0.999620 | No four-tone signature |
| Hispanic HANES | E2 | A2 | 0.989701 | 0.984085 | 0.996174 | 0.999077 | No four-tone signature |
| Hispanic HANES | E3 | A3 | 0.995889 | 0.991380 | 0.996174 | 0.999583 | No four-tone signature |
| Pooled NHANES I and II | C1 | A1 | 0.972308 | 0.968777 | 0.956292 | 0.999907 | Partial four-tone signature |
| Pooled NHANES I and II | C2 | A2 | 0.930481 | 0.921430 | 0.956292 | 0.999658 | Partial four-tone signature |
| Pooled NHANES I and II | C3 | A3 | 0.957950 | 0.950544 | 0.956292 | 0.999430 | Partial four-tone signature |
| Pooled NHANES I and II | C4 | A4 | 0.999307 | 0.997952 | 0.956292 | 0.999784 | Partial four-tone signature |
| Pooled NHANES I and II | E1 | A1 | 0.984270 | 0.980408 | 0.996174 | 0.999891 | Partial four-tone signature |
| Pooled NHANES I and II | E2 | A2 | 0.984246 | 0.980351 | 0.996174 | 0.999645 | Partial four-tone signature |
| Pooled NHANES I and II | E3 | A3 | 0.999470 | 0.998363 | 0.996174 | 0.999806 | Partial four-tone signature |
| Pooled, all three four-tone surveys | C1 | A1 | 0.966568 | 0.963027 | 0.956292 | 0.999922 | Partial four-tone signature |
| Pooled, all three four-tone surveys | C2 | A2 | 0.922772 | 0.915775 | 0.956292 | 0.999770 | Partial four-tone signature |
| Pooled, all three four-tone surveys | C3 | A3 | 0.955279 | 0.949626 | 0.956292 | 0.999639 | Partial four-tone signature |
| Pooled, all three four-tone surveys | C4 | A4 | 0.998703 | 0.997294 | 0.956292 | 0.999815 | Partial four-tone signature |
| Pooled, all three four-tone surveys | E1 | A1 | 0.986078 | 0.983117 | 0.996174 | 0.999907 | Partial four-tone signature |
| Pooled, all three four-tone surveys | E2 | A2 | 0.985434 | 0.982211 | 0.996174 | 0.999670 | Partial four-tone signature |
| Pooled, all three four-tone surveys | E3 | A3 | 0.998987 | 0.997761 | 0.996174 | 0.999859 | Partial four-tone signature |

Four observed dimensions cannot identify five axes, so no Atlas recovery verdict is issued for the within-NIOSH resolution control. The historical four-tone surveys are reported against their own signature axes, labelled C1 and upward, and are not five-axis Atlas recoveries.

*Panel e. Transport of the coordinate frame derived from NIOSH alone, applied unchanged with no target fitting, centring, rotation or rematching.*

| Receiving cohort | HAC axis | Pearson r | 2.5% lower | 97.5% upper | Null 99.9% | Exceeds null |
|---|---|---|---|---|---|---|
| NHES Cycle II | HAC1 | 0.941504 | 0.937570 | 0.945429 | Not applicable | Not applicable |
| NHES Cycle II | HAC2 | 0.922066 | 0.917497 | 0.926550 | 0.598387 | Clears null |
| NHES Cycle II | HAC3 | 0.738789 | 0.727541 | 0.750438 | 0.598387 | Clears null |
| NHES Cycle II | HAC4 | 0.913216 | 0.909359 | 0.916891 | 0.598387 | Clears null |
| NHES Cycle II | HAC5 | 0.806234 | 0.797700 | 0.814843 | 0.598387 | Clears null |
| NHES Cycle II | HAC6 | 0.931613 | 0.928706 | 0.934561 | 0.598387 | Clears null |
| NHES Cycle III | HAC1 | 0.941033 | 0.937022 | 0.944744 | Not applicable | Not applicable |
| NHES Cycle III | HAC2 | 0.937798 | 0.933534 | 0.941570 | 0.583687 | Clears null |
| NHES Cycle III | HAC3 | 0.715941 | 0.704578 | 0.727628 | 0.583687 | Clears null |

| Receiving cohort | HAC axis | Pearson r | 2.5% lower | 97.5% upper | Null 99.9% | Exceeds null |
|---|---|---|---|---|---|---|
| NHES Cycle III | HAC4 | 0.909338 | 0.905195 | 0.913300 | 0.583687 | Clears null |
| NHES Cycle III | HAC5 | 0.796999 | 0.788771 | 0.805330 | 0.583687 | Clears null |
| NHES Cycle III | HAC6 | 0.925493 | 0.922574 | 0.928301 | 0.583687 | Clears null |
| Münster/Basel | HAC1 | 0.964174 | 0.957324 | 0.970202 | Not applicable | Not applicable |
| Münster/Basel | HAC2 | 0.978716 | 0.973316 | 0.983113 | 0.532940 | Clears null |
| Münster/Basel | HAC3 | 0.663617 | 0.597732 | 0.719803 | 0.532940 | Clears null |
| Münster/Basel | HAC4 | 0.924532 | 0.908856 | 0.937787 | 0.532940 | Clears null |
| Münster/Basel | HAC5 | 0.759396 | 0.714228 | 0.798167 | 0.532940 | Clears null |
| Münster/Basel | HAC6 | 0.920127 | 0.903997 | 0.933888 | 0.532940 | Clears null |

Values are Pearson correlations between transported and Atlas scores. HAC1 carries no rotation null; those three comparisons are marked not applicable and are reported for their correlation alone.

#### Supplementary Table S5. Reconstruction and decomposition of longitudinal hearing change in repeated same-ear audiograms

*The frozen coordinates, derived from cross-sectional population audiograms, applied to the change observed between successive examinations of the same ear. No longitudinal representation was fitted.*

| Coordinates | Number | MAE, dB | 95% CI | Threshold changes within 5 dB, % | Change variance reconstructed, % |
|---|---|---|---|---|---|
| dHAC1 | 1 | 4.2823 | 4.2775 to 4.2869 | 71.644 | 39.646 |
| dHAC1-dHAC2 | 2 | 3.5740 | 3.5702 to 3.5778 | 75.496 | 58.772 |
| dHAC1-dHAC3 | 3 | 2.5310 | 2.5278 to 2.5339 | 86.355 | 77.935 |
| dHAC1-dHAC4 | 4 | 1.8176 | 1.8152 to 1.8201 | 93.220 | 87.382 |
| dHAC1-dHAC5 | 5 | 1.0148 | 1.0132 to 1.0164 | 98.211 | 95.321 |
| dHAC1-dHAC6 | 6 | 0.0000 | exact | 100.000 | 100.000 |

Intervals are 95% percentile intervals from worker-clustered bootstrap resamples.

*Panel b. Decomposition of observed longitudinal movement and its correspondence with between-person variation.*

| Quantity | Value |
|---|---|
| Successive same-ear comparisons | 1,361,350 edges from 446,146 workers |
| Movement along overall hearing level | 42.839% (95% CI 42.642 to 43.045) |
| Movement in audiogram shape | 57.161% |
| Edges contributing to the level and shape partition | 1,358,368 from 444,820 workers |
| Between-person and within-ear leading subspace overlap | 0.9202 |
| Random-orientation null, median | 0.403 |
| Random-orientation null, 95th percentile | 0.663 |
| Monte Carlo P | 0.0005 |
| Edges contributing to the subspace comparison | 1,294,932 from 421,868 workers |

The level and shape partition is computed in the isometric embedding, in which coordinate distance equals audiogram distance in decibels. Overall hearing level is removed before the subspace comparison.

#### Supplementary Table S6. Dataset inventory of the Human Hearing Atlas

*Every dataset analysed, ordered by cohort family and survey year. Counts are post-QC, after cleaning and harmonisation, and need not match the raw deposits.*

| Dataset | Country | Years | Role in this paper | Design | Cohort | Frequency grid, kHz unless stated | Reference standard | Participants | Ear-curves |
|---|---|---|---|---|---|---|---|---|---|
| NHES Cycle II | United States | 1963-1965 | Historical external validation | Cross-sectional | Population | 8: 250, 500, 1k, 2k, 3k, 4k, 6k, 8k | ASA-1951 ‡ | 7,092 | 14,184 |
| NHES Cycle III | United States | 1966-1970 | Historical external validation | Cross-sectional | Population | 8: 250, 500, 1k, 2k, 3k, 4k, 6k, 8k | ASA-1951 ‡ | 6,764 | 13,528 |

| Dataset | Country | Years | Role in this paper | Design | Cohort | Frequency grid, kHz unless stated | Reference standard | Participants | Ear-curves |
|---|---|---|---|---|---|---|---|---|---|
| NHANES I | United States | 1971-1974 | External validation | Cross-sectional | Population | 4: 500, 1k, 2k, 4k | ISO/ANSI | 6,713 | 13,426 |
| NHANES II | United States | 1976-1980 | External validation | Cross-sectional | Population | 4: 500, 1k, 2k, 4k | ISO/ANSI | 5,692 | 11,384 |
| Hispanic HANES | United States | 1982-1984 | External validation | Cross-sectional | Population | 4: 500, 1k, 2k, 4k | ISO/ANSI | 6,931 | 13,862 |
| NHANES 1999-2000 | United States | 1999-2000 | Canonical derivation | Cross-sectional | Population | 7: 500, 1k, 2k, 3k, 4k, 6k, 8k | ISO/ANSI | 1,655 | 3,310 |
| NHANES 2001-2002 | United States | 2001-2002 | Canonical derivation | Cross-sectional | Population | 7: 500, 1k, 2k, 3k, 4k, 6k, 8k | ISO/ANSI | 1,861 | 3,722 |
| NHANES 2003-2004 | United States | 2003-2004 | Canonical derivation | Cross-sectional | Population | 7: 500, 1k, 2k, 3k, 4k, 6k, 8k | ISO/ANSI | 1,750 | 3,500 |
| NHANES 2005-2006 | United States | 2005-2006 | Canonical derivation | Cross-sectional | Population | 7: 500, 1k, 2k, 3k, 4k, 6k, 8k | ISO/ANSI | 2,663 | 5,326 |
| NHANES 2007-2008 | United States | 2007-2008 | Canonical derivation | Cross-sectional | Population | 7: 500, 1k, 2k, 3k, 4k, 6k, 8k | ISO/ANSI | 1,134 | 2,268 |
| NHANES 2009-2010 | United States | 2009-2010 | Canonical derivation | Cross-sectional | Population | 7: 500, 1k, 2k, 3k, 4k, 6k, 8k | ISO/ANSI | 2,111 | 4,222 |
| NHANES 2011-2012 | United States | 2011-2012 | Canonical derivation | Cross-sectional | Population | 7: 500, 1k, 2k, 3k, 4k, 6k, 8k | ISO/ANSI | 3,818 | 7,636 |
| NHANES 2015-2016 | United States | 2015-2016 | Canonical derivation | Cross-sectional | Population | 7: 500, 1k, 2k, 3k, 4k, 6k, 8k | ISO/ANSI | 4,263 | 8,526 |
| NHANES 2017-2020 | United States | 2017-2020 | Canonical derivation | Cross-sectional | Population | 7: 500, 1k, 2k, 3k, 4k, 6k, 8k | ISO/ANSI | 3,837 | 7,674 |
| KNHANES 2009 | South Korea | 2009 | Canonical derivation | Cross-sectional | Population | 6: 500, 1k, 2k, 3k, 4k, 6k | ISO/ANSI | 3,780 | 7,560 |
| KNHANES 2010 | South Korea | 2010 | Canonical derivation | Cross-sectional | Population | 6: 500, 1k, 2k, 3k, 4k, 6k | ISO/ANSI | 6,320 | 12,640 |
| KNHANES 2011 | South Korea | 2011 | Canonical derivation | Cross-sectional | Population | 6: 500, 1k, 2k, 3k, 4k, 6k | ISO/ANSI | 6,092 | 12,184 |
| KNHANES 2012 | South Korea | 2012 | Canonical derivation | Cross-sectional | Population | 6: 500, 1k, 2k, 3k, 4k, 6k | ISO/ANSI | 5,592 | 11,184 |
| KNHANES 2019 | South Korea | 2019 | Held-out external validation | Cross-sectional | Population | 5: 500, 1k, 2k, 4k, 8k | ISO/ANSI | 1,830 | 3,660 |
| KNHANES 2020 | South Korea | 2020 | Held-out external validation | Cross-sectional | Population | 5: 500, 1k, 2k, 4k, 8k | ISO/ANSI | 3,682 | 7,364 |
| KNHANES 2021 | South Korea | 2021 | Held-out external validation | Cross-sectional | Population | 5: 500, 1k, 2k, 4k, 8k | ISO/ANSI | 3,564 | 7,128 |
| KNHANES 2022 | South Korea | 2022 | Held-out external validation | Cross-sectional | Population | 5: 500, 1k, 2k, 4k, 8k | ISO/ANSI | 4,978 | 9,956 |
| KNHANES 2023 | South Korea | 2023 | Held-out external validation | Cross-sectional | Population | 5: 500, 1k, 2k, 4k, 8k | ISO/ANSI | 4,854 | 9,708 |
| OHHR | Germany | 2013-2015 | External validation | Cross-sectional | Clinical | 8: 125, 250, 500, 750, 1k, 1.5k, 2k, 4k | ISO/ANSI | 569 | 1,138 |
| BEAR | Denmark | 2017-2018 | External validation | Cross-sectional | Clinical | 6: 250, 500, 1k, 2k, 4k, 8k | ISO/ANSI | 86 | 172 |
| NIOSH OHL Surveillance | United States | 1981-2010 | Longitudinal external validation | Longitudinal | Occupational | 7: 500, 1k, 2k, 3k, 4k, 6k, 8k | ISO/ANSI | 1,365,737 † | 4,092,824 † |
| Münster glycerol series | Germany | 2013 * | Acute longitudinal validation | Longitudinal, hours 0 to 4 | Clinical | 10: 125, 250, 500, 1k, 1.5k, 2k, 3k, 4k, 6k, 8k | ISO/ANSI | — ¶ | 1,778 ¶ |
| Iranian clinical cohort | Iran | 2023 | External validation | Cross-sectional | Clinical | 6: 250, 500, 1k, 2k, 4k, 8k ‖ | ISO/ANSI | 495 ‖ | 962 ‖ |
| Meltem Hospital | Türkiye | 2024 * | External validation | Cross-sectional | Clinical | 6: 250, 500, 1k, 2k, 4k, 8k | ISO/ANSI | 324 | 648 |
| Cadenza CAD1 listener panel | United Kingdom | 2023 * | External validation | Cross-sectional | Experimental | 8: 250, 500, 1k, 2k, 3k, 4k, 6k, 8k | ISO/ANSI | 53 | 106 |
| Macquarie (Izmaylova) # | Australia | 2024 * | External validation | Cross-sectional | Experimental | 7: 125, 250, 500, 1k, 2k, 4k, 8k | ISO/ANSI | 121 | 242 |
| Regev DTU deposit 25134611 | Denmark | 2024 * | External validation | Cross-sectional | Experimental | 9: 125, 250, 500, 1k, 2k, 3k, 4k, 6k, 8k | ISO/ANSI | 13 | 13 § |
| Regev DTU deposit 25771884 | Denmark | 2025 * | External validation | Cross-sectional | Experimental | 9: 125, 250, 500, 1k, 2k, 3k, 4k, 6k, 8k | ISO/ANSI | 28 | 28 § |
| Total, 33 datasets | 8 countries | 1963-2025 | | | 4 cohort types | | | 1,464,402 | 4,291,863 |

Derivation: 13 datasets, 2 countries, 44,876 participants, 89,752 ear-curves. Non-derivation: 20 datasets, 8 countries, 1,419,526 participants, 4,202,111 ear-curves. Twenty-three of the 33 are population survey waves; the remainder are clinical, occupational and experimental cohorts.

Independent-recovery or transport analyses additionally used the occupational archive, NHES Cycles II and III, the held-out KNHANES waves, OHHR, the Münster glycerol series, the Iranian cohort and Meltem Hospital.

No total is given for sessions or examinations, which are not additive across cohort types.

* Sample size after quality control, cleaning and harmonisation, which may not reflect the original raw dataset; in Years, a single reported year is the dataset deposit or release.

‡ ASA-1951 thresholds converted frequency by frequency to ANSI-1969 at decoding.

† The occupational archive contributes 1,365,737 workers, 2,046,412 examinations and 4,092,824 ear-examinations. Repeated ear-examinations are observations, not unique ears.

¶ The glycerol series contributes 1,778 serial ear-observations from 356 ear-series in 347 patients, each ear measured before and 1, 2, 3 and 4 hours after administration. Ear-series carry no patient linkage in the deposited data, so no participant count is entered and the series contributes none to the participant total.

### Original separate left- and right-ear thresholds provided directly by Tatiana Izmaylova; the public Dryad release contains participant-level bilateral averages.

§ Participant counts; no ear-level count is available for these two deposits.

‖ Dataset-level denominator: the cohort of 962 ear-charts from 495 participants on the grid shown, one session per participant; the source dataset contained 1,004 charts from 502 participants. The measured clinical grid omits 3 and 6 kHz, two of the six canonical frequencies, so analyses requiring the canonical six use a smaller eligible subset reported in Supplementary Table S3. Coordinates requiring unmeasured frequencies were treated as unavailable and were not imputed.

*Supplementary Table S6c. Ethics, consent and governance of source datasets.*

| Dataset or cohort | Ethics or approval | Consent or governance | Provenance |
|---|---|---|---|
| NHES II (1963-1965) | Historical NCHS human-subject oversight; no modern-format IRB approval number identified | Original consent record not located | NCHS national examination survey; official public-use data |
| NHES III (1966-1970) | Same historical NCHS framework | Original consent or parental-permission record not located | NCHS national examination survey; official public-use data |
| NHANES I (1971-1974) | Historical NCHS human-subject oversight | Original consent documentation not located | NCHS national health examination survey; official public-use data |
| NHANES II (1976-1980) | Historical NCHS human-subject oversight | Original consent documentation not located | NCHS national health examination survey; official public-use data |
| HHANES (1982-1984) | Exact historical review record not located | Original consent documentation not located | NCHS Hispanic Health and Nutrition Examination Survey; official public-use data |
| NHANES 1999-2004 | NCHS Protocol #98-12 | Informed consent; parental permission and child assent documented | Official CDC/NCHS public-use files |
| NHANES 2005-2010 | NCHS Protocol #2005-06 | Informed consent framework documented | Official CDC/NCHS public-use files |
| NHANES 2011-2016 | NCHS Protocol #2011-17 | Adult consent plus child assent and parental permission | Official CDC/NCHS public-use files |
| NHANES 2017-March 2020 | Protocol #2011-17, then #2018-01 from 26 October 2017 | Written consent; parental consent and assent for minors | Official CDC/NCHS pre-pandemic release |
| KNHANES 2009-2012 | KCDC IRB; reported references 2009-01CON-03-2C, 2010-02CON-21-C, 2011-02CON-06-C, 2012-01EXP-01-2C | Consent and governance documented at survey level. | Official KNHANES/KDCA raw-data system |
| KNHANES 2019-2023 | KDCA IRB; reported references for 2019-2022: 2018-01-03-C-A, 2018-01-03-2C-A, 2018-01-03-5C-A and 2018-01-03-4C-A; 2023 approval number not located in the sources reviewed. | Consent and governance documented at survey level. | Official KNHANES/KDCA raw-data system |
| NIOSH OHL, 1981-2010 | NIOSH IRB determined the programme was research not involving human subjects because audiograms were de-identified | Occupational and regulatory audiograms; de-identified before transfer to NIOSH | Audiometric service providers, occupational clinics, hospitals and others, supplied to the NIOSH OHL surveillance dataset [8] |
| OHHR, Germany | Local ethics committee, Carl von Ossietzky University Oldenburg, reviewed and authorised the 2013-2015 collection; approval number not reported in the descriptor article [10] | Original consent; about 40% of participants re-consented to publication of anonymised data and a waiver of consent for data sharing was granted by the data protection officer for the remainder after anonymisation [10] | 2013-2015 research cohort; anonymised public dataset, version 1.5.0.0; data-protection approval DSM-H4A Open Dataset/20241113-0009 [9,10] |
| Münster glycerol series (Basel and Lütkenhöner) | Formal ethics approval, exemption or waiver was not reported in the 2013 Ear and Hearing article, the 2014 reappraisal, or the Dryad dataset record [11,12,13]. At the time, medical research involving humans at Münster fell within the remit of the joint Ärztekammer Westfalen-Lippe and University of Münster Ethics Committee under the applicable professional rules | The glycerol tests were performed as part of clinical diagnostic care. Research-consent or waiver documentation for the later retrospective analysis was not reported. Archived clinical audiograms were subsequently transcribed into computer-readable form for retrospective analysis and publicly deposited by the investigators [12] | ENT Clinic, Münster University Hospital; clinical glycerol tests performed 1999-2009 in patients with suspected Ménière's disease; 347 patients, 356 ears; same data reused in the 2014 analysis [11,12,13] |
| BEAR, Denmark | Science-Ethics Committee, Capital Region Denmark, H-16036391 | Written informed consent | BEAR hearing-aid research cohort; public Zenodo data, version 1.1 [14] |
| Regev DTU 25134611 | Science-Ethics Committee, Capital Region Denmark, H-16036391 [15] | Written informed consent [15] | DTU experimental hearing cohort [15] |
| Regev DTU 25771884 | Same committee, H-16036391 [16] | Written informed consent [16] | DTU follow-up cohort; participant overlap with earlier Regev work documented [16] |
| Meltem Hospital, Türkiye | Private Meltem Hospital Clinical Research Ethics Committee, No. 47, 18 March 2024 | Consent or waiver detail not documented in the sources reviewed | Meltem Hospital audiograms; the deposit states that permission for data sharing was granted by the hospital [17] |
| Cadenza CAD1, UK | University of Leeds REC, FAHC 21-125 | Written informed consent plus consent for publication | Real-listener evaluation data; anonymised. The listener audiogram file is identical in deposit versions 1.0.0 and 1.1.0 [18] |
| Macquarie (Izmaylova), Australia | Macquarie University Ethics Committee, 5201956309964 | Consent wording not located in the sources reviewed; participants were recruited for neurophysiological research | 121 participants tested, 117 in the analytical sample; Dryad deposit [19] |
| Urmia, Iran | Urmia University of Medical Sciences, IR.UMSU.REC.1402.284 | Written consent not required for retrospective study | Retrospective audiometry and tinnitus cohort, Imam Khomeini Hospital, Urmia; Urmia University of Medical Sciences ethics approval IR.UMSU.REC.1402.284; anonymised dataset subsequently distributed through a public Kaggle deposit [20] |

Bracketed numbers cite the Supplement references list. A dataset citation supports the identity and provenance of the deposit; it is used for an ethics or consent statement only where the repository record itself carries that statement.

Absence of a located record is reported as such. It is not evidence that approval, exemption or consent was absent, and historical cohorts are described in the terms of their own time. No approval number has been inferred or supplied where none was located.

For the Münster series, the existence of the institutional ethics oversight framework is stated; no specific approval, exemption or waiver decision has been located and none is attributed to that committee. KNHANES primary annual approval records remain to be retrieved; the references shown are as reported at survey level.
